# Interoceptive Performance is Unrelated to Mental Health Symptoms: Evidence From a Large Scale Multi-Domain Psychophysical Investigation

**DOI:** 10.1101/2025.08.25.25334366

**Authors:** Leah Banellis, Niia Nikolova, Jesper Fischer Ehmsen, Arthur S. Courtin, Melina Vejlø, Ashley Tyrer, Rebecca A. Böhme, Francesco Bavato, Kelly Hoogervorst, Francesca Fardo, Micah G. Allen

**Author notes:** **Materials and correspondence** Corresponding Author: Leah Banellis,.

## Abstract

Interoception—the sensing and perception of the internal viscera—is widely cast as a transdiagnostic mechanism linking brain–body interaction to mental illness. Prevailing models propose that altered interoceptive performance is associated with psychiatric vulnerability across a range of symptoms. We tested this hypothesis in a large community sample using psychophysically optimised tasks spanning cardiac and respiratory domains (N = 456 cardiac, N = 245 respiratory), combined with hierarchical Bayesian modelling and comprehensive symptom profiling. Contrary to this central prediction, objective interoceptive performance metrics spanning sensitivity, precision, and metacognition were largely unrelated to general symptom burden or specific mental health dimensions across linear, categorical, and network-based models. In contrast, self-reported interoceptive sensibility showed moderate associations with symptoms, but semantic similarity analyses suggest these reflect higher order interpretative or affective beliefs rather than interoceptive specific processing. These findings challenge the prevailing view that objective interoceptive sensitivity is a broad marker of psychopathology, prompting a reconsideration of how we measure and interpret interoception in mental health research.

## Introduction

Interoception, the processing and perception of internal bodily states like heartbeat, breathing, and gastric activity, is increasingly cast as pivotal to mental health^1,2^. Neuropsychiatric research has intensified around this idea, with studies reporting interoceptive hypersensitivity in anxiety and panic disorders, and hypo-responsiveness in conditions such as depression, autism spectrum, and attention deficit hyperactivity disorder (ADHD)^3–6^. These findings have led to growing interest in interoception as a candidate clinical marker, which proposes interoceptive dysfunction as a core transdiagnostic factor of psychopathology itself^7–9^. Yet despite this theoretical momentum, empirical support for a broad interoception– psychopathology link remains inconsistent and methodologically constrained.

Numerous studies using behavioural interoceptive tasks have reported heightened or diminished interoception across psychiatric conditions, with anxiety and panic often associated with heightened interoceptive sensitivity^5,10–15^, and depression, autism, and ADHD linked to blunted or impaired interoceptive performance^3,4,6^. However, most of this work has narrowly focused on heartbeat perception as overgeneralised proxies for interoception as a whole^16^. Despite reported associations between interoceptive dysfunction and mental illness, meta-analytic findings highlight no robust link across interoceptive performance studies^17–21^. These discrepancies may partially reflect methodological limitations of commonly used cardiac perception tasks, including poor construct and convergent validity^22–26^. Furthermore, prior studies have relied on simple accuracy indices that conflate perceptual sensitivity with response bias, and provide limited insight into interoceptive precision and reliability^27,28^, which are theorised to be distinct and important components of interoceptive processing in mental health^29–31^. Other bodily domains, such as respiratory interoception, remain relatively underexplored, despite theoretical and preliminary evidence suggesting an equal or greater role in anxiety and related psychopathology^32–35^. Self-report measures of interoceptive sensibility^36^ often yield correlations with mental health symptoms^37– 39^, but remain limited by attentional biases, semantic beliefs, and weak correspondence among survey instruments^36,40–42^. Together, these issues underscore the need for a comprehensive, multimodal approach to clarify whether and how interoception relates to general mental health across sensory domains and representational levels.

Dimensional models of psychopathology, including ‘the p factor’ (i.e., general factor of psychopathology), offer a powerful alternative to traditional diagnosis by capturing shared variance across diverse symptoms such as depression, anxiety, obsessive compulsive disorder (OCD), and schizophrenic symptoms^31,43,44^. These dimensional approaches align with evidence linking transdiagnostic symptom variation to neural and cognitive functioning in the continuum of healthy to pathological mental states^45– 48^. This parallels the shift in computational psychiatry away from categorical approaches towards continuous, biologically grounded models of mental dysfunction^49,50^. Studying these associations in non-clinical samples is critical for identifying trans-symptomatic features that operate across the spectrum of mental health, as it could enable the identification of early vulnerability markers, offering a crucial window for preventative intervention^51,52^. Notably, interoception can be modified through training and feedback, raising the possibility of targeted intervention strategies for individuals at risk^53,54^. However, whether interoceptive function relates to general symptom burden, before the onset of clinical illness, remains an open question.

To address these uncertainties, we conducted a multimodal psychophysical investigation of interoceptive performance and mental health in a large, predominantly non-clinical sample (N=456 cardiac, N=245 respiratory). We administered psychophysical interoception tasks targeting both cardiac (heart rate discrimination) and respiratory (breathing resistance sensitivity) interoception, allowing us to separately quantify interoceptive sensitivity, precision, and metacognition, addressing psychometric limitations of previous studies. We additionally collected self-report measures of interoceptive sensibility and administered a comprehensive mental health battery spanning depression, anxiety, sleep, fatigue, autism spectrum, and ADHD symptoms. Using computational factor analysis methods^46^, we derived latent mental health factors to index general symptom burden. Our multi-domain, high-powered design allowed us to robustly test whether core dimensions of interoceptive ability relate to general psychopathological dimensions, and whether such links reflect genuine alterations in bodily sensing or methodological confounds in assessing interoception. Consistent with transdiagnostic accounts that posit interoceptive dysfunction as a core feature of psychopathology^7–9^, we hypothesised that interoceptive sensing and beliefs would be associated with general symptom burden across multiple domains. Specifically, we expected that both behavioural indices of interoceptive performance (sensitivity, precision, and metacognition) and self-reported interoceptive sensibility would show relationships with latent dimensions of psychopathology, reflecting a transdiagnostic interoceptive mechanism.

## Results

### Overview of mental health and interoception analyses

To investigate the relationship between interoceptive ability and mental health, we analysed data from a large, predominantly non-clinical sample who completed a multimodal interoceptive test battery and an extensive psychiatric assessment. Exploratory factor analysis of individual symptom items yielded eleven first-level and two higher-level latent mental health factors spanning trans-symptomatic dimensions. The interoceptive test battery included a heart rate discrimination task (HRDT)^55^ and a respiratory resistance sensitivity task (RRST)^56^. Hierarchical Bayesian models were used to estimate perceptual metrics of interoceptive sensitivity (threshold) and interoceptive precision (slope). For further information on the demonstrated reliability and parameter recovery of the hierarchical modelling approach used here, see Courtin et al. (2025)^57^. Metacognition was assessed via confidence ratings and signal detection theoretic models^58,59^, yielding indices of metacognitive bias (mean confidence) and metacognitive efficiency (M-Ratio) (see Supplementary Table 1 for descriptive statistics). Subjective interoception (interoceptive sensibility) was assessed using the eight subscales of the Multidimensional Assessment of Interoceptive Awareness (MAIA) questionnaire (see Supplementary Table 2 for descriptive statistics). We tested associations between interoceptive ability and psychopathology (N=456 cardiac, N=245 respiratory; 91% participant overlap of RRST with HRDT) using FDR-corrected Spearman correlations, with Bayesian hypothesis testing applied to evaluate evidence for effects and null results.

### Mental health exploratory factor analysis

We performed item-level exploratory factor analysis (EFA) on the survey responses to uncover latent mental health variables. This is a common approach to identify a statistical and descriptive summary of the covariance structure within a high-dimensional symptom space^31,45–47,60^. In total 47 multivariate outliers were detected and removed via Mahalanobis distance, resulting in 500 full-dataset remaining participants. Bartlett’s test of sphericity (X^2^(6328) = 28393.24, p < .001) and the Kaiser-Meyer-Olkin (KMO) statistic (MSA = 0.921) confirmed that the correlation matrix of 113 survey responses was non-random, factorable, and thus appropriate for EFA. The scree plot and parallel analysis suggested 11 factors to retain. We aimed to achieve a very simple structure (VSS) by removing items which did not load on any item or cross-loaded on multiple items, facilitating a theoretically meaningful factor solution^61^.

### Multi-level mental health factor solution

The 11 VSS factor solution of 80 items revealed the following factors: social anxiety, self-confidence, sleep, inattentiveness, depression, negative thoughts, restlessness, stress, autism spectrum, somatic symptoms, and impulsivity (see Figure 2, Supplementary Table 3 for pattern coefficients, and Supplementary Table 4 for item details in each factor). The fit indices were between acceptable and excellent (TLI = 0.892, CFI = 0.921, RMSEA = 0.033). The reliability of each of the 11 factors was between acceptable and excellent (*α range* = 0.707 - 0.925, *ω range* = 0.729 - 0.936), as indicated by Cronbach’s alpha and total omega coefficients.

**Figure 1:**
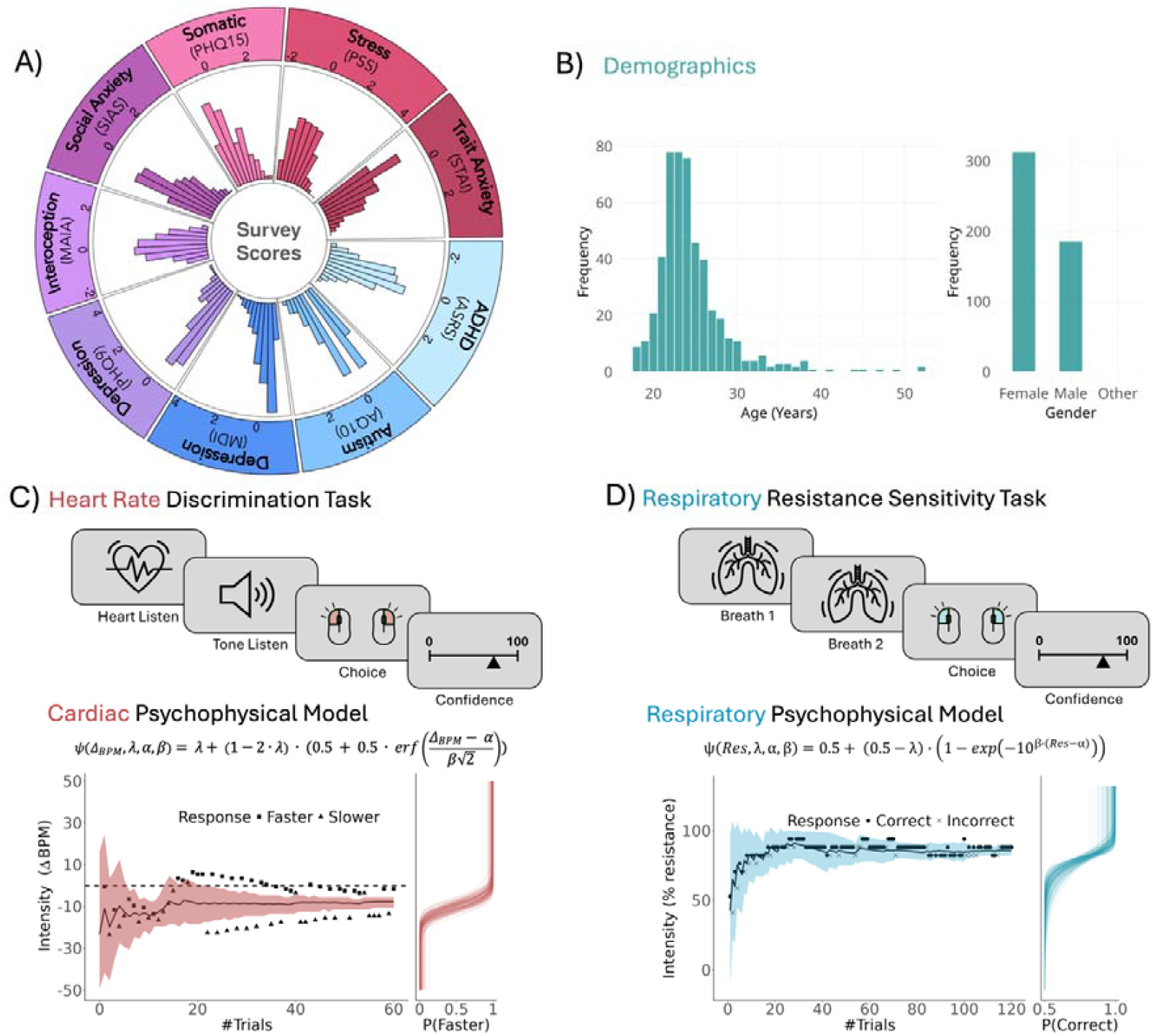
Mental health assessment and multimodal interoception test battery. A) Normalised distributions of mental health survey scores across eight instruments (autism spectrum (AQ10), ADHD (ASRS), depression (MDI and PHQ9), somatic symptoms (PHQ15), stress (PSS), social anxiety (SIAS), trait anxiety (STAI trait)), and interoceptive sensibility (MAIA). B) Age and gender demographics distributions of the N = 500 sample for the mental health factor analysis. Participants performed two interoceptive psychophysical tasks of two sensory modalities: C) the Heart Rate Discrimination Task (HRDT), D) the Respiratory Resistance Sensitivity Task (RRST). Each task employed an adaptive psychophysical model estimating threshold (α), precision (β), and lapse rate (λ). In the HRDT, participants compared the rate of auditory tones to their real-time heart rate (via pulse oximeter) and rated confidence. In the RRST, participants inhaled twice through a respiratory circuit, judged which breath felt more difficult, and reported their confidence via visual analogue scale. In all conditions, the stimulus intensity was adaptively adjusted using the Psi algorithm; see Methods for more details. Example plots (bottom) display the stimulus adjustments (respiratory resistance, tone frequency) across trials for a single participant, alongside the psychometric fits that quantify interoceptive (cardiac or respiratory) performance.

**Figure 2:**
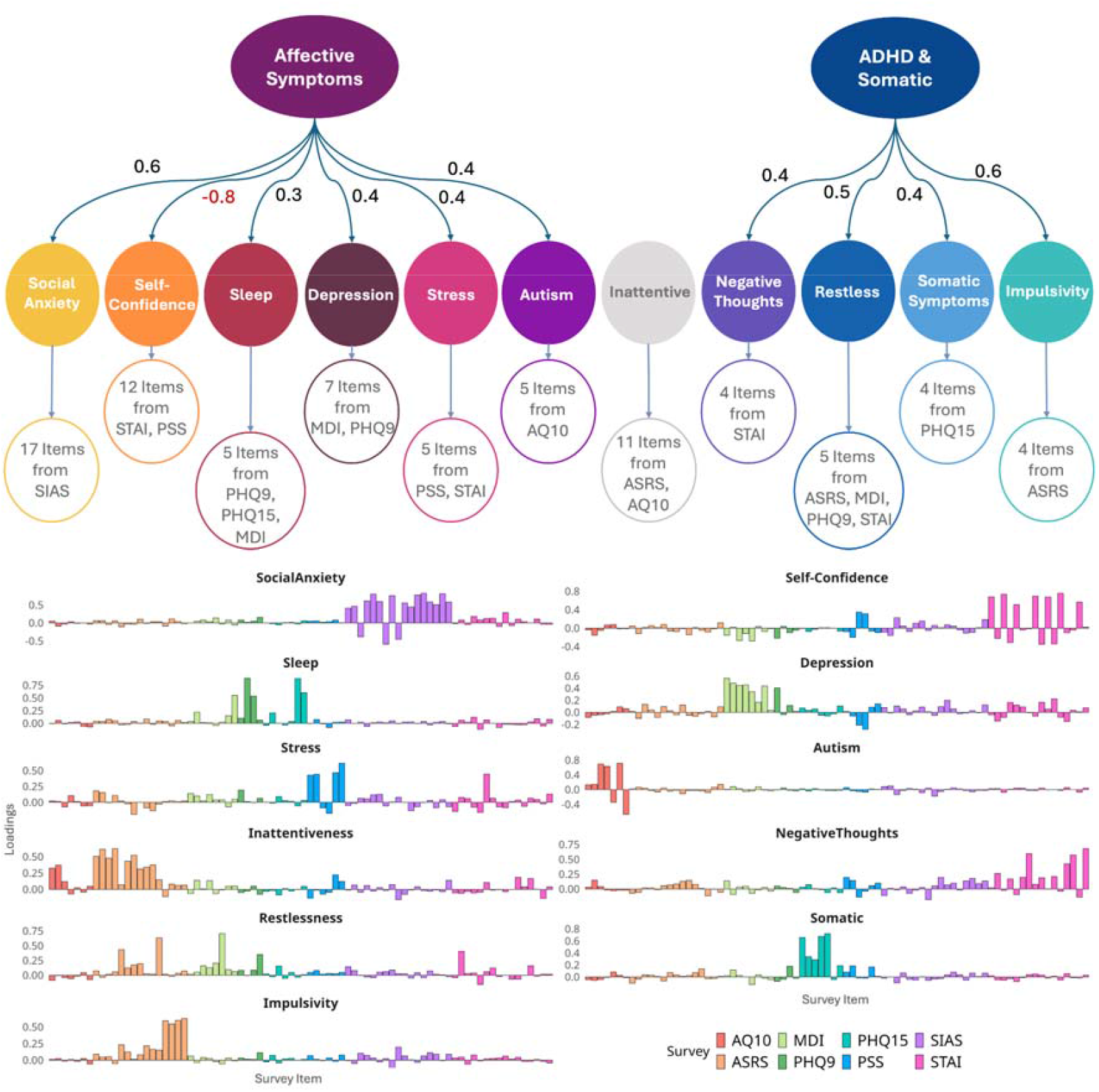
Multi-level factor solution of mental health dimensions. Exploratory Factor Analysis (EFA) of items from eight mental health surveys. The top figure shows the EFA path diagram, revealing 11 lower-level factors: social anxiety, self-confidence, sleep, inattentiveness, depression, negative thoughts, restlessness, stress, autism spectrum, somatic symptoms, and impulsivity, as well as two higher-level factors: affective symptoms, and ADHD & somatic symptoms. The bottom shows the lower-level factor loadings (pattern coefficients) on the inputted survey items (autism spectrum (AQ10), ADHD (ASRS), depression (MDI and PHQ9), somatic symptoms (PHQ15), stress (PSS), social anxiety (SIAS), and trait anxiety (STAI trait)). For the first-level factor analysis: see Supplementary Table 3 for full item pattern coefficients, and Supplementary Table 4 for a full list of items in each factor. For the higher-level factor analysis: see Supplementary Table 5 for pattern coefficients, and Supplementary Table 4 for details of lower-level factors in each high-level factor.

Two higher-level factors were estimated based on the factor scores of the 11-factor solution using parallel analysis and the inflexion point of the scree plot. The first higher-level factor encompassed affective symptoms (first level factors: social anxiety, self-confidence (negative loading), sleep, depression, stress, autism spectrum), and the second higher-level factor incorporated ADHD and somatic symptoms (negative thoughts, restlessness, somatic symptoms, impulsivity) (see Figure 2, Supplementary Table 5 for pattern coefficients, and Supplementary Table 4 for lower-level factor details in each high-level factor).

### Null relationship of mental health with interoceptive psychophysics

Interoceptive performance variables, encompassing both perceptual and metacognitive measures, were generally unrelated to mental health factors (see Figure 3), apart from a weak positive correlation of depressive symptoms with cardiac metacognitive efficiency (HRDT M-Ratio) (rs = 0.139, pFDR = 0.024).

**Figure 3:**
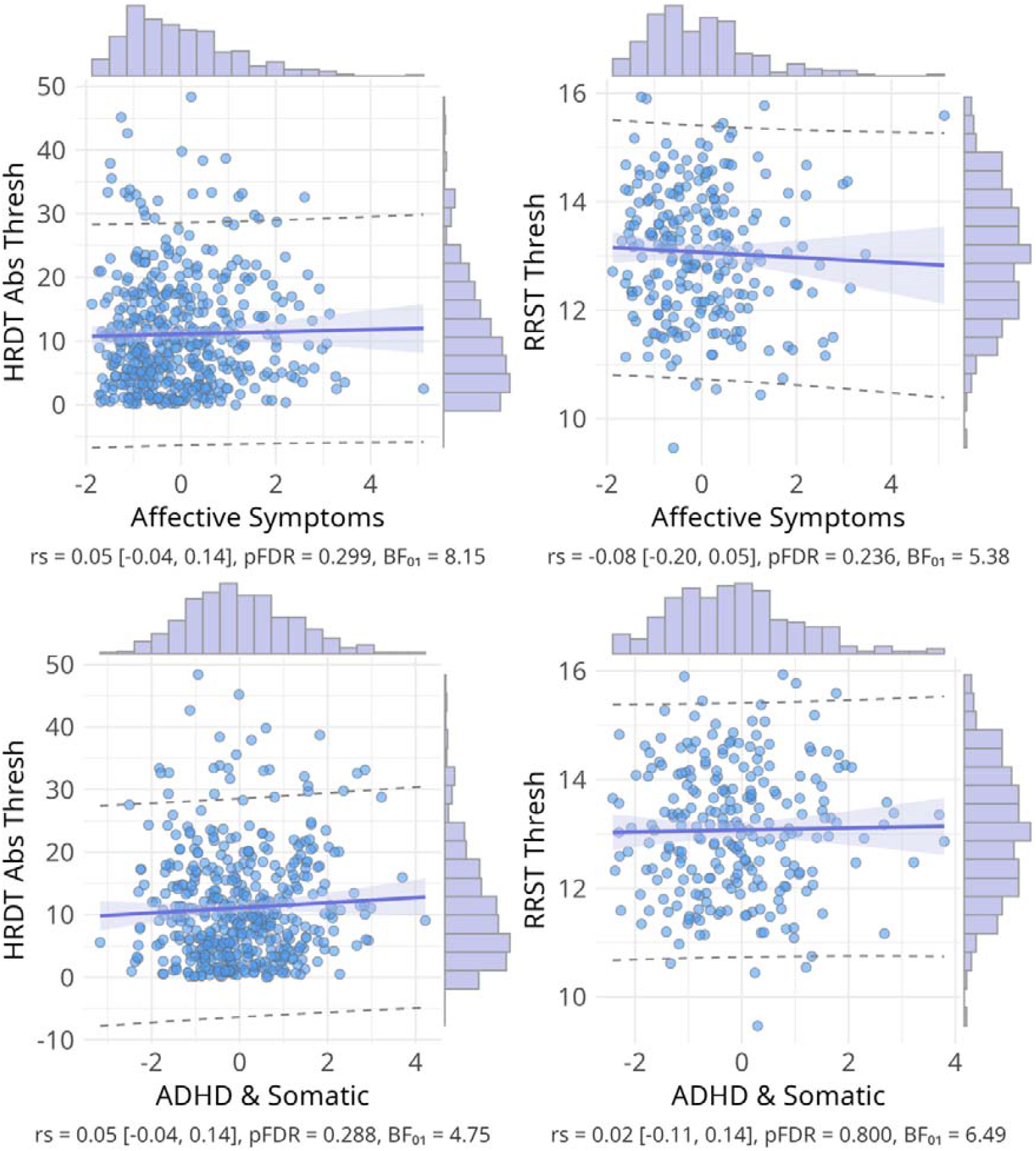
Scatter plots of the null relationship between interoceptive performance (threshold) and mental health higher-level factors. Scatter plots of Spearman correlations across interoception task variables and mental health higher-level factors. Interoceptive performance is demonstrated via sensitivity for the cardiac (HRDT absolute threshold) and respiratory (RRST threshold) psychophysical tasks. For mental health correlations with interoceptive precision (slope) see Supplementary Figure 1. Above shows statistics for Spearman correlations with 95% confidence intervals in square brackets, p-values are corrected for multiple comparisons using the false discovery rate (Benjamini-Hochberg procedure at < 0.05), and null bayes factors (BF01).

The rest of the interoceptive mental health correlations were not significant once corrected for multiple comparisons (see Figure 4). Notably, applying a more liberal multiple-comparison correction restricted to only the interoceptive psychophysics– mental health test family (rather than the full correlation matrix) yielded no significant associations, including the previously observed weak depressive symptom–cardiac metacognition correlation. In general, Bayes Factor analysis provided anecdotal to moderate evidence in favour of a lack of relationship between mental health and interoceptive performance^62^, with slightly stronger evidence observed for HRDT variables compared to RRST variables. Partial correlations controlling for age, gender, and body mass index reveal consistent significant interoceptive mental health correlations (see Supplementary Figure 2). Full cross-correlation and Bayes Factor matrices are provided in Supplementary Figures 3 and 4. We also observed no significant correlations of mental health dimensions with task accuracy in all sensory modalities (see Supplementary Figure 21). For mental health correlations with the exteroceptive auditory control condition, a single significant association was observed between autism traits and greater auditory discrimination sensitivity (see Supplementary Figures 5-7).

**Figure 4:**
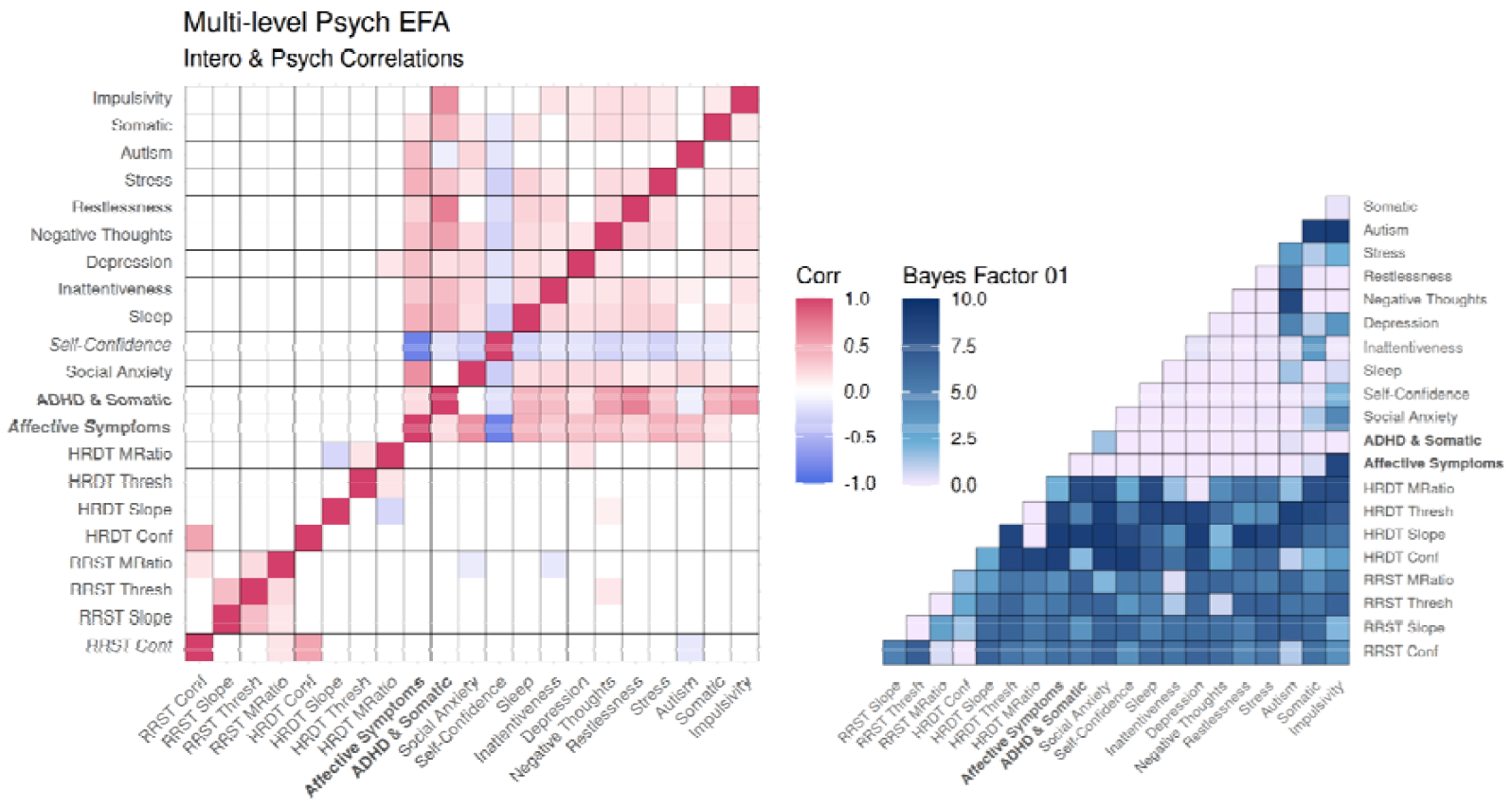
Correlations between interoceptive psychophysics and mental health factors. Left: Cross Spearman correlations across interoception task variables and mental health factors (11 lower-level and two higher-level EFA). Interoceptive task variables include sensitivity (threshold (absolute for HRDT)), precision (slope), metacognitive bias (mean confidence), and metacognitive efficiency (M-Ratio) across cardiac (HRDT) and respiratory (RRST) domains. The upper diagonal depicts correlations that survive false discovery rate correction for multiple comparisons (Benjamini-Hochberg procedure at < 0.05 shown in upper diagonal coloured boxes), whereas the lower diagonal depicts correlation coefficients significant at the uncorrected threshold (uncorrected p < .05; lower diagonal coloured boxes). In red are significant positive correlations and in blue are significant negative correlations (p < 0.05 shown in coloured boxes). Right: Heatmap depicting null bayes factors (BF01) for interoceptive performance and mental health factors. The majority of observed Null Bayes Factors show anecdotal (BF01 1 - 3 in light blue) or moderate (BF01 3 - 10 in dark blue) evidence for a lack of association^62^, with slightly stronger null evidence for cardiac variables compared to respiratory variables^63^. However, some exceptions include anecdotal Bayesian evidence for the alternative hypothesis (BF01 0.33 - 1 in light purple) with inattentiveness, autism, and negative thoughts factors with interoception metrics (as well as BF01=0.07 for the depressive symptoms with cardiac metacognition relationship described above).

These null patterns remained consistent when using psychiatric survey sum scores instead of latent factors. The only notable exception was a weak negative correlation between cardiac precision (HRDT slope) and autism spectrum traits, as measured by the AQ10 (rs = -0.110, pFDR = 0.013) (see Supplementary Figure 8).

### Relationship of mental health with interoceptive sensibility (MAIA)

In contrast, we observed pervasive moderate correlations between interoceptive sensibility, as measured by the MAIA subscales and mental health factors (see Figure 5). The strongest and most consistent association was found between the bodily trusting subscale and all the mental health factors. These bodily-trusting correlations were predominantly negative (except for the positive correlation with the self-confidence factor), indicating that worse mental health (i.e., higher depressive symptoms, sleep problems, negative thoughts, and lower self-confidence etc) is associated with less subjective ‘experience of one’s body as safe and trustworthy’ (affective symptoms factor with MAIA Trusting subscale: rs = -0.548, pFDR < .001).

**Figure 5:**
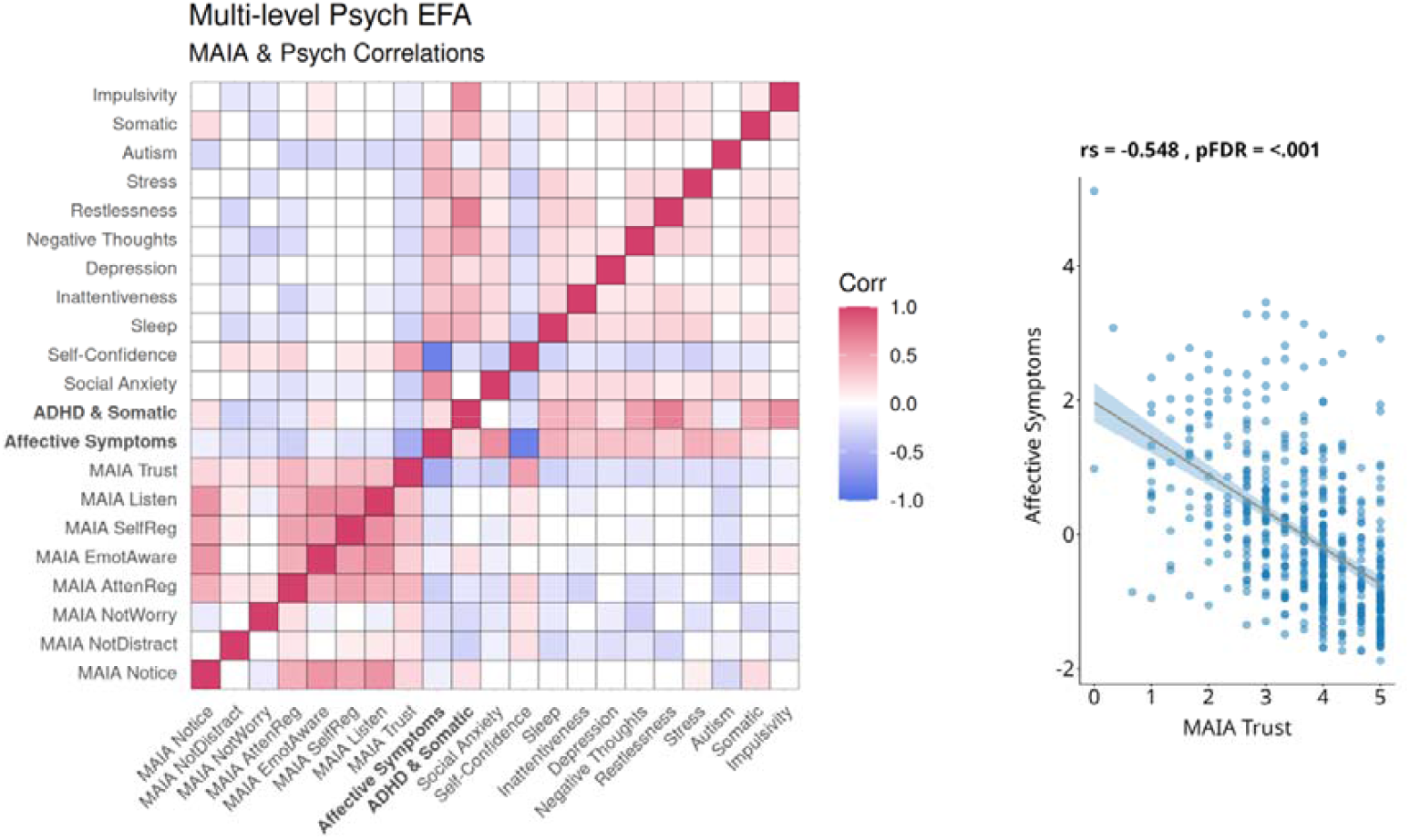
Correlations between interoceptive sensibility and mental health factors. Left: Cross Spearman correlations between the 8 MAIA subscales (interoceptive sensibility) and the multi-level mental health factors (11 lower-level and two higher-level EFA). The upper diagonal depicts correlations that survive false discovery rate correction for multiple comparisons (Benjamini-Hochberg procedure at < 0.05 shown in upper diagonal coloured boxes), whereas the lower diagonal depicts correlation coefficients significant at the uncorrected threshold (uncorrected p < .05; lower diagonal coloured boxes). In red are significant positive correlations and in blue are significant negative correlations (p < 0.05 shown in coloured boxes). Right: scatter plot of the mean MAIA score across all eight subscales (interoceptive sensibility) and the higher-level ‘affective symptoms’ factor from the multilevel factor analysis, above shows the Spearman correlation statistics. The negative correlation indicates the relationship of worse mental health (higher depressive symptoms, sleep problems, negative thoughts, and lower self-confidence etc) with a reduced subjective sense of internal bodily sensing and reduced subjective attentional/emotional control of this sensing. Heatmap of the corresponding null Bayes factors (BF_01_) can be found in Supplementary Figure 10.

We also observed predominantly negative correlations of mental health factors with the not-distracting, not-worrying, and attention regulation subscales. This further demonstrated that worse mental health (higher self-reported symptoms) was associated with a reduced subjective tendency to not ignore or not worry about painful/discomforting bodily sensations (ADHD & somatic symptoms factor with MAIA Not-Distracting: rs = -0.293, pFDR < .001, ADHD & somatic symptoms factor with MAIA Not-Worrying: -0.239, pFDR < .001), and a reduced subjective ability to control their attention to bodily sensations (affective symptoms factor with MAIA Attentional Regulation: rs = -0.305, pFDR < .001).

Of the mental health factors, autism spectrum and self-confidence displayed the most persistent correlations across the MAIA sensibility subscales. Additionally, the higher-order factors, affective symptoms and ADHD & somatic symptoms, displayed moderate correlations with nearly all the interoceptive sensibility subscales. See Supplementary Figure 2 for equivalent partial correlations controlling for age, gender, and body mass index. Full cross-correlation and Bayes Factor matrices are provided in Supplementary Figures 9 and 10.

### Relationship of objective (HRDT/RRST) and subjective (MAIA) interoception

When comparing psychophysical-task and self-report measures, we again found a robust pattern of no association between either interoception task or the MAIA sub-scales (see Supplementary Figure 11). The one exception was with respiratory and cardiac average confidence, which was positively related to the following MAIA subscales: Attention Regulation, Emotional Awareness, Self-Regulation, and Body Listening subscales (as well as the MAIA Noticing subscale with HRDT cardiac confidence), with similar correlation coefficients for both tasks (rs range = 0.146 - 0.212, pFDR range = 0.012 - < .001). Null Bayes factor analysis found generally between anecdotal (BF01 > 1) to moderate (BF01 > 3) evidence for the lack of association between the objective task and subjective survey interoception measures^62^. The highest Bayesian evidence for a null relationship was between cardiac sensitivity (HRDT threshold) and the MAIA subscales (BF01 range = 6.85 - 9.57 for the 8 MAIA subscales). Additional analyses controlling for age, gender, and body mass index yielded equivalent results (see Supplementary Figure 2). Full cross-correlation and Bayes Factor matrices are provided in Supplementary Figures 12 and 13.

### Clinical severity analyses

To assess whether interoceptive–mental health associations emerged selectively at higher symptom severity, we conducted clinical cut-off group comparisons and upper symptom percentile correlation analyses. Participants survey scores were stratified using established clinical screening thresholds (see Supplementary Table 7 and 8), and interoceptive performance measures were compared between groups above versus below each cut-off using Mann–Whitney U tests (see Supplementary Table 9). Although a small number of uncorrected effects were observed (autism traits with cardiac/respiratory confidence (HRDT confidence: U = 13476, pFDR = .094, rank-biserial r = 0.20; RRST confidence: U = 3715, pFDR = .073, rank-biserial r = 0.33), and ADHD symptoms with cardiac precision and respiratory sensitivity (HRDT slope: U = 22521, pFDR = .169, rank-biserial r = 0.13; RRST threshold: U = 6151, pFDR = .169, rank-biserial r = 0.22)), none survived multiple comparisons correction (see Supplementary Figures 14-15).

In parallel, percentile-based correlation analyses restricted symptom scores to the top 50%, 75%, and 90% of the sample. While autism spectrum traits correlated with lower confidence in the cardiac (top 50% analysis) and respiratory (top 75% analysis) task, these analyses revealed no consistent strengthening of interoceptive–symptom associations with increasing symptom severity, and no effects survived correction for multiple comparisons at the highest 90% symptom percentile (see Supplementary Figure 16). Together, these results provide no evidence that interoceptive psychophysics measures (sensitivity, precision, or metacognition) are selectively associated with mental health symptoms at higher severity levels, but future research should investigate clinically diagnosed patients at the highest severity levels.

### Network mental health model

To assess whether observed interoceptive–mental health relationships were consistent across alternative representations of psychopathology, we conducted an item-level psychiatric symptom network analysis. Using a spinglass community detection algorithm, we identified 9 clusters of psychiatric symptoms spanning self/social-confidence, restlessness, attentional dysregulation, mentalising, impulsivity, depressive symptoms, somatic symptoms, stress reactivity, and social anxiety (see Supplementary Figure 17 and Supplementary Table 10 for item details for each network community). Crucially, examining interoceptive ability associations within this network framework yielded a pattern largely consistent with the factor-analytic results, with minimal evidence of interoceptive–mental health relationships. This was in the exception to the correlations of the autistic trait mentalising with respiratory confidence (RRST Conf rs = -0.169, pFDR = 0.024) and cardiac metacognition (HRDT M-Ratio: rs = 0.132, pFDR = 0.031) (see Supplementary Figure 18). This convergence of primarily null results highlights robustness across both latent-factor and network-based representations of mental health.

### Mental health and interoception network analysis

In a joint network embedding interoceptive psychophysics, interoceptive sensibility (MAIA), and mental health network communities (see Supplementary Figure 18), An EBICglasso-estimated network yielded a sparse pattern of cross-domain connections. Direct associations between mental health domains and interoceptive psychophysics variables were weak and infrequent (partial r ≤ .02), indicating minimal direct coupling after controlling for all other variables (see Supplementary Figure 19 and Supplementary Table 11).

In contrast, several MAIA subscales showed moderate associations with mental health symptoms, particularly involving self/social-confidence with MAIA trusting subscale, stress reactivity/somatic symptoms with MAIA not-worrying subscale, and attentional/mentalising-related domains with MAIA attentional regulation subscale. Bridge strength analysis further indicated that interoceptive sensibility nodes (MAIA subscales) exhibited substantially greater cross-community connectivity than interoceptive psychophysics nodes (HRDT and RRST parameters), with MAIA subscales trusting, not-worrying, not-distracting, attentional regulation, and emotional regulation subscales emerging as the strongest bridges (see Supplementary Table 12).

Notably, most interoceptive psychophysics measures showed near-zero bridge strength, suggesting that objective interoceptive psychophysical performance was largely decoupled from psychiatric symptom domains within the structure of the network (see Supplementary Table 12). Bootstrap analyses demonstrated that mental health–psychophysics edges were consistently stable near zero, whereas mental health–interoceptive sensibility edges showed greater dispersion across resamples, indicating comparatively stronger cross-domain coupling.

### Semantic similarity analysis of MAIA

The MAIA items cluster semantically with a heterogeneous set of existing questionnaires. The strongest overlaps are with scales on sexual objectification, transgender congruence, generic body-awareness, hypochondriacal depression, physical health functioning, health orientation, mindfulness skills, death-related disgust, depressive mood, somatisation, femininity ideology, somatic anxiety and therapist countertransference (all cosine similarity > 0.4). This suggests that MAIA items may tap into broader constructs related to body image, psychopathology and general health attitudes, rather than uniquely capturing interoceptive sensibility, as shown by a latent semantic analysis of over 4,000 scales^64^. This analysis reflects shared lexical content and should be interpreted as an index of potential construct overlap. Therefore, such diffuse semantic neighbourhood is consistent with limited construct specificity at the lexical level (see Supplementary Table 13). See Figure 6 word cloud sized by cosine similarity of the MAIA survey to other questionnaires (bigger = more similar). Terms in the word cloud reflect lexical neighbours derived from an external corpus. The word cloud illustrates that the lexical content of MAIA drifts toward disparate constructs (objectification, transgender identity, depression, somatisation, anxiety) instead of coalescing around a single coherent interoception theme (see Figure 6). We also investigated semantic similarity separately for all eight MAIA subscales. All subscales, including those most strongly related to the mental health factors in the present study, show substantial semantic overlap with non-interoceptive constructs (Supplementary Table 14 and Supplementary Figure 20).

**Figure 6:**
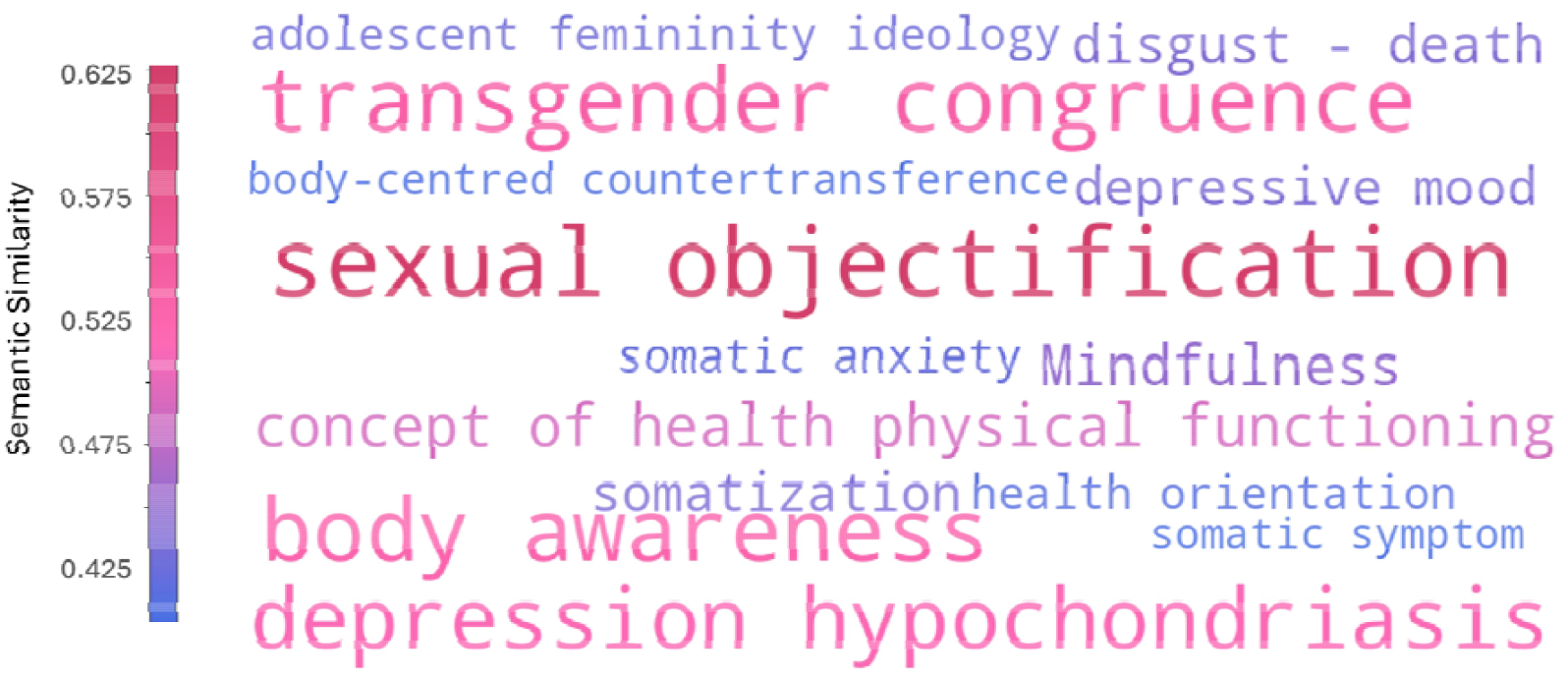
Latent Semantic Similarity Analysis of MAIA survey. Word cloud of latent semantic similarity analysis of the Multidimensional Assessment of Interoceptive Awareness (MAIA) questionnaire items in relation to other psychological scales, using the online Semantic Scale Network tool^64^. The size and colour of the words represent the cosine similarity of the MAIA survey to other psychological questionnaires, with larger pink/red words indicating higher similarity. Therefore, terms reflect lexical neighbours derived from an external corpus of 4,000 scales.

## Discussion

We tested whether core dimensions of interoceptive performance relate to transdiagnostic dimensions of psychopathology in a large, predominantly non-clinical cohort^43–48^. Contrary to the broad transdiagnostic hypothesis of interoceptive ability in mental health, we observed no robust evidence that interoceptive task performance indexes general symptom burden or specific symptom categories in this community sample. This was consistent across cardiac and respiratory modalities, spanning sensitivity, precision, and metacognitive interoceptive performance. The sole exception was a weak correlation of depressive symptoms with cardiac metacognition. This pattern converges with subclinical and clinical meta-analytic evidence suggesting minimal or inconsistent links between interoceptive accuracy and anxiety, depression, and autism spectrum^18–21^, challenging the notion that altered interoceptive performance at rest constitutes a transdiagnostic marker of general psychopathology^4,5,10,11,13,14^.

Although we observed largely null relationships between mental health and interoceptive psychophysics, we found a weak association in which higher cardiac metacognitive efficiency correlated with heightened depressive symptoms. To our knowledge, no previous studies have reported this link specifically. While some depression studies report weak impairment in interoceptive accuracy^4^, a large meta-analysis found no overall relationship such that even trend level associations accounted for just 0.6% of symptom variance^4,18^. Our result, which was in the opposite direction, accounted for only 1.93% of symptom variance and did not survive even a more liberal correction for multiple comparisons. It should therefore be interpreted with caution.

In other cases where interoceptive mental health correlations emerged, none survived correction for multiple comparisons. Bayesian analyses overall supported the null hypothesis, mirroring earlier findings that heartbeat detection tasks often fail to correlate significantly with mental health outcomes^18,20,22^. Conflicting reports in the literature about whether various subclinical and clinical mental health dimensions such as anxiety^18,20^, autism spectrum^6,19^, depression^18,21^, and ADHD^3^ correlate positively or negatively with interoceptive accuracy may reflect known psychometric limitations of interoceptive tasks^22,24–26^. Beyond psychometrics, mixed previous findings may further reflect task⍰specific biases (e.g., reliance on confounding factors such as timing strategies or prior beliefs), and underpowered designs combined with selective reporting of positive results, all of which can inflate or obscure true interoception–symptom relationships^65^. The present data reinforce these concerns by showing that even a multi-domain psychophysical approach, explicitly separating perceptual bias and precision from simple accuracy measures, and encompassing both cardiac and respiratory modalities, reveals no robust associations with mental health symptoms. Critically, this may suggest that interoceptive dysfunction relevant to mental health may be state-dependent or context-sensitive, and thus not readily detected by resting-state tasks focused on isolated bodily signals, in the absence of affectively meaningful or ecologically valid contexts.

To assess whether interoceptive–mental health associations emerged non-linearly at higher levels of symptom severity or depended on alternative or non-linear representations of psychopathology, we conducted analyses using clinically defined symptom cut-offs, upper symptom percentiles, raw survey sum scores, and an alternative network-based representation. Across all approaches, the central null pattern remained robust: interoceptive psychophysical performance—spanning sensitivity, precision, and metacognitive efficiency—did not systematically or robustly correlate with mental health symptoms. The only recurring exception involved autism-related mentalising features, which showed modest associations with interoceptive confidence, most consistently for respiratory confidence at higher symptom percentiles and within the symptom network, with weaker and less consistent effects observed for cardiac confidence and cardiac metacognition. However, these effects were neither uniformly found across analytic frameworks, nor did they systematically strengthen at higher symptom percentile ranges. Ultimately, future work in clinically diagnosed samples with greater symptom severity is needed to assess whether such effects arise beyond the symptom ranges examined here.

By contrast, self-reported interoceptive sensibility (MAIA subscale scores) showed moderate correlations with several latent symptom factors (i.e., ADHD, somatic and affective symptoms). Specifically, worse mental health was associated with reduced bodily trust and ability to manage attention or anxiety to bodily sensations and pain. In particular, autism spectrum scores showed prominent negative associations with interoceptive sensibility subscales, aligning with some prior findings^66–68^, though contrasting with null overall meta-analytic evidence^6^. These findings suggest that mental health difficulties may relate to altered subjective beliefs, attention, and interpretation of bodily sensations, rather than objective interoceptive sensing of resting physiology.

While self-report measures offer useful information about subjective experience, they lack specificity for interoceptive sensibility, and here showed no relationship to interoceptive sensitivity, cautioning against using self-report as a proxy for objective interoceptive processing. Rather than reflecting a unitary interoceptive construct, this pattern is consistent with a multidimensional framework in which perceptual sensitivity, subjective reports, and metacognitive beliefs can diverge^36^. However, many such instruments mix bodily awareness with emotional, attentional, and affective traits, which creates conceptual and semantic overlap with mental health symptoms. Correlations among self-report measures are also vulnerable to shared method variance and demand characteristics, which can inflate apparent associations.

Our latent semantic similarity analysis showed strong overlap between MAIA items and questionnaires on broad constructs including sexual objectification, transgender identity, depression, hypochondriasis, body image, and general health attitudes^64^. Such overlap may contribute to the poor correspondence observed across self-report interoception instruments^40,69^, raising concerns about construct validity. The latent semantic similarity results suggests that self-report scales may index hypervigilance or semantic overlap with emotional distress^32,41,42^ rather than genuine interoceptive sensibility.

Including respiratory interoceptive measures provided an opportunity to move beyond the traditional cardiac focus but ultimately yielded little evidence for any link between respiratory interoceptive psychophysics and mental health. This contrasted with previous research reporting a relationship between respiratory interoceptive task performance and anxiety^32,33,35^. Although theoretical models propose that respiratory awareness should be highly relevant to anxiety^34^, the null findings across a large sample question whether these proposed relationships hold in broader clinical symptom or community contexts. Note, recent evidence suggests interoceptive performance is largely organ-specific, supporting the view that these represent separable rather than unified interoceptive processes^16,70–72^. This suggests that null associations with mental health in one interoceptive domain cannot be assumed to generalise to another, and that clinical research may need to treat cardiac and respiratory interoception as independent targets for investigation. This pattern echoes long-standing calls for robust, well-powered investigations of interoceptive dysfunction across multiple physiological systems^22,24,26,27^.

Several limitations temper the generalisability of these conclusions. The sample was predominantly Danish, with a skew toward younger women, so results may not extend to older, more diverse, or clinical populations. Future studies should include broader demographics and well-powered clinical groups, including those with diagnosed psychiatric conditions, to determine whether clinically meaningful alterations in interoception emerge at higher levels of symptom severity, particularly in treatment-resistant conditions that may involve qualitatively distinct mechanisms not captured in community samples. Our use of self-report symptom inventories rather than formal diagnostic interviews reflects a common approach aligned with a call for dimensional transdiagnostic schema in mental health research^45,46,61,62^, but remains vulnerable to biases; incorporating more ecological or clinical assessments may provide additional richer information.

A critical point is that in our study interoception was assessed at rest, whereas bodily signal perception has been shown to differ under stress^73^ or physiological arousal^74^. Intuitively, associations between somatic symptom perception and for example anxiety or panic attack, requires assessing interoception during these fluctuating bodily states, during the transition from baseline to arousal and recovery. Our conclusion is that the search for interoceptive markers of clinical symptoms must move beyond the laboratory to study interoception in these more dynamic and ecologically valid scenarios.

Similarly, while our use of psychophysical batteries rules out several competing explanations for our null findings, including our mechanistic specificity for delineating sensitivity, bias, and precision, it remains plausible that matching task and symptom domain more carefully (e.g., gastric interoception for patients with gastric-related symptoms) might reveal stronger associations. Nevertheless, our results challenge the common assumption that lab-based, resting-state measures of interoception are strongly related to mood and affective traits. In our data, these measures showed limited association with the symptom dimensions assessed here. Our work thus challenges the field to expand the measurement of interoceptive performance into ecological, longitudinal, organ specific approaches that can be measured in concurrence with causal perturbations of arousal during clinically relevant states.

In conclusion, this study is evidence that the objective ability to detect cardiac and respiratory signals at physiological rest may not be a meaningful transdiagnostic marker of general psychopathology. In a well-powered, multi-domain psychophysical design, we found minimal reliable links between cardiac or respiratory interoceptive perception and latent psychiatric dimensions spanning stress, anxiety, depression, ADHD, and autism spectrum. In contrast, self-reported bodily attention and beliefs showed moderate associations, likely reflecting subjective appraisal or affective responses rather than objective insight into bodily signals. Specifically, these associations indicate that subjective interoception may primarily reflect clinically relevant higher-order beliefs concerning bodily mistrust, hypervigilance, and difficulty regulating attention toward internal sensations, rather than subjective sensory experience alone. Future work should examine diverse interoceptive modalities, clinical cohorts, and arousal or emotional manipulations to clarify when, and in whom, altered interoception contributes to general psychopathology.

## Methods and Materials

### Participants

We recruited participants as part of the Visceral Mind Project, a large-scale brain imaging project at the Centre of Functionally Integrative Neuroscience, Aarhus University. A total of 547 participants provided complete survey data (350 females, 196 males, 1 other gender, median age = 24, age range = 18-52). After the removal of multivariate survey outliers, a total of 500 participants were included in the multi-level exploratory factor analysis (313 females, 186 males, 1 other gender, median age = 24, age range = 18-52). From the interoceptive psychophysical test battery, 544 individuals completed the heart rate discrimination task (HRDT), and 315 completed the respiratory resistance sensitivity task (RRST). Due to a technical error, 31 participants were excluded from the HRDT dataset, yielding a final sample of 513 (324 females, 188 males, 1 other gender, median age = 24, age range = 18-56). Additionally, 48 participants were excluded from the RRST dataset due to suspected equipment malfunction and/or task noncompliance indicated by unusually low discrimination accuracy (less than 1.5 times interquartile range below the 25th percentile), resulting in a final RRST sample of 267 (184 females, 83 males, median age = 24, age range = 18-52). A total of 456 participants had both mental health factor scores and HRDT data (283 females, 172 males, 1 other gender, median age = 24, age range = 18-52), whereas 245 had both mental health factor scores and RRST data (167 females, 78 males, median age = 24, age range = 18-52).

As our aim was to assess a continuum of mental health symptoms, we adopted a participant recruitment strategy that sought to maximise between-subject symptom variance from fully healthy to subclinical to clinically relevant psychiatric conditions. Accordingly, we recruited participants from a wide range of possible online communities and backgrounds. We recruited participants in two data collection cohorts by advertising on nationwide participant pools, social media, newspapers, and posted fliers. We did not explicitly exclude participants with psychiatric diagnosis, and included participants did not report any major physical illnesses, or medication beyond over-the-counter antihistamines or contraceptives, furthermore they reported abstinence from alcohol/drugs 48 hours before participation. Symptom variance for psychiatric scores spanned from completely healthy individuals to those experiencing clinically relevant mental health burden, such that 47.2% of the sample exhibited medium or high levels of psychosocial stress, 28.6% mild depressive symptoms, 20.2% exhibited clinically significant levels of trait anxiety, 20.4% social anxiety, 17% ADHD symptoms, 15.8% medium or more severe somatic symptoms, 9.6% moderate or more severe depressive symptoms, and 8.6% autism spectrum symptoms (see Supplementary Table 7-8 for all percentage cut-offs).

As additional criteria, participants had normal or corrected-to-normal vision and were fluent in Danish or English. Furthermore, we only included participants compatible with MRI scanning (not pregnant or breastfeeding, no metal implants, claustrophobia, etc). Participants took part in multiple sessions including fMRI scans, behavioural tasks, physiological recordings, and psychiatric/lifestyle inventories. In this article, we focus on the psychiatric assessment battery and interoception tasks to evaluate the relationship of cardiac and respiratory interoceptive performance with mental health. All participants received remuneration for their participation. The local Region Midtjylland Ethics Committee granted approval for the study and all participants provided informed consent. The study was conducted in accordance with the Declaration of Helsinki (2013).

### Interoceptive Psychophysics Test Battery

To quantify interoceptive performance in the cardiac and respiratory domains, we employed a battery of validated psychophysical measures, including the Heart Rate Discrimination (HRDT) task^55^ and the Respiratory Resistance Sensitivity Task (RRST)^56^ (Figure 1C and 1D). These tasks used adaptive Bayesian methods to estimate key psychophysical parameters, including interoceptive sensitivity (threshold) and precision (slope)^57^. Participants completed 80 trials (cohort 1) or 60 trials (cohort 2) of the HRDT and 120 trials of the RRST. Interoceptive task order was counterbalanced between participants.

Both tasks featured a two-interval forced-choice design and were carefully designed to minimise non-interoceptive cues (e.g., auditory or visual feedback) that might influence performance. Custom-designed setups ensured robust and reproducible delivery of sensory stimuli, with inter-trial intervals and task durations optimised to maintain participant engagement and minimise fatigue. The adaptive Bayesian approaches used in both tasks provide efficient and reliable estimates of key psychophysical parameters while reducing the number of trials needed compared to traditional staircase methods.

#### Heart Rate Discrimination Task (HRDT)

In the HRDT^55^, participants were instructed to silently focus on their own heart rate without physically checking their pulse during a 5 second ‘heart listening’ period (Figure 1C). Following this, participants listened to a sequence of tones at varying tempos, presented at a fixed frequency above or below their actual heart rate, and indicated whether the feedback tones were faster or slower than their perceived heart rate using the left and right computer mouse buttons. After each trial, participants rated their confidence in their judgement accuracy using a visual analogue scale (VAS) ranging from ‘Guess’ to ‘Certain’. Throughout the task, heart rate was continuously monitored by a Nonin soft-clip pulse oximeter attached on the fourth finger of the non-dominant hand. The auditory feedback tones were dynamically adjusted using the Psi adaptive algorithm^75^, which efficiently estimated the threshold (absolute threshold being cardiac sensitivity) and slope (inverted slope being cardiac precision) of the psychometric function, expressed in units of delta-beats per minute (Δ-BPM).

#### Respiratory Resistance Sensitivity Task (RRST)

The RRST^56^ is a psychophysical paradigm in which participants took two successive inhalations through a computer-controlled respiratory circuit (Figure 1D). On each trial, one of the two breaths was randomly selected to receive a resistive load applied by a precision step motor compressing the circuit tubing, while the other breath remained without resistive load. The magnitude of the resistance was controlled using an adaptive psychophysical staircase method, allowing for the estimation of both threshold and slope. After completing the two inhalations, participants indicated whether the first or second breath was more difficult using the left and right computer mouse buttons. Participants then rated their confidence in their judgement accuracy using a VAS ranging from ‘Guess’ to ‘Certain’. To eliminate auditory cues from the motorised apparatus, participants wore headphones playing continuous white noise for noise-cancelling. Resistance levels were modulated across trials using the Psi-adaptive algorithm, enabling efficient estimation of the psychometric threshold (sensitivity to breathing resistance) and slope (respiratory precision).

### Hierarchical Bayesian Modelling of Interoceptive Psychophysics

We used Bayesian hierarchical psychometric function models to simultaneously estimate group and single-subject level parameters, allowing for a parsimonious model accounting for the structure of the data^57,76,77^.

For both conditions of the HRDT, we modelled the binary decision (i.e., “is the tone faster or slower than your heart rate?”) as a function of the difference between the frequency of auditory tones and the participant’s heart rate (cardiac condition: HRDT-Interoception) or reference tones (auditory condition: HRDT-Exteroception) in beats per minute (BPM) using a cumulative normal distribution with three subject-specific parameters: threshold (α, sensitivity via absolute threshold), slope (β, precision via inverted slope) and lapse rate (λ). The psychophysical model is therefore as follows:

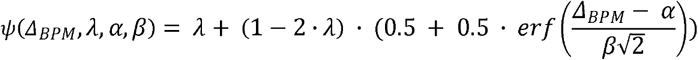

For the RRST, we applied a cumulative Gumbel function and modelled the accuracy of the binary decision (“was the first or second breath more difficult?”) as a function of the absolute difference between the resistive loads for the two breaths, estimating the same three subject-specific parameters as with the HRDT. The respiroceptive psychophysical model is as follows, where Res denotes the resistive load:

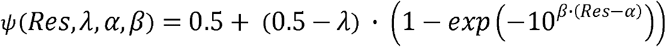

To evaluate these models, we used Stan, a No-U-Turn Hamiltonian Monte Carlo Markov Chain Monte Carlo sampler^78^. Each model was sampled using four chains each comprising 5000 iterations (2500 warm-up/2500 sampling), a target average acceptance probability (adapt_delta) of 0.99 and a maximum tree depth of 12. Each model was investigated for convergence and proper sampling by ensuring no divergences, satisfactory chain mixing, R_hat values less than 1.05^79^, and at least 400 effective samples for group-level parameters. For each model we selected a set of priors that contained a feasible range of behavioural responses. Subject-level parameters for threshold and slope were extracted for all tasks for further statistical analysis.

### Interoceptive Psychophysics Metrics

To assess sensitivity in the cardiac HRDT, we calculated the absolute threshold, which reflects the overall deviation from the reference beats without distinguishing between over- and underestimation of heart rate. A higher cardiac absolute threshold therefore indicates lower sensitivity: a greater discrepancy between the perceived and actual heart rate. Additionally, to maintain consistency in interpretation across measures, we inverted the scale of the cardiac slope values so that higher slopes uniformly indicate greater precision, as in the respiratory slope used in the RRST. For the RRST, a higher threshold indicates a greater respiratory resistance was needed to achieve an 82% probability of a correct response, thus lower sensitivity.

We analysed metacognition for the interoceptive tasks using M-Ratio (meta-d’/d’), a Bayesian measure of metacognitive efficiency (adjustment of confidence with performance) that controls first-order discrimination performance and variability in confidence ratings^58,80^. The model was fitted to each subject separately using bespoke functions from the HMeta-d toolbox in R, including Markov chain Monte Carlo sampling in JAGS^58^. As M-Ratio values can be unstable when task performance is low or when confidence ratings are extreme^81^, we excluded individuals based on two criteria: those with either negative or extreme M-Ratio values (more than three times the median absolute deviation from the median), as these are psychophysically implausible and imply either poor task adherence or model estimation problems. This resulted in 10 participants being excluded from metacognitive analyses for the respiratory task, and 96 excluded for cardiac metacognition (see Supplementary Table 1 for descriptive statistics of all task metrics).

Therefore, for each cardiac and respiratory task we obtained the following interception metrics: sensitivity (threshold), precision (slope), metacognitive bias (mean confidence), and metacognitive efficiency (M-Ratio: meta-d’/d’).

### Interoceptive Sensibility (MAIA)

The 32-item, self-administered Multidimensional Assessment of Interoceptive Awareness (MAIA)^82^ questionnaire measures interoceptive sensibility, defined as “the subjective perception and beliefs about an individual’s internal focus and/or accuracy in perceiving interoceptive signals”^83^. Participants rated their responses on a scale from 0 (Never) to 5 (Always). Its eight subscales include: Noticing, Not-Distracting, Not-Worrying, Attention Regulation, Emotional Awareness, Self-Regulation, Body Listening, and Trusting. These subscale dimensions were calculated as mean scores, with items 5-9 reverse coded following the published guidelines (see Supplementary Table 15 for MAIA individual item details, and Supplementary Table 2 for subscale descriptive statistics).

### Psychiatric Symptom Assessment Battery

Participants completed a battery of psychiatric and lifestyle assessments. To ensure an adequate sample-to-variable ratio for factor analysis, we focused our analysis on eight survey instruments assessing mental health spanning autism spectrum (AQ10), ADHD (ASRS), depression (MDI and PHQ9), somatic symptoms (PHQ15), stress (PSS), social anxiety (SIAS), and trait anxiety (STAI trait) (see Supplementary Table 7 for survey abbreviations and details). Together, these surveys encompassed 116 items. All scales utilised validated Danish translations, except in cases where participants spoke English as their first language, in which case validated English versions were used.

### Statistical Analyses

#### Survey data handling and multivariate outliers

Two survey items were excluded due to excessive missing values: item 10 of PHQ9 (additional follow-up question) and item 4 of PHQ15 (for women only) (see Supplementary Table 16). We detected multivariate outliers via Mahalanobis Distance, with outliers defined as larger than the critical chi-squared value for the number of variables = 114, at an alpha = 0.001. This identified 47 multivariate outliers, resulting in a final survey sample of 500 participants.

#### Mental Health Exploratory Factor Analysis

First, we inspected the correlation and sampling adequacy of the survey responses for factor analysis via Bartlett’s Test of Sphericity^84^ and Kaiser-Meyer-Olkin’s statistic^85^. Thus, we excluded Item 9 of PHQ15 as the Kaiser-Meyer-Olkin’s statistic was < 0.5. Thus, 113 final items were included in the Exploratory Factor Analysis (see Supplementary Table 16). The Bartlett’s Test of Sphericity revealed the 113 mental health responses were not random (X^2^(6328) = 28393.24, p < .001) and the Kaiser-Meyer-Olkin statistic indicated the items were factorable (MSA = 0.921: indicated as ‘marvellous’).

We conducted an item-level exploratory factor analysis using a weighted least square (WLS) estimation method, and an oblique (oblimin) rotation as we analysed ordinal survey data which are expected to correlate^86,87^. To identify the number of factors, we visually inspected the inflexion point of the scree plot and completed parallel analysis. We then assessed the interpretability of the factors and fit indices (Tucker-Lewis index (TLI), comparative fit index (CFI), and root mean square error of approximation (RMSEA)) for the factor solution^88^. In addition, we aimed to achieve a very simple structure (VSS) by analysing the factor solution after removing items which cross-loaded on multiple factors (r > 0.3) or did not load any factors (r > 0.3), resulting in 80 items in the final exploratory factor analysis^61^. Finally, we computed reliability estimates of the factors (Cronbach’s alpha and Omega coefficients), and factor scores for each participant via the Bartlett method. The EFA analyses were conducted using the *‘psych’* R statistical package.

As we identified an 11 factor solution, we performed a multi-level hierarchical factor analysis to further reduce the dimensions of the mental health scores using the R ‘*fa*.*multi*’ function. The number of factors for the 2nd level EFA was estimated using the factor scores from the 1st level EFA.

#### Cross-Correlation and Bayes Factor Analyses

We used Spearman’s rank correlation coefficients to test the association between mental health and interoception across cardiac and respiratory domains. Mental health included the 11 factors from the exploratory factor analysis and the two higher-level factors encompassing the 11 lower-level factors (higher-level factors: overall negative mental health (encompasses the first level factors: social anxiety, self-confidence, sleep, depressive symptoms, stress, autism spectrum), and ADHD & somatic symptoms (first level factors: negative thoughts, restlessness, somatic symptoms, impulsivity), with the first level factor ‘inattentiveness’ not strongly loading onto either higher-level factor). Interoceptive performance metrics included hierarchically estimated measures of interoceptive sensitivity (threshold) and interoceptive precision (slope) from cardiac (HRDT) and respiratory (RRST) psychophysical tasks, as well as metacognitive bias (mean confidence on the tasks) and metacognitive efficiency (M-Ratio). We also completed a Spearman cross-correlation matrix between mental health and self-reported interoceptive sensibility measured via the eight MAIA subscales, as well as a Spearman cross-correlation matrix between the MAIA subscales and the interoceptive performance metrics described above (threshold, slope, mean confidence, and M-Ratio).

Correlation matrices’ p-values were corrected for multiple comparisons using the false discovery rate (FDR) correction method (Benjamini-Hochberg procedure at < 0.05). All correlations were also repeated via partial correlations which controlled for age, gender, and body mass index. Finally, we computed Bayes Factors to quantify the evidence in favour of the null hypothesis (BF01 = no association) for each correlation pair, with default priors applied (‘rscale=medium’ for correlationBF function in R). We performed Bayesian hypothesis testing using the ‘BayesFactor’ package^89^. These Bayes Factors were presented in a correlation matrix for visualisation.

#### Clinical severity analyses

To examine whether interoceptive–mental health associations emerged selectively at higher levels of symptom severity, we conducted complementary clinical cut-off group comparisons and upper symptom percentile-based correlation analyses. For the cut-off group analyses, participants survey scores were stratified according to commonly used clinical screening thresholds for each mental health questionnaire (see Supplementary Table 7-9 for cut-off definitions and sample sizes). Interoceptive psychophysics measures (cardiac and respiratory sensitivity, precision, confidence, and metacognition) were compared between participants above versus below each threshold using two-sided Mann–Whitney U tests, with false discovery rate (FDR) correction applied to control for multiple comparisons.

In parallel, percentile-based threshold analyses were conducted by restricting symptom score distributions to progressively higher severity ranges (top 50%, 75%, and 90% of the sample). Within each restricted subsample, Spearman rank correlations were computed between interoceptive measures and symptom scores. Multiple-comparison correction was applied within each percentile-specific correlation matrix using FDR correction. These analyses were designed to test whether interoceptive–symptom relationships strengthened or emerged selectively at higher symptom severity levels, independent of linear associations observed across the full sample.

### Mental health network analysis

To investigate an alternative representation of psychopathology, we conducted a mental health network analysis. To this end, we applied the ‘bootnet’ R package to represent the 114 mental health survey items as nodes, with edges signifying partial correlation coefficients^90^. We employed the EBICglasso method for network estimation, which incorporates a graphical least absolute shrinkage and selection operator (glasso) that shrinks edge weights towards zero, fostering a sparse network and reducing spurious edges. An Extended Bayesian Information Criterion (EBIC) is used to set a regularisation tuning parameter for estimating a weighted network. Community structure of clustered psychiatric symptoms was identified using the spin-glass algorithm.

#### Semantic similarity analysis of MAIA

To examine the construct validity of the interoceptive sensibility scale, we conducted a semantic similarity analysis of the Multidimensional Assessment of Interoceptive Awareness (MAIA) questionnaire items in relation to other psychological scales. We used the Semantic Scale Network^64^, an online tool that employs semantic similarity analysis to estimate the similarity between survey items based on large-scale language corpora of psychological scales. This tool applies standard text preprocessing, including lowercasing, removing punctuation, stop words, and numbers, as well as lemmatization and stemming. It compares psychological instruments based on word matching, word importance (rarity within the corpus), and latent semantic analysis, which captures deeper similarities in meaning between non-identical terms. The MAIA items were entered as the target scale, and its semantic similarity was computed against a database of over 4000 psychological instruments. We recorded the top 15 scales with the highest similarity scores (cosine similarity > 0.4). In addition, semantic similarity analyses were repeated separately for each of the eight MAIA subscales, using the corresponding item sets as target scales.

## Acknowledgements

This research is financially supported by a Lundbeckfonden Fellowship R272-2017-4345 (MA, LB, NN), a European Research Council Grant ERC-2020-StG-948788 (MA, LB, AT, MV), a Lundbeckfonden Experiment Grant R436-2023-991 (FF, JFE, RB), a European Research Council Grant (ERC-2020-StG-948838) (FF, AC), a Lundbeck Foundation grant R389-2021-1596 Neuroscience Academy Denmark (KH), and a Swiss National Science Foundation Postdoc.Mobility Nr. 217637 (FB). The funding source had no involvement in the study design, collection, analysis, interpretation, or writing of the manuscript.

## Author contributions

LB and MA analysed the data, interpreted the results, and wrote the manuscript, MA also provided supervision, NN provided conceptual advice, collected data, and preprocessed the RRST data, JFE and ASC completed computational modelling of psychophysical tasks, and provided conceptual advice. MV and RB collected data and provided conceptual advice. AT, FB, KH, RB, and FF provided conceptual advice and edited the manuscript. All authors read and approved the manuscript.

## Conflict of interest

All authors declare no conflicts of interest.

## Data availability

Deidentified participant data (interoception task and mental health factor analysis) and scripts implemented in this paper are available here: https://github.com/embodied-computation-group/InteroMentalHealth Due to Danish privacy law, raw mental health data are available upon reasonable request, with the formation of a data sharing agreement. Researchers who wish to access these data may contact M.A. at The Center of Functionally Integrative Neuroscience, Aarhus University, Denmark.

## Notes

### Competing Interest Statement

The authors have declared no competing interest.

### Author Declarations

We recruited participants as part of the Visceral Mind Project, a large-scale brain imaging project at the Centre of Functionally Integrative Neuroscience, Aarhus University. The local Region Midtjylland Ethics Committee granted approval for the study and all participants provided informed consent. The study was conducted in accordance with the Declaration of Helsinki (2013).

### Summary of Updates

Revisions from peer review - including additional non-linear clinical severity analyses and network analyses.

