## Supplementary Material for "Interoceptive Performance is Unrelated to Mental Health Symptoms: Evidence From a Large Scale Multi-Domain Psychophysical Investigation"

**Supplementary Table 1: Descriptive Statistics.** Descriptive statistics of the performance metrics on the cardiac (HRDT) and respiratory (RRST) interoception tasks, and the multi-level mental health factor analysis scores. Absolute values are given for cardiac threshold (higher threshold = lower sensitivity), and cardiac precision has been inverted to be in the same direction as respiratory slope (higher slope = higher precision).

| Variable | N | Mean | Standard Deviation | 95% CI |
|---|---|---|---|---|
| RRST Conf | 267 | 60.199 | 13.125 | 58.617, 61.78 |
| RRST Slope | 267 | -1.306 | 0.517 | -1.368, -1.244 |
| RRST Thresh | 267 | 13.092 | 1.175 | 12.951, 13.234 |
| RRST MRatio | 257 | 0.880 | 0.236 | 0.851, 0.909 |
| HRDT Conf | 513 | 54.284 | 15.740 | 52.918, 55.649 |
| HRDT Slope | 513 | -9.975 | 1.914 | -10.141, -9.809 |
| HRDT Thresh | 513 | 11.063 | 8.802 | 10.3, 11.827 |
| HRDT MRatio | 417 | 0.711 | 0.378 | 0.675, 0.748 |
| Affective Symptoms | 500 | 0.000 | 1.126 | -0.099, 0.099 |
| ADHD & Somatic | 500 | 0.000 | 1.194 | -0.105, 0.105 |
| Social Anxiety | 500 | 0.000 | 1.031 | -0.091, 0.091 |
| Self-Confidence | 500 | 0.000 | 1.051 | -0.092, 0.092 |
| Sleep | 500 | 0.000 | 1.056 | -0.093, 0.093 |
| Inattentiveness | 500 | 0.000 | 1.110 | -0.098, 0.098 |
| Depression | 500 | 0.000 | 1.129 | -0.099, 0.099 |
| Negative Thoughts | 500 | 0.000 | 1.124 | -0.099, 0.099 |
| Restlessness | 500 | 0.000 | 1.134 | -0.1, 0.1 |
| Stress | 500 | 0.000 | 1.138 | -0.1, 0.1 |
| Autism Spectrum | 500 | 0.000 | 1.116 | -0.098, 0.098 |
| Somatic | 500 | 0.000 | 1.133 | -0.1, 0.1 |
| Impulsivity | 500 | 0.000 | 1.159 | -0.102, 0.102 |

**Supplementary Table 2: Descriptive Statistics for interoceptive sensibility (MAIA).** Descriptive statistics for interoceptive sensibility: eight subscales from Multidimensional Assessment of Interoceptive Awareness (MAIA) survey[1].

| MAIA Subscale | N | Mean | Standard Deviation | 95% CI |
|---|---|---|---|---|
| Noticing | 551 | 2.758 | 1.143 | 2.663, 2.854 |
| Not-Distracting | 551 | 1.331 | 0.523 | 1.287, 1.375 |
| Not-Worrying | 551 | 1.642 | 0.619 | 1.59, 1.694 |
| Attention Regulation | 551 | 2.605 | 0.985 | 2.523, 2.688 |
| Emotional Awareness | 551 | 2.850 | 1.126 | 2.756, 2.944 |
| Self-Regulation | 551 | 2.247 | 1.203 | 2.147, 2.348 |
| Body Listening | 551 | 2.079 | 1.263 | 1.973, 2.184 |
| Trusting | 551 | 3.572 | 1.129 | 3.478, 3.667 |

**Supplementary Table 3**: Hierarchical 11 factor lower-level Exploratory Factor Analysis results of mental health survey items including pattern coefficients for each of the factors (bold represents items > absolute 0.3), communality (commun), uniqueness (unique) and complexity (complex) estimates. This included the following factors: social anxiety, self-confidence, sleep, inattentiveness, depression, negative thoughts, restlessness, stress, autism spectrum, somatic symptoms, and impulsivity.

| | Social Anxiety | *Self.Conf* | *Sleep* | *Inattent* | *Depress* | *Neg Thought* | *Restless* | *Stress* | *Autism* | *Somatic* | *Implusiv* | Commun | Unique | Complex |
|---|---|---|---|---|---|---|---|---|---|---|---|---|---|---|
| **sias_16** | 0.823 | *-0.038* | *0.034* | *-0.017* | *-0.023* | *-0.017* | *-0.042* | *-0.002* | *0.017* | *0.038* | *0.063* | 0.71 | 0.29 | 1.03 |
| **sias_19** | 0.810 | *0.007* | *0.037* | *-0.034* | *-0.007* | *0.090* | *0.007* | *0.041* | *-0.017* | *0.005* | *-0.006* | 0.71 | 0.29 | 1.04 |
| **sias_7** | 0.799 | *-0.072* | *0.009* | *-0.026* | *-0.039* | *-0.071* | *-0.040* | *0.117* | *0.030* | *-0.041* | *0.032* | 0.68 | 0.32 | 1.10 |
| **sias_15** | 0.777 | *0.044* | *-0.004* | *0.041* | *0.057* | *0.099* | *0.042* | *-0.070* | *0.047* | *0.081* | *-0.032* | 0.71 | 0.29 | 1.11 |
| **sias_10** | 0.755 | *-0.052* | *0.050* | *0.072* | *0.010* | *-0.179* | *0.088* | *0.005* | *0.043* | *-0.057* | *-0.142* | 0.66 | 0.34 | 1.28 |
| **sias_6** | 0.608 | *0.046* | *-0.059* | *0.060* | *0.056* | *0.037* | *-0.030* | *0.057* | *-0.020* | *0.067* | *0.072* | 0.42 | 0.58 | 1.16 |
| **sias_8** | 0.595 | *0.108* | *0.028* | *0.028* | *0.022* | *-0.019* | *0.053* | *0.127* | *0.012* | *-0.051* | *-0.027* | 0.37 | 0.63 | 1.21 |
| **sias_9** | -0.592 | *0.156* | *0.009* | *-0.015* | *0.086* | *0.025* | *0.061* | *-0.056* | *-0.085* | *0.004* | *0.022* | 0.45 | 0.55 | 1.28 |
| **sias_17** | 0.585 | *-0.110* | *-0.030* | *-0.028* | *-0.010* | *0.138* | *0.074* | *0.029* | *-0.028* | *-0.052* | *0.113* | 0.51 | 0.49 | 1.34 |
| **sias_20** | 0.577 | *0.185* | *-0.007* | *0.139* | *0.125* | *0.137* | *-0.025* | *-0.070* | *-0.020* | *0.065* | *0.090* | 0.44 | 0.56 | 1.71 |
| **sias_12** | 0.557 | *0.007* | *0.023* | *-0.076* | *0.093* | *0.196* | *0.024* | *-0.001* | *-0.057* | *-0.026* | *0.065* | 0.44 | 0.56 | 1.42 |
| **sias_18** | 0.506 | *-0.090* | *0.020* | *-0.008* | *0.027* | *0.179* | *0.017* | *-0.086* | *-0.060* | *0.065* | *0.092* | 0.42 | 0.58 | 1.55 |
| **sias_4** | 0.467 | *-0.159* | *-0.002* | *0.067* | *-0.010* | *0.013* | *0.081* | *-0.018* | *0.098* | *0.001* | *-0.051* | 0.39 | 0.61 | 1.48 |
| **sias_11** | -0.454 | *0.063* | *-0.004* | *-0.156* | *-0.051* | *0.053* | *0.025* | *0.079* | *-0.179* | *-0.034* | *0.197* | 0.39 | 0.61 | 2.23 |
| **sias_14** | 0.442 | *0.069* | *0.028* | *-0.009* | *0.198* | *0.069* | *-0.046* | *-0.132* | *0.008* | *0.035* | *0.005* | 0.27 | 0.73 | 1.77 |
| **sias_2** | 0.410 | *-0.088* | *0.071* | *-0.010* | *0.078* | *-0.111* | *0.142* | *-0.052* | *0.081* | *-0.021* | *-0.006* | 0.29 | 0.71 | 1.81 |
| **sias_5** | -0.382 | *0.227* | *0.022* | *-0.042* | *0.104* | *-0.015* | *-0.004* | *0.063* | *-0.135* | *-0.098* | *0.103* | 0.31 | 0.69 | 2.61 |
| **stai_36** | *0.038* | 0.758 | *-0.025* | *0.041* | *-0.080* | *-0.038* | *-0.002* | *-0.063* | *-0.019* | *-0.050* | *-0.020* | 0.70 | 0.30 | 1.07 |
| **stai_23** | *-0.047* | 0.737 | *0.022* | *-0.032* | *-0.085* | *-0.022* | *0.030* | *0.059* | *-0.076* | *-0.052* | *-0.062* | 0.65 | 0.35 | 1.11 |
| **stai_30** | *-0.097* | 0.698 | *-0.086* | *-0.005* | *-0.062* | *0.079* | *-0.022* | *-0.072* | *-0.021* | *0.049* | *0.034* | 0.66 | 0.34 | 1.15 |
| **stai_21** | *-0.013* | 0.680 | *-0.060* | *-0.053* | *-0.043* | *0.022* | *-0.040* | *-0.145* | *0.022* | *0.020* | *0.034* | 0.65 | 0.35 | 1.15 |
| **stai_33** | *-0.071* | 0.678 | *-0.022* | *-0.002* | *0.054* | *0.003* | *-0.066* | *0.014* | *-0.005* | *-0.016* | *0.017* | 0.51 | 0.49 | 1.06 |
| **stai_39** | *-0.036* | 0.565 | *-0.053* | *-0.137* | *0.008* | *-0.138* | *0.017* | *-0.025* | *-0.071* | *-0.016* | *-0.020* | 0.54 | 0.46 | 1.32 |
| **stai_27** | *0.086* | 0.509 | *-0.118* | *-0.041* | *0.119* | *-0.094* | *-0.158* | *-0.161* | *0.036* | *-0.076* | *-0.012* | 0.53 | 0.47 | 1.91 |
| **pss_4** | *0.020* | 0.351 | *-0.010* | *-0.072* | *-0.213* | *-0.136* | *0.019* | *-0.087* | *-0.054* | *-0.017* | *-0.006* | 0.37 | 0.63 | 2.35 |
| **stai_32** | *0.284* | -0.351 | *-0.016* | *-0.034* | *0.166* | *0.192* | *0.001* | *-0.085* | *0.042* | *0.032* | *0.061* | 0.49 | 0.51 | 3.37 |
| **stai_35** | *0.103* | -0.344 | *-0.027* | *0.186* | *0.223* | *0.210* | *-0.065* | *0.036* | *0.003* | *0.013* | *0.043* | 0.49 | 0.51 | 3.56 |
| **stai_24** | *0.185* | -0.312 | *0.119* | *-0.055* | *0.163* | *0.170* | *0.037* | *-0.078* | *0.011* | *-0.027* | *0.078* | 0.40 | 0.60 | 3.83 |
| **pss_5** | *0.038* | 0.311 | *-0.080* | *-0.044* | *-0.281* | *-0.038* | *0.030* | *-0.173* | *-0.071* | *-0.019* | *0.034* | 0.40 | 0.60 | 3.04 |
| **phq_9_3** | *0.014* | *0.040* | 0.888 | *-0.026* | *-0.037* | *0.025* | *-0.004* | *0.011* | *0.045* | *-0.039* | *0.016* | 0.76 | 0.24 | 1.02 |
| **phq15_14** | *-0.026* | *-0.003* | 0.883 | *-0.004* | *-0.026* | *0.020* | *-0.034* | *-0.054* | *-0.026* | *-0.043* | *-0.003* | 0.71 | 0.29 | 1.02 |
| **phq15_15** | *0.059* | *-0.064* | 0.601 | *0.046* | *0.105* | *-0.060* | *0.011* | *0.031* | *-0.016* | *0.189* | *0.012* | 0.58 | 0.42 | 1.35 |
| **mdi_9a** | *-0.061* | *-0.009* | 0.551 | *0.052* | *0.031* | *-0.037* | *0.060* | *0.050* | *-0.033* | *-0.044* | *-0.011* | 0.34 | 0.66 | 1.12 |
| **phq_9_4** | *0.049* | *-0.099* | 0.536 | *0.050* | *0.114* | *0.010* | *0.086* | *-0.003* | *-0.016* | *0.181* | *0.012* | 0.58 | 0.42 | 1.50 |
| **asrs_a_4** | *0.060* | *0.053* | *0.051* | 0.623 | *0.038* | *0.018* | *0.057* | *0.108* | *-0.021* | *0.032* | *-0.052* | 0.49 | 0.51 | 1.16 |
| **asrs_a_2** | *0.064* | *-0.119* | *0.034* | 0.613 | *-0.100* | *-0.056* | *-0.035* | *0.154* | *0.060* | *-0.036* | *0.045* | 0.51 | 0.49 | 1.36 |
| **asrs_b_2** | *-0.037* | *-0.061* | *0.036* | 0.523 | *0.024* | *0.069* | *0.176* | *-0.189* | *-0.045* | *0.074* | *0.015* | 0.40 | 0.60 | 1.67 |
| **asrs_a_1** | *0.066* | *0.044* | *0.041* | 0.506 | *0.016* | *-0.071* | *0.074* | *0.180* | *0.017* | *-0.105* | *0.088* | 0.40 | 0.60 | 1.59 |
| **asrs_a_3** | *-0.047* | *0.012* | *0.080* | 0.479 | *0.133* | *0.020* | *0.015* | *-0.004* | *0.035* | *0.037* | *0.052* | 0.32 | 0.68 | 1.30 |
| **asrs_b_1** | *-0.030* | *0.078* | *-0.022* | 0.430 | *0.096* | *0.055* | *0.122* | *-0.025* | *0.045* | *-0.067* | *0.125* | 0.29 | 0.71 | 1.68 |
| **asrs_b_5** | *0.032* | *-0.143* | *-0.018* | 0.375 | *-0.047* | *0.147* | *0.016* | *-0.134* | *-0.023* | *0.023* | *0.164* | 0.28 | 0.72 | 2.49 |
| **aq10_4** | *-0.088* | *-0.154* | *0.060* | 0.371 | *-0.045* | *0.150* | *-0.005* | *0.015* | *0.141* | *-0.058* | *0.022* | 0.28 | 0.72 | 2.38 |

| | | | | | | | | | | | | | | |
|---|---|---|---|---|---|---|---|---|---|---|---|---|---|---|
| **asrs_b_4** | -0.009 | -0.009 | 0.083 | 0.342 | 0.121 | 0.132 | 0.024 | -0.087 | -0.110 | 0.010 | 0.216 | 0.31 | 0.69 | 2.99 |
| **aq10_3** | 0.055 | -0.035 | -0.004 | 0.324 | -0.083 | 0.032 | -0.086 | 0.020 | 0.130 | -0.044 | 0.012 | 0.15 | 0.85 | 1.82 |
| **asrs_b_3** | 0.102 | -0.075 | 0.003 | 0.310 | -0.043 | 0.085 | 0.195 | 0.000 | -0.015 | 0.034 | 0.083 | 0.30 | 0.70 | 2.56 |
| **mdi_1** | -0.001 | -0.153 | 0.037 | -0.062 | 0.564 | 0.140 | 0.018 | 0.140 | 0.008 | 0.021 | 0.061 | 0.59 | 0.41 | 1.49 |
| **mdi_2** | 0.051 | 0.024 | 0.220 | 0.139 | 0.481 | -0.093 | 0.055 | 0.099 | 0.081 | 0.117 | -0.036 | 0.50 | 0.50 | 2.10 |
| **mdi_5** | 0.030 | -0.118 | 0.008 | 0.134 | 0.453 | 0.048 | 0.128 | 0.124 | -0.056 | 0.000 | 0.027 | 0.48 | 0.52 | 1.77 |
| **mdi_4** | 0.077 | -0.267 | 0.014 | 0.006 | 0.445 | 0.034 | 0.160 | 0.022 | 0.026 | 0.024 | 0.039 | 0.56 | 0.44 | 2.07 |
| **mdi_8b** | 0.079 | -0.069 | 0.149 | 0.048 | 0.436 | 0.026 | 0.095 | -0.072 | -0.011 | -0.019 | 0.032 | 0.36 | 0.64 | 1.59 |
| **phq_9_2** | 0.081 | -0.218 | 0.102 | -0.048 | 0.401 | 0.052 | 0.084 | 0.192 | -0.006 | -0.073 | 0.034 | 0.54 | 0.46 | 2.65 |
| **mdi_6** | 0.136 | -0.279 | -0.044 | -0.064 | 0.339 | -0.072 | 0.206 | 0.037 | -0.038 | -0.127 | -0.024 | 0.38 | 0.62 | 3.72 |
| **stai_40** | -0.038 | 0.024 | 0.076 | 0.039 | 0.073 | 0.682 | 0.016 | 0.132 | 0.041 | 0.031 | -0.041 | 0.59 | 0.41 | 1.16 |
| **stai_29** | 0.130 | -0.012 | 0.001 | -0.012 | -0.007 | 0.594 | 0.122 | 0.063 | -0.019 | 0.039 | 0.024 | 0.53 | 0.47 | 1.23 |
| **stai_38** | 0.106 | -0.043 | 0.087 | 0.003 | 0.034 | 0.575 | -0.014 | 0.043 | 0.000 | -0.010 | 0.011 | 0.46 | 0.54 | 1.15 |
| **stai_37** | 0.001 | -0.100 | 0.030 | 0.167 | -0.152 | 0.424 | 0.161 | 0.066 | 0.054 | 0.056 | -0.019 | 0.40 | 0.60 | 2.25 |
| **mdi_8a** | -0.029 | 0.034 | 0.022 | -0.006 | 0.168 | -0.015 | 0.705 | 0.040 | 0.073 | 0.037 | -0.055 | 0.58 | 0.42 | 1.17 |
| **asrs_b_7** | 0.033 | 0.009 | 0.047 | 0.149 | -0.068 | 0.079 | 0.633 | -0.028 | -0.020 | 0.073 | 0.150 | 0.64 | 0.36 | 1.34 |
| **asrs_a_6** | -0.105 | 0.015 | -0.031 | 0.070 | -0.086 | -0.012 | 0.435 | 0.019 | -0.066 | 0.006 | 0.233 | 0.31 | 0.69 | 1.91 |
| **stai_22** | 0.077 | -0.220 | 0.062 | -0.060 | -0.146 | 0.264 | 0.405 | 0.077 | 0.028 | -0.048 | 0.045 | 0.49 | 0.51 | 3.10 |
| **phq_9_8** | 0.156 | -0.044 | 0.064 | -0.075 | 0.016 | -0.058 | 0.352 | 0.064 | -0.033 | -0.022 | 0.114 | 0.23 | 0.77 | 2.05 |
| **pss_10** | 0.006 | -0.094 | 0.024 | 0.122 | 0.141 | 0.100 | 0.049 | 0.615 | 0.006 | 0.012 | 0.078 | 0.71 | 0.29 | 1.35 |
| **pss_6** | 0.080 | -0.065 | 0.021 | 0.222 | 0.084 | 0.056 | 0.026 | 0.468 | 0.017 | 0.166 | -0.019 | 0.54 | 0.46 | 1.98 |
| **stai_28** | 0.127 | -0.042 | 0.066 | 0.105 | 0.089 | 0.197 | 0.059 | 0.445 | 0.039 | -0.009 | 0.020 | 0.54 | 0.46 | 1.95 |
| **pss_3** | 0.057 | -0.199 | 0.075 | -0.012 | -0.062 | 0.145 | 0.083 | 0.443 | -0.033 | 0.185 | 0.012 | 0.56 | 0.44 | 2.33 |
| **pss_1** | 0.051 | -0.077 | -0.004 | -0.135 | 0.013 | 0.197 | 0.048 | 0.428 | 0.054 | 0.083 | 0.068 | 0.39 | 0.61 | 1.99 |
| **aq10_9** | -0.002 | -0.013 | -0.057 | -0.040 | 0.091 | -0.020 | 0.041 | -0.056 | 0.714 | 0.027 | 0.024 | 0.51 | 0.49 | 1.08 |
| **aq10_5** | -0.031 | -0.046 | -0.008 | 0.119 | -0.041 | 0.042 | -0.043 | -0.072 | 0.694 | -0.042 | -0.014 | 0.52 | 0.48 | 1.13 |
| **aq10_10** | -0.031 | -0.010 | -0.070 | 0.049 | 0.061 | -0.019 | -0.073 | -0.053 | -0.670 | -0.011 | -0.057 | 0.49 | 0.51 | 1.11 |
| **aq10_6** | 0.036 | 0.066 | 0.017 | -0.070 | -0.025 | -0.016 | -0.058 | 0.105 | 0.628 | -0.006 | 0.011 | 0.40 | 0.60 | 1.14 |
| **aq10_7** | -0.007 | 0.077 | 0.027 | 0.026 | 0.014 | -0.028 | -0.015 | 0.012 | -0.346 | 0.084 | -0.018 | 0.14 | 0.86 | 1.28 |
| **phq15_13** | 0.014 | 0.007 | 0.035 | -0.024 | -0.061 | -0.059 | 0.048 | 0.015 | -0.019 | 0.723 | 0.027 | 0.53 | 0.47 | 1.05 |
| **phq15_12** | 0.009 | 0.006 | -0.034 | 0.012 | -0.047 | -0.009 | -0.052 | 0.026 | -0.007 | 0.677 | 0.020 | 0.44 | 0.56 | 1.03 |
| **phq15_1** | -0.003 | -0.007 | -0.038 | -0.030 | 0.071 | 0.033 | 0.022 | 0.001 | 0.018 | 0.660 | -0.025 | 0.45 | 0.55 | 1.05 |
| **phq15_6** | -0.022 | -0.072 | 0.206 | -0.016 | 0.033 | 0.074 | -0.026 | -0.013 | -0.040 | 0.337 | 0.073 | 0.24 | 0.76 | 2.10 |
| **phq15_8** | -0.052 | -0.027 | 0.002 | -0.083 | 0.055 | 0.006 | 0.156 | 0.091 | 0.024 | 0.288 | -0.040 | 0.15 | 0.85 | 2.26 |
| **asrs_b_12** | -0.021 | 0.117 | 0.013 | 0.065 | 0.092 | -0.053 | 0.074 | 0.021 | 0.140 | 0.007 | 0.628 | 0.46 | 0.54 | 1.29 |
| **asrs_b_11** | 0.040 | -0.011 | 0.059 | 0.062 | -0.073 | -0.003 | 0.031 | 0.009 | 0.060 | -0.025 | 0.600 | 0.41 | 0.59 | 1.11 |
| **asrs_b_9** | -0.086 | -0.044 | 0.038 | -0.111 | -0.019 | 0.009 | 0.017 | 0.015 | -0.035 | 0.133 | 0.596 | 0.41 | 0.59 | 1.25 |
| **asrs_b_10** | 0.113 | -0.087 | -0.047 | 0.080 | 0.015 | -0.113 | -0.017 | 0.024 | -0.081 | -0.031 | 0.546 | 0.33 | 0.67 | 1.36 |

**Supplementary Table 4**: Hierarchical 11 lower-level factors and two higher level-factors Exploratory Factor Analysis item details for each mental health factor (items > absolute 0.3 loading).

| Item | Question | Loading |
|---|---|---|
| | **Factor 1: Social Anxiety** | |
| sias_16 | I am nervous mixing with people I don't know well. | 0.823 |
| sias_19 | I am tense mixing in a group. | 0.810 |
| sias_7 | When mixing socially, I am uncomfortable. | 0.799 |
| sias_15 | I find myself worrying that I won't know what to say in social situations. | 0.777 |
| sias_10 | I have difficulty talking with other people. | 0.755 |
| sias_6 | I tense up if I meet an acquaintance in the street. | 0.608 |
| sias_8 | I feel tense if I am alone with just one other person. | 0.595 |
| sias_9 | I am at ease meeting people at parties, etc. | -0.592 |
| sias_17 | I feel I'll say something embarrassing when talking. | 0.585 |
| sias_20 | I am unsure whether to greet someone I know only slightly." | 0.577 |
| sias_12 | I worry about expressing myself in case I appear awkward. | 0.557 |
| sias_18 | When mixing in a group, I find myself worrying I will be ignored. | 0.506 |
| sias_4 | I find it difficult to mix comfortably with the people I work with. | 0.467 |
| sias_11 | I find it easy to think of things to talk about. | -0.454 |
| sias_14 | I have difficulty talking to attractive persons of the opposite sex. | 0.442 |
| sias_2 | I have difficulty making eye contact with others. | 0.410 |
| sias_5 | I find it easy to make friends my own age. | -0.382 |
| | **Factor 2: Self-Confidence** | |
| stai_36 | I am content | 0.758 |
| stai_23 | I feel satisfied with myself | 0.737 |
| stai_30 | I am happy | 0.698 |
| stai_21 | I feel pleasant | 0.680 |
| stai_33 | I feel secure | 0.678 |
| stai_39 | I am a steady person | 0.565 |
| stai_27 | I am 'calm, cool, and collected' | 0.509 |
| pss_4 | In the last month, how often have you felt confident about your ability to handle your personal problems? | 0.351 |
| stai_32 | I lack self-confidence | -0.351 |
| stai_35 | I feel inadequate | -0.344 |
| stai_24 | I wish I could be as happy as others seem to be | -0.312 |
| pss_5 | In the last month, how often have you felt that things were going your way? | 0.311 |
| | **Factor 3: Sleep** | |
| phq_9_3 | Trouble falling or staying asleep, or sleeping too much | 0.888 |
| phq15_14 | Trouble falling asleep, sleeping or sleeping too much | 0.883 |
| phq15_15 | Felt tired or only had little energy | 0.601 |
| mdi_9a | Have you been sleeping too little? | 0.551 |
| phq_9_4 | Feeling tired or having little energy | 0.536 |
| | **Factor 4: Inattentiveness** | |
| asrs_a_4 | When you have a task that requires a lot of thought, how often do you avoid or delay getting started? | 0.623 |
| asrs_a_2 | How often do you have difficulty getting things in order when you have to do a task that requires organization? | 0.613 |
| asrs_b_2 | How often do you have difficulty keeping your attention when you are doing boring or repetitive work? | 0.523 |
| asrs_a_1 | How often do you have trouble wrapping up the final details of a project, once the challenging parts have been done? | 0.506 |
| asrs_a_3 | How often do you have problems remembering appointments or obligations? | 0.479 |
| asrs_b_1 | How often do you make careless mistakes when you have to work on a boring or difficult project? | 0.430 |
| asrs_b_5 | How often are you distracted by activity or noise around you? | 0.375 |
| aq10_4 | If there is an interruption, I can switch back to what I was doing very quickly | 0.371 |
| asrs_b_4 | How often do you misplace or have difficulty finding things at home or at work? | 0.342 |
| aq10_3 | I find it easy to do more than one thing at once | 0.324 |

| | | |
|---|---|---|
| asrs_b_3 | How often do you have difficulty concentrating on what people say to you, even when they are speaking to you directly? | 0.310 |
| **Factor 5: Depression** | | |
| mdi_1 | Have you felt low in spirits or sad? | 0.564 |
| mdi_2 | Have you lost interest in your daily activities? | 0.481 |
| mdi_5 | Have you had a bad conscience or feelings of guilt? | 0.453 |
| mdi_4 | Have you felt less self-confident? | 0.445 |
| mdi_8b | Have you felt subdued or slowed down? | 0.436 |
| phq_9_2 | Feeling down, depressed, or hopeless | 0.401 |
| mdi_6 | Have you felt that life wasn't worth living? | 0.339 |
| **Factor 6: Negative Thoughts** | | |
| stai_40 | I get in a state of tension or turmoil as I think over my recent concerns and interests | 0.682 |
| stai_29 | I worry too much over something that really doesn't matter | 0.594 |
| stai_38 | I take disappointments so keenly that I can't put them out of my mind | 0.575 |
| stai_37 | Some unimportant thought runs through my mind and bothers me | 0.424 |
| **Factor 7: Restlessness** | | |
| mdi_8a | Have you felt very restless? | 0.705 |
| asrs_b_7 | How often do you feel restless or fidgety? | 0.633 |
| asrs_a_6 | How often do you feel overly active and compelled to do things, like you were driven by a motor? | 0.435 |
| stai_22 | I feel nervous and restless | 0.405 |
| phq_9_8 | Moving or speaking so slowly that other people could have noticed? Or the opposite — being so fidgety or restless that you have been moving around a lot more than usual | 0.352 |
| **Factor 8: Stress** | | |
| pss_10 | In the last month, how often have you felt difficulties were piling up so high that you could not overcome them? | 0.615 |
| pss_6 | In the last month, how often have you found that you could not cope with all the things that you had to do? | 0.468 |
| stai_28 | I feel that difficulties are piling up so that I cannot overcome them | 0.445 |
| pss_3 | In the last month, how often have you felt nervous and stressed? | 0.443 |
| pss_1 | In the last month, how often have you been upset because of something that happened unexpectedly? | 0.428 |
| **Factor 9: Autism Spectrum** | | |
| aq10_9 | I find it easy to work out what someone is thinking or feeling just by looking at their face | 0.714 |
| aq10_5 | I find it easy to 'read between the lines' when someone is talking to me | 0.694 |
| aq10_10 | I find it difficult to work out people's intentions | -0.670 |
| aq10_6 | I know how to tell if someone listening to me is getting bored | 0.628 |
| aq10_7 | When I'm reading a story I find it difficult to work out the characters' intentions | -0.346 |
| **Factor 10: Somatic Symptoms** | | |
| phq15_13 | Nausea, gas, or indigestion | 0.723 |
| phq15_12 | Constipation, loose bowels, or diarrhea | 0.677 |
| phq15_1 | Stomach pain | 0.660 |
| phq15_6 | Headaches | 0.337 |
| **Factor 11: Impulsivity** | | |
| asrs_b_12 | How often do you interrupt others when they are busy? | 0.628 |
| asrs_b_11 | How often do you have difficulty waiting your turn in situations when turn taking is required? | 0.600 |
| asrs_b_9 | How often do you find yourself talking too much when you are in social situations? | 0.596 |
| asrs_b_10 | When you're in a conversation, how often do you find yourself finishing the sentences of the people you are talking to, before they can finish them themselves? | 0.54 |
| **Higher-Level Factor 1: Affective Symptoms** | | |
| Factor 1 | Social Anxiety | 0.580 |
| Factor 2 | Self-Confidence | -0.754 |
| Factor 3 | Sleep | 0.323 |
| Factor 5 | Depression | 0.408 |

| | | |
|---|---|---|
| Factor 8 | Stress | 0.379 |
| Factor 9 | Autism Spectrum | 0.386 |
| **Higher-Level Factor 2: ADHD & Somatic** | | |
| Factor 6 | Negative Thoughts | 0.384 |
| Factor 7 | Restlessness | 0.545 |
| Factor 10 | Somatic Symptoms | 0.364 |
| Factor 11 | Impulsivity | 0.571 |

**Supplementary Table 5: Hierarchical two factor higher-level Exploratory Factor Analysis results of mental health survey items.** This includes pattern coefficients for each of the factors (bold represents items > absolute 0.3 loading), communality (commun), uniqueness (unique) and complexity (complex) estimates. The first higher-level factor encompassed affective symptoms (first level factors: social anxiety, self-confidence, sleep, depression, stress, autism spectrum). The second higher-level factor incorporated ADHD & somatic symptoms (negative thoughts, restlessness, somatic symptoms, impulsivity).

| | Affective Symptoms | ADHD & Somatic | Commun | Unique | Complex |
|---|---|---|---|---|---|
| **Self-Confidence** | -0.754 | *-0.014* | 0.58 | 0.42 | 1.00 |
| **Social Anxiety** | 0.580 | *-0.045* | 0.31 | 0.69 | 1.01 |
| **Depression** | 0.408 | *0.110* | 0.22 | 0.78 | 1.15 |
| **Autism Spectrum** | 0.386 | *-0.171* | 0.12 | 0.88 | 1.38 |
| **Stress** | 0.379 | *0.192* | 0.25 | 0.75 | 1.48 |
| **Sleep** | 0.323 | *0.281* | 0.27 | 0.73 | 1.96 |
| **Impulsivity** | *-0.151* | 0.571 | 0.27 | 0.73 | 1.14 |
| **Restlessness** | *0.141* | 0.545 | 0.39 | 0.61 | 1.13 |
| **Negative Thoughts** | *0.230* | 0.384 | 0.28 | 0.72 | 1.63 |
| **Somatic** | *0.016* | 0.364 | 0.14 | 0.86 | 1.00 |
| **Inattentiveness** | *0.213* | *0.283* | 0.18 | 0.82 | 1.86 |

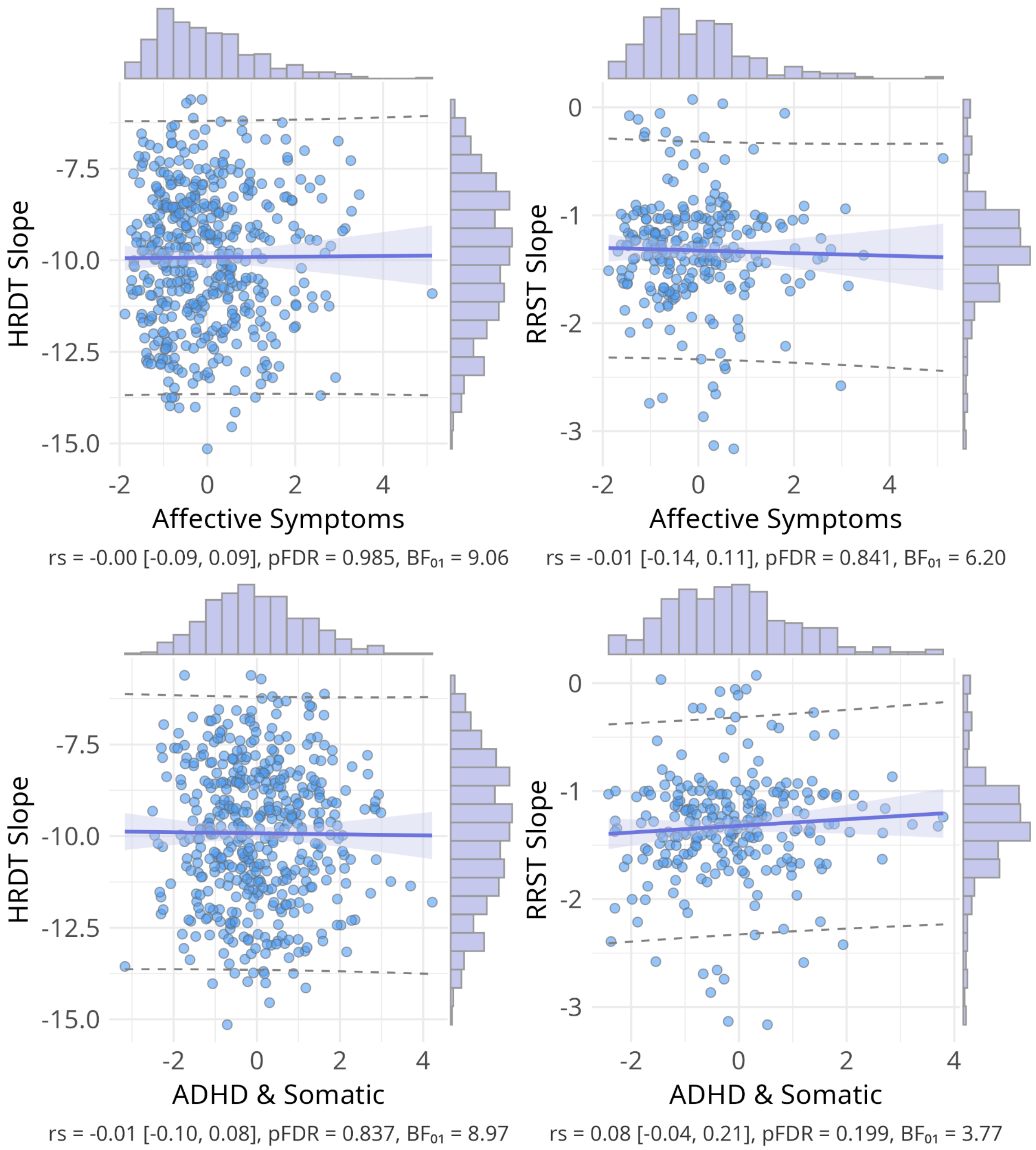


**Supplementary Figure 1: Scatter plots of the null relationship between interoceptive precision (slope) and mental health higher-level factors.** Scatter plots of Spearman correlations across interoception task variables and mental health higher-level factors. Interoceptive performance is demonstrated via precision (slope) for the cardiac (HRDT) and respiratory (RRST) psychophysical tasks. Above shows statistics for Spearman correlations with 95% confidence intervals in square brackets, p-values are corrected for multiple comparisons using the false discovery rate (Benjamini-Hochberg procedure at < 0.05), and null bayes factors (BF01).

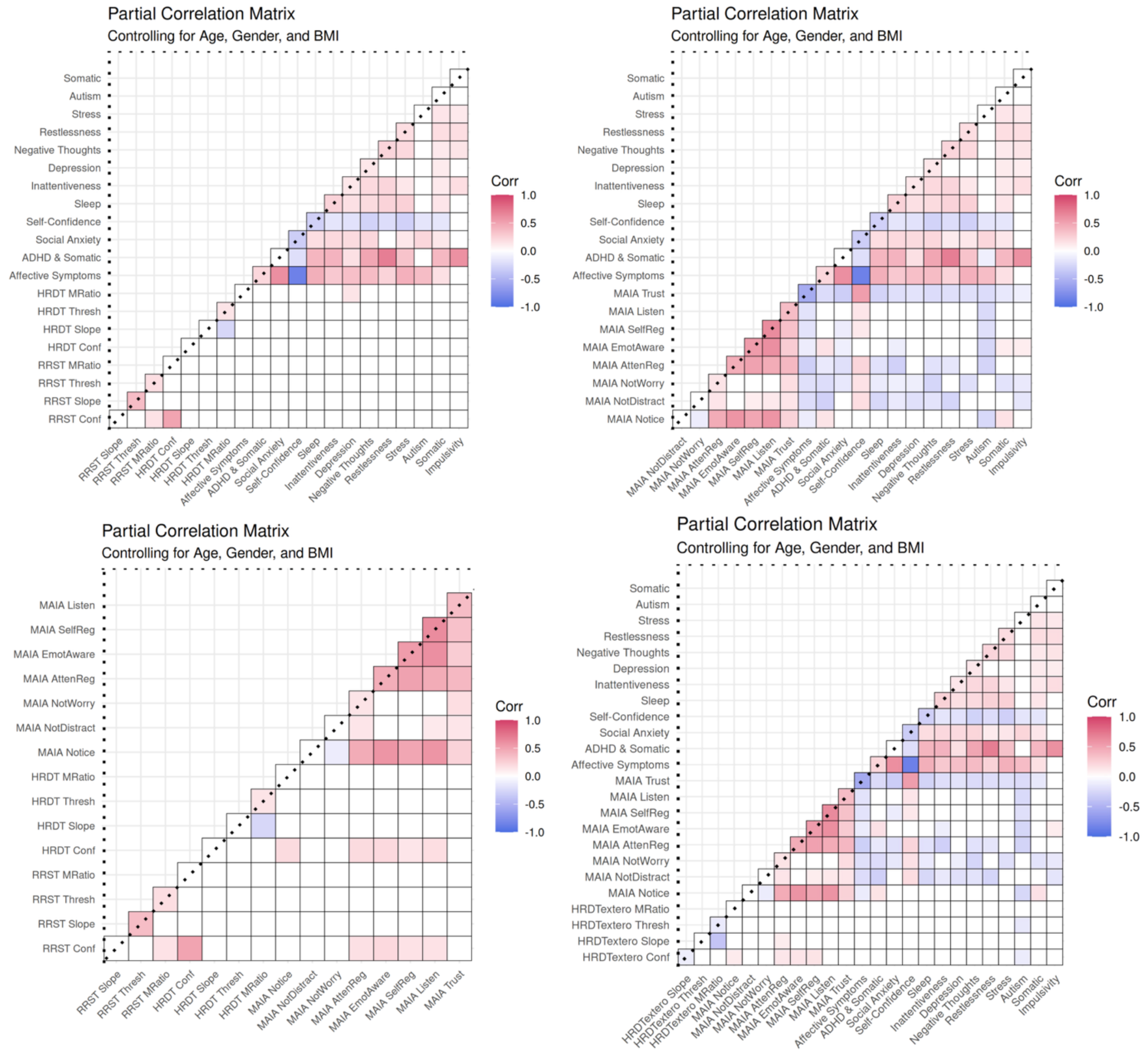


**Supplementary Figure 2: Partial correlation matrices controlling for age, gender, and body mass index.** Heatmap of partial cross-correlations between interoception/exteroception and the multi-factor mental health measures, controlling for age, gender, and body mass index. These matrices show Spearman correlation coefficients (colour scale from −1 in blue to 1 in red), with coloured squares showing FDR-corrected significant partial correlations (pFDR < 0.05). Top left is mental health factor partial correlations with interoceptive task metrics (RRST and HRDT), top right is mental health factor partial correlations with interoceptive sensibility (MAIA subscales), bottom left is interoceptive sensibility (MAIA subscales) partial correlations with interoceptive task metrics (RRST and HRDT), bottom right is auditory exteroceptive control task metrics partial correlations with mental health factors and interoceptive sensibility (MAIA subscales).

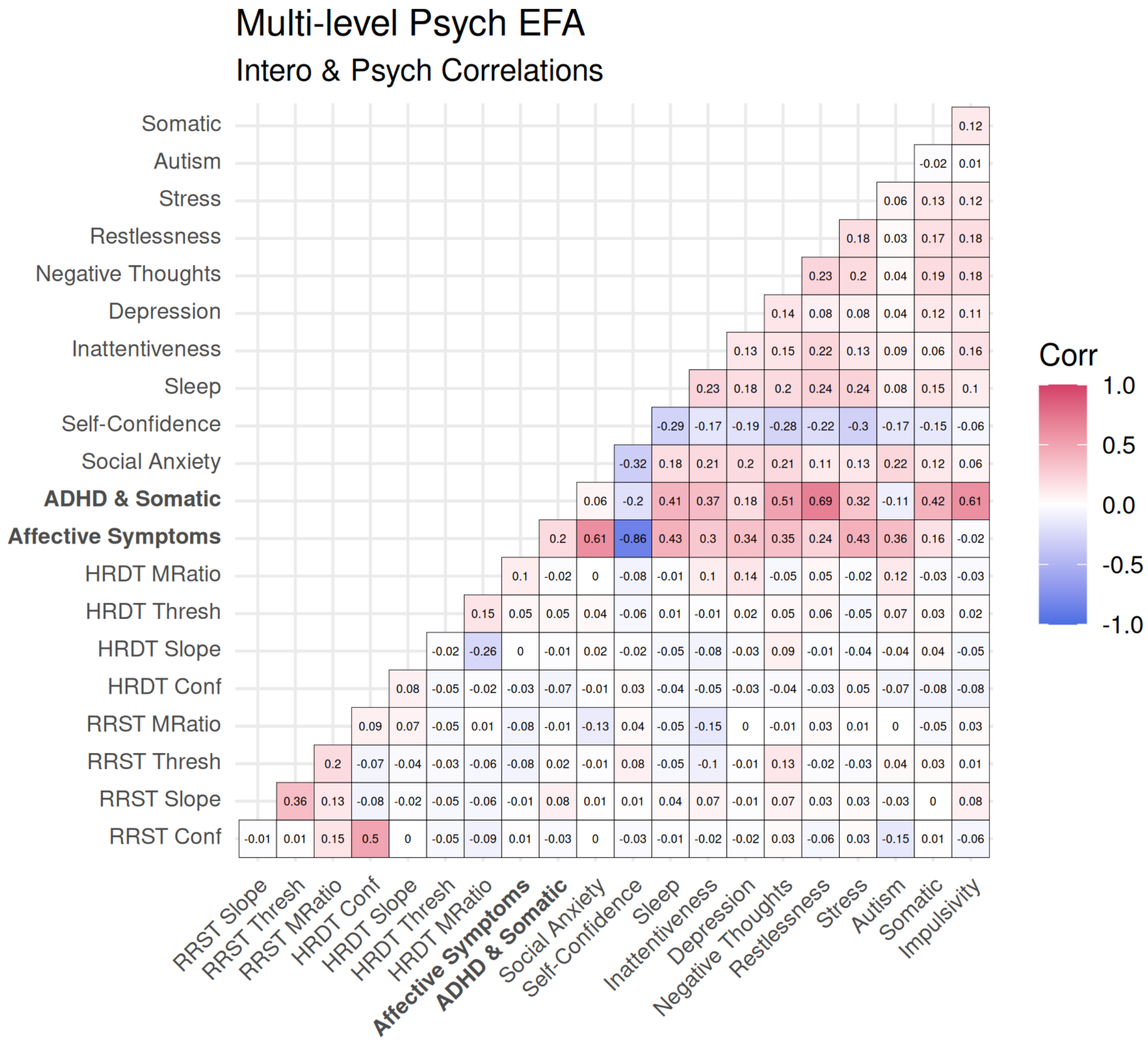


**Supplementary Figure 3: Full Spearman Correlation Coefficients between interoceptive psychophysics and mental health factors.** Cross Spearman correlations across interoception task variables and mental health factors (11 lower-level and two higher-level EFA). Interoceptive task variables include sensitivity (threshold), precision (slope), metacognitive bias (mean confidence), and metacognitive efficiency (M-Ratio) across cardiac (HRDT) and respiratory (RRST) domains. In red are positive correlations and in blue are negative correlations.

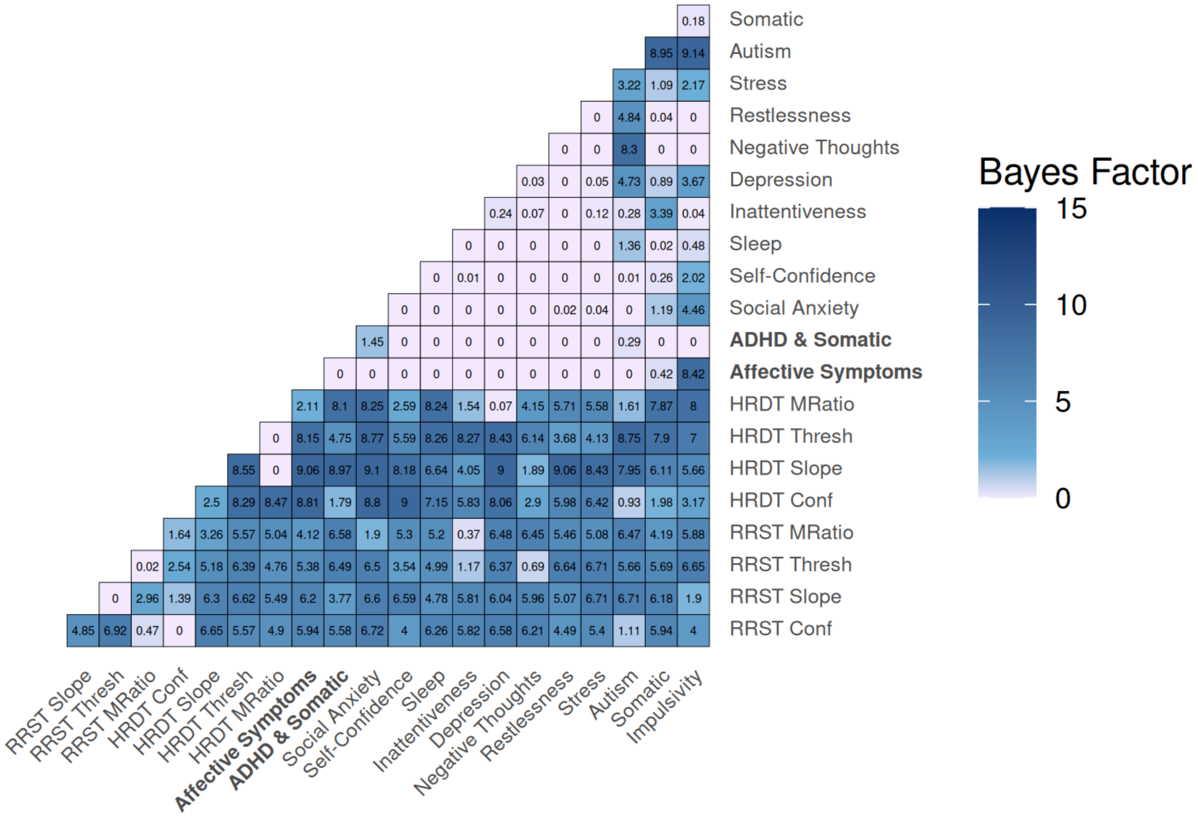


**Supplementary Figure 4: Full null Bayes factors (BF01) between interoceptive psychophysics and mental health factors.** Heatmap depicting null bayes factors (BF01) for interoception and mental health factors. The majority of observed Null Bayes Factors show anecdotal (BF01 > 1) or moderate (BF01 > 3) evidence for a lack of association[2], with slightly stronger null evidence for cardiac variables compared to respiratory variables.

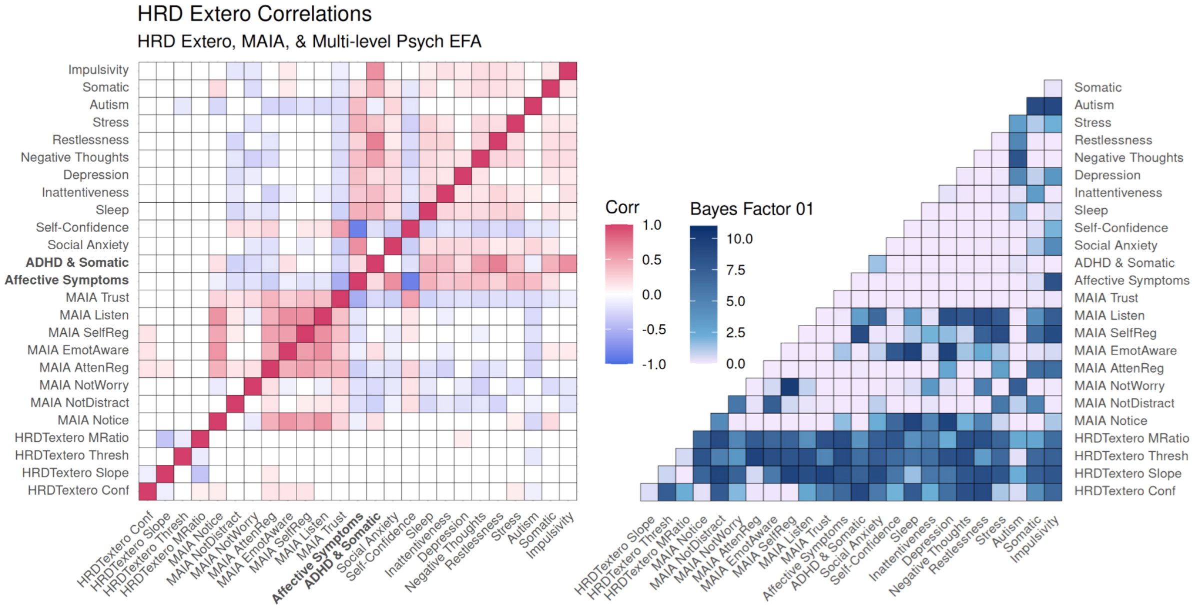


**Supplementary Figure 5: Relationship of auditory exteroceptive performance with mental health factors and interoceptive sensibility (MAIA subscales).** Left: Raw cross correlations across auditory exteroception task variables from the HRDT, 11 EFA mental health factors, two higher-level factors, and interoceptive sensibility (MAIA subscales). Upper diagonal depicts correlations after false discovery rate correction for multiple comparisons, lower depicts raw correlation coefficients. Right: Heatmap depicting raw null bayes factors (BF01) for exteroception, mental health factors, and interoceptive sensibility (MAIA subscales). BF01 > 3 is generally considered to be moderate evidence for the null[2].

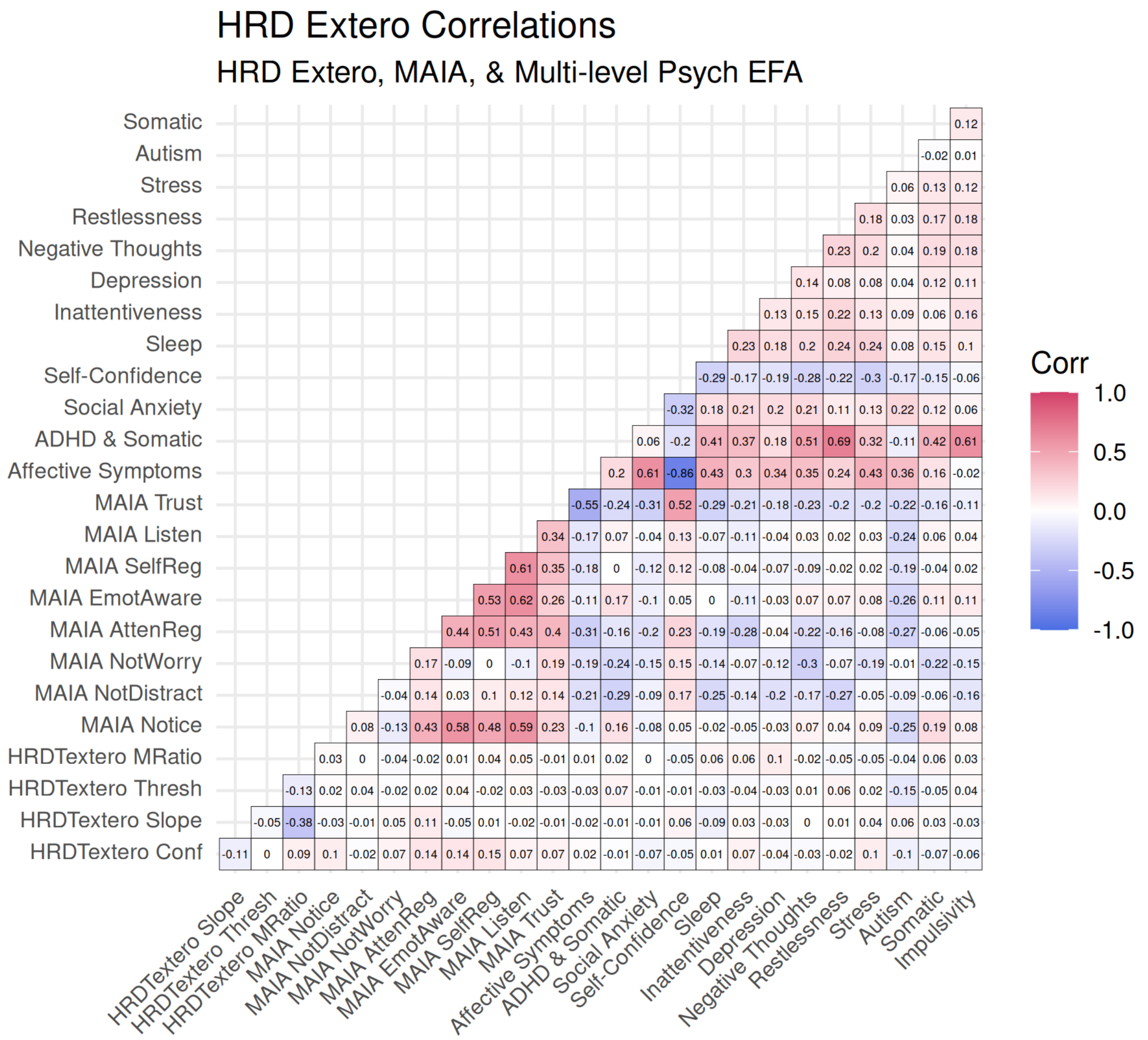


**Supplementary Figure 6: Full Spearman Correlation Coefficients between auditory exteroceptive performance and mental health factors and interoceptive sensibility (MAIA subscales).** Cross Spearman correlations between auditory exteroceptive control task metrics and mental health factors and interoceptive sensibility (MAIA subscales). In red are positive correlations and in blue are negative correlations.

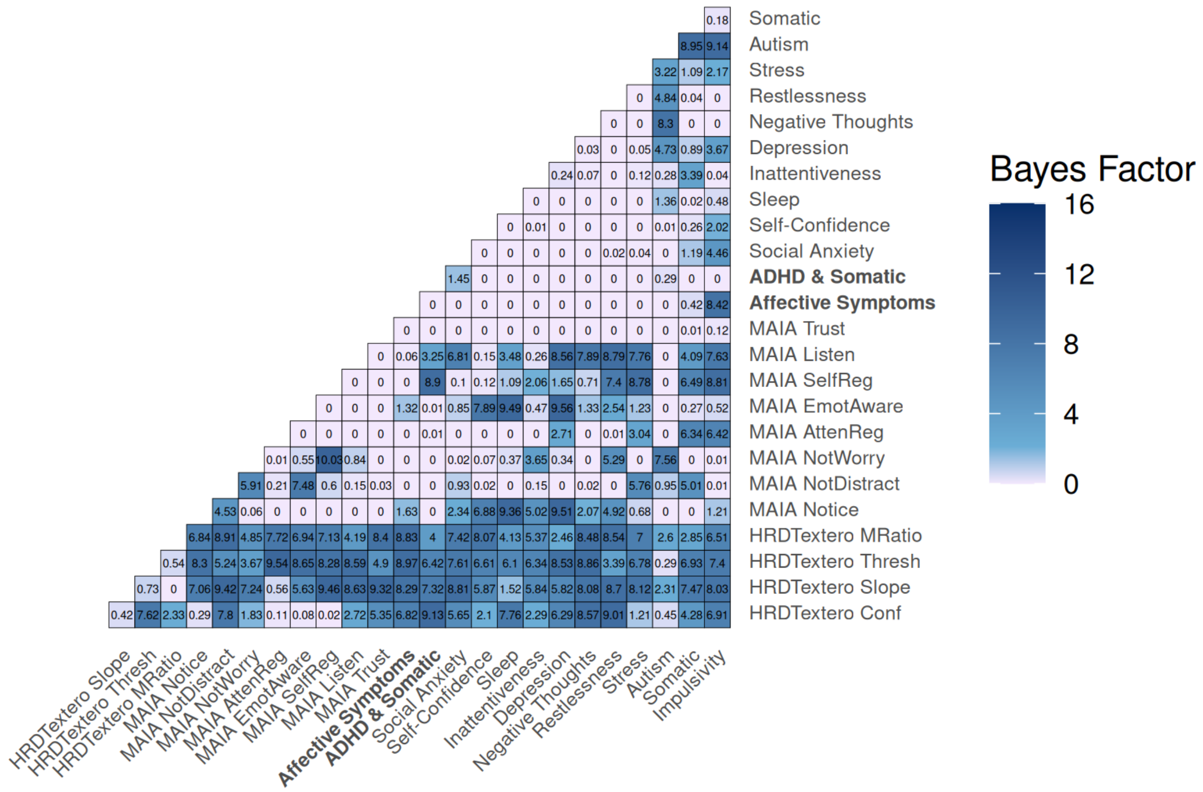


**Supplementary Figure 7: Full null Bayes factors (BF01) between auditory exteroceptive performance and mental health factors and interoceptive sensibility (MAIA subscales).** Heatmap of the null Bayes factors ($BF_{01}$) between auditory exteroceptive control task metrics and mental health factors and interoceptive sensibility (MAIA subscales). $BF_{01}$ > 1 reflects at least anecdotal evidence supporting no association.

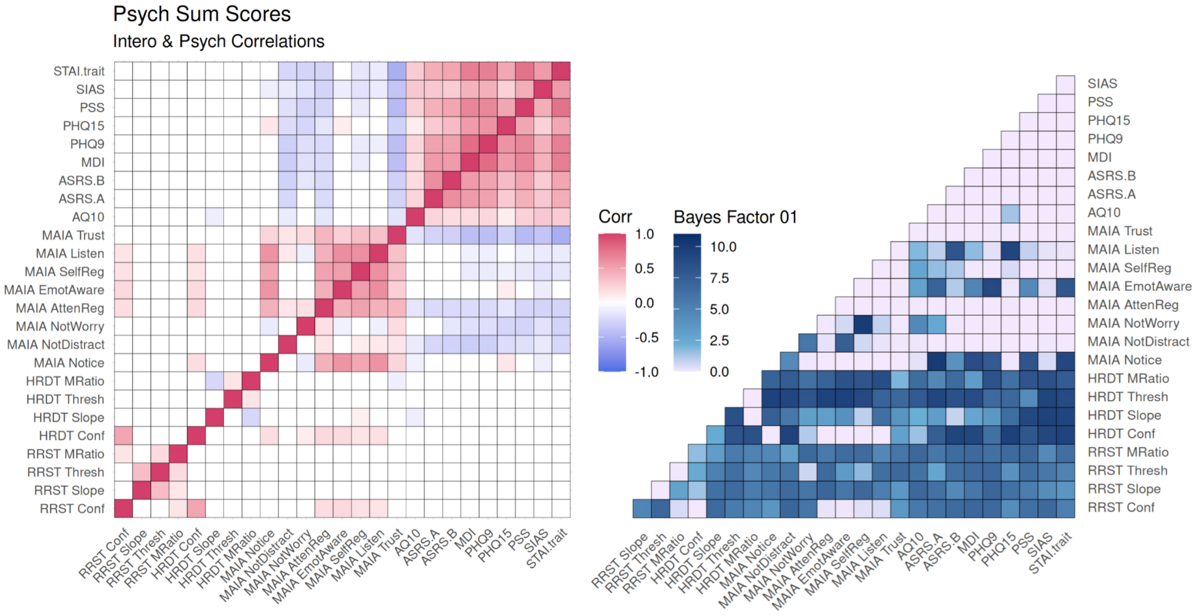


**Supplementary Figure 8: Correlations between mental health scale scores and interoception.** Left: Cross Spearman correlations across mental health scale scores and interoception task/sensibility variables. Mental health scale scores include: autism spectrum (AQ10), ADHD (ASRS A and B), depression (MDI and PHQ9), somatic symptoms (PHQ15), stress (PSS), social anxiety (SIAS), and trait anxiety (STAI trait). Interoceptive task variables include sensitivity (threshold), precision (slope), metacognitive bias (mean confidence), and metacognitive efficiency (M-Ratio) across cardiac (HRDT) and respiratory (RRST) domains. Interoceptive sensibility variables include the 8 MAIA subscales. Upper diagonal depicts correlations after false discovery rate correction for multiple comparisons (Benjamini-Hochberg procedure at < 0.05 shown in coloured boxes), lower depicts raw uncorrected correlation statistics. In red are significant positive correlations and in blue are significant negative correlations (p < 0.05 shown in coloured boxes). Right: Heatmap depicting null bayes factors (BF01) for interoception and mental health scale scores. The majority of observed Null Bayes Factors show anecdotal (BF01 > 1) or moderate (BF01 > 3) evidence for a lack of association between mental health scale scores and interoceptive psychophysics[2].

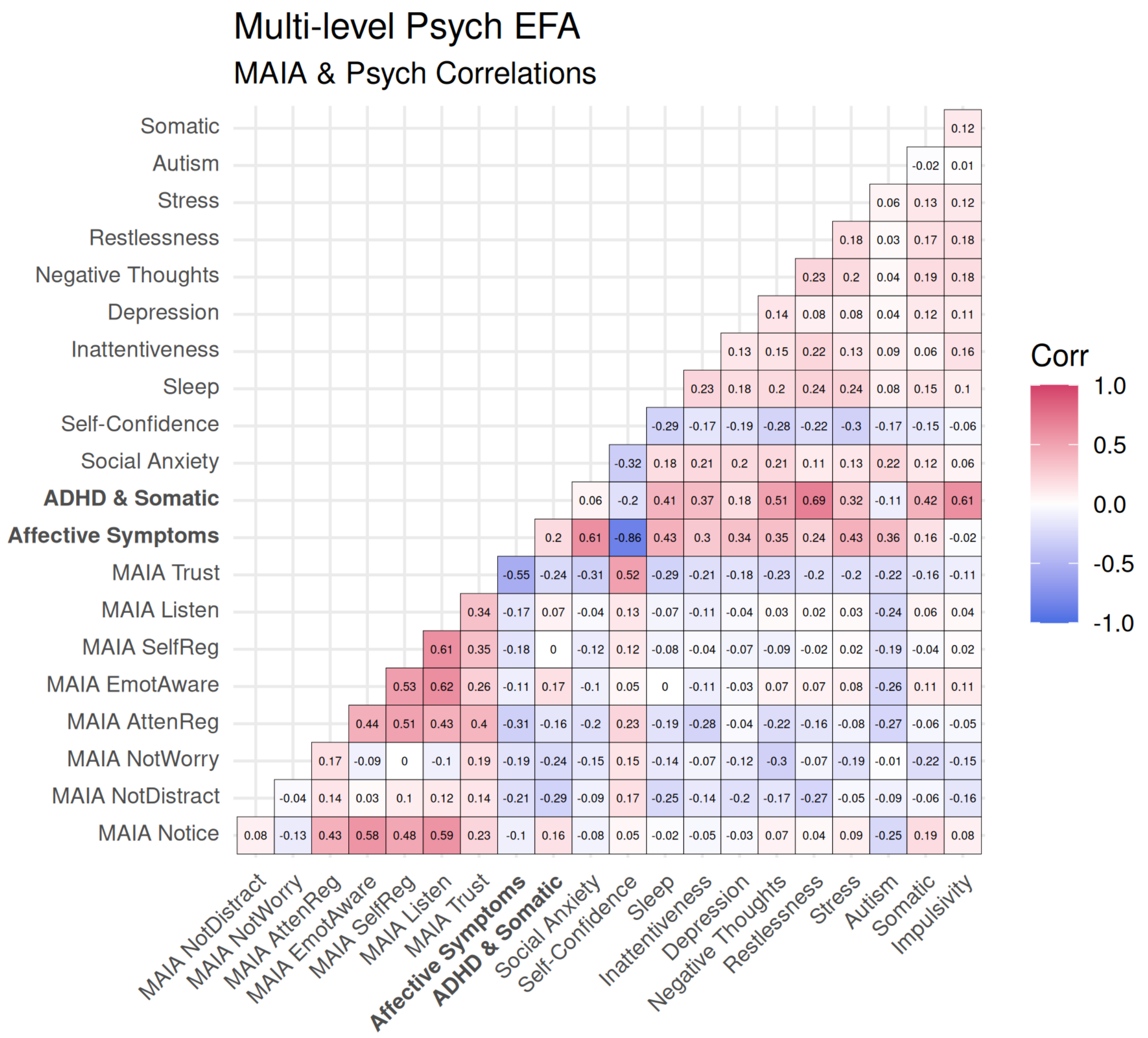


**Supplementary Figure 9: Full Spearman Correlation Coefficients between interoceptive sensibility and mental health factors.** Cross Spearman correlations between the eight MAIA subscales (interoceptive sensibility) and the multi-level mental health factors (11 lower-level and two higher-level EFA). In red are positive correlations and in blue are negative correlations.

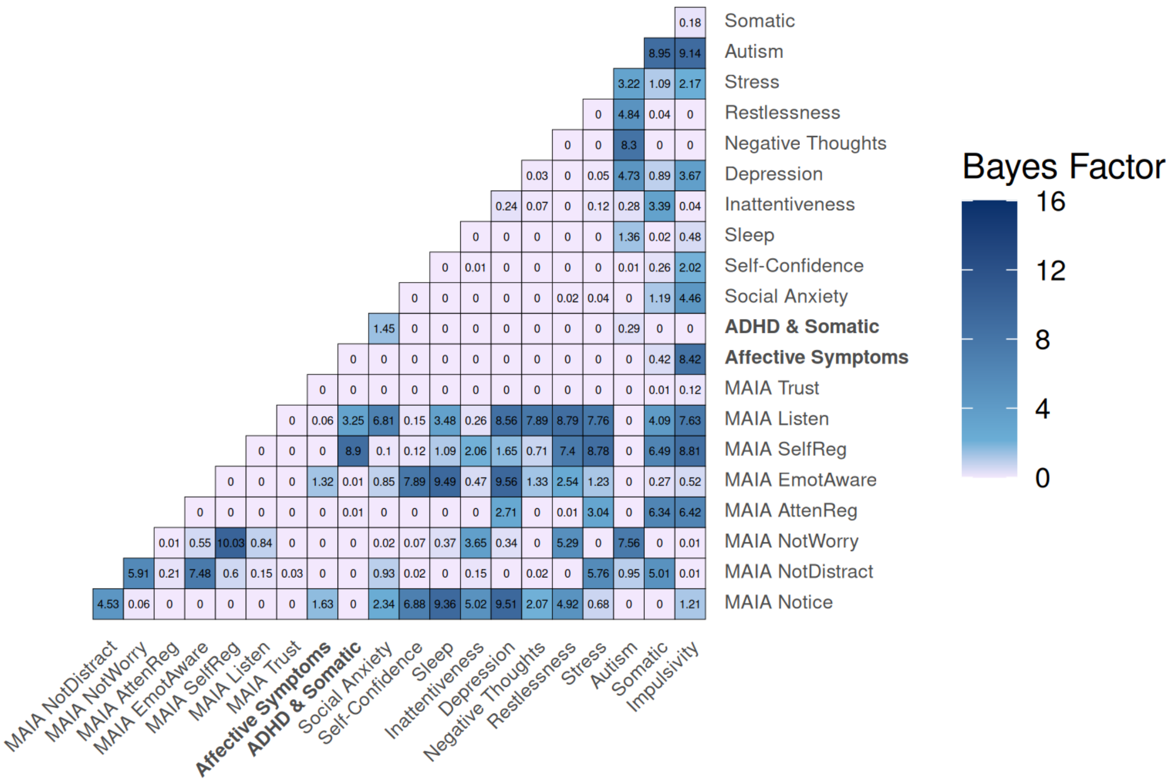


**Supplementary Figure 10: Full null Bayes factors (BF01) between interoceptive sensibility and mental health factors.** Heatmap of the null Bayes factors ($BF_{01}$) between interoceptive sensibility and mental health factors.

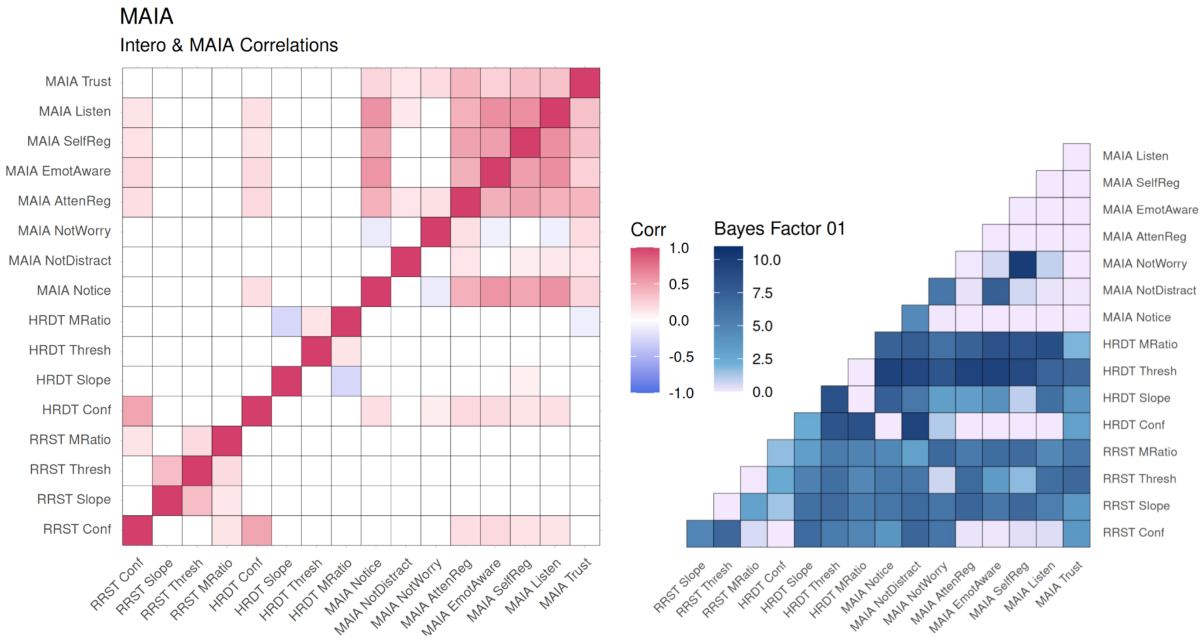


**Supplementary Figure 11: Correlations between interoceptive sensibility and interoceptive psychophysics**. Left: Cross Spearman correlations between the eight MAIA subscales and the respiratory (RRST) and cardiac (HRDT) interoception task measures. Interoceptive task variables include sensitivity (threshold (absolute for HRDT)), precision (slope), metacognitive bias (mean confidence), and metacognitive efficiency (M-Ratio) across cardiac (HRDT) and respiratory (RRST) domains. The upper diagonal depicts correlations that survive false discovery rate correction for multiple comparisons (Benjamini-Hochberg procedure at < 0.05 shown in upper diagonal coloured boxes), whereas the lower diagonal depicts correlation coefficients significant at the uncorrected threshold (uncorrected $p < .05$; lower diagonal coloured boxes). In red are significant positive correlations and in blue are significant negative correlations ($p < 0.05$ shown in coloured boxes). Right: Heatmap of the corresponding null Bayes factors ($BF_{01}$). $BF_{01} > 1$ reflects at least anecdotal evidence supporting no association.

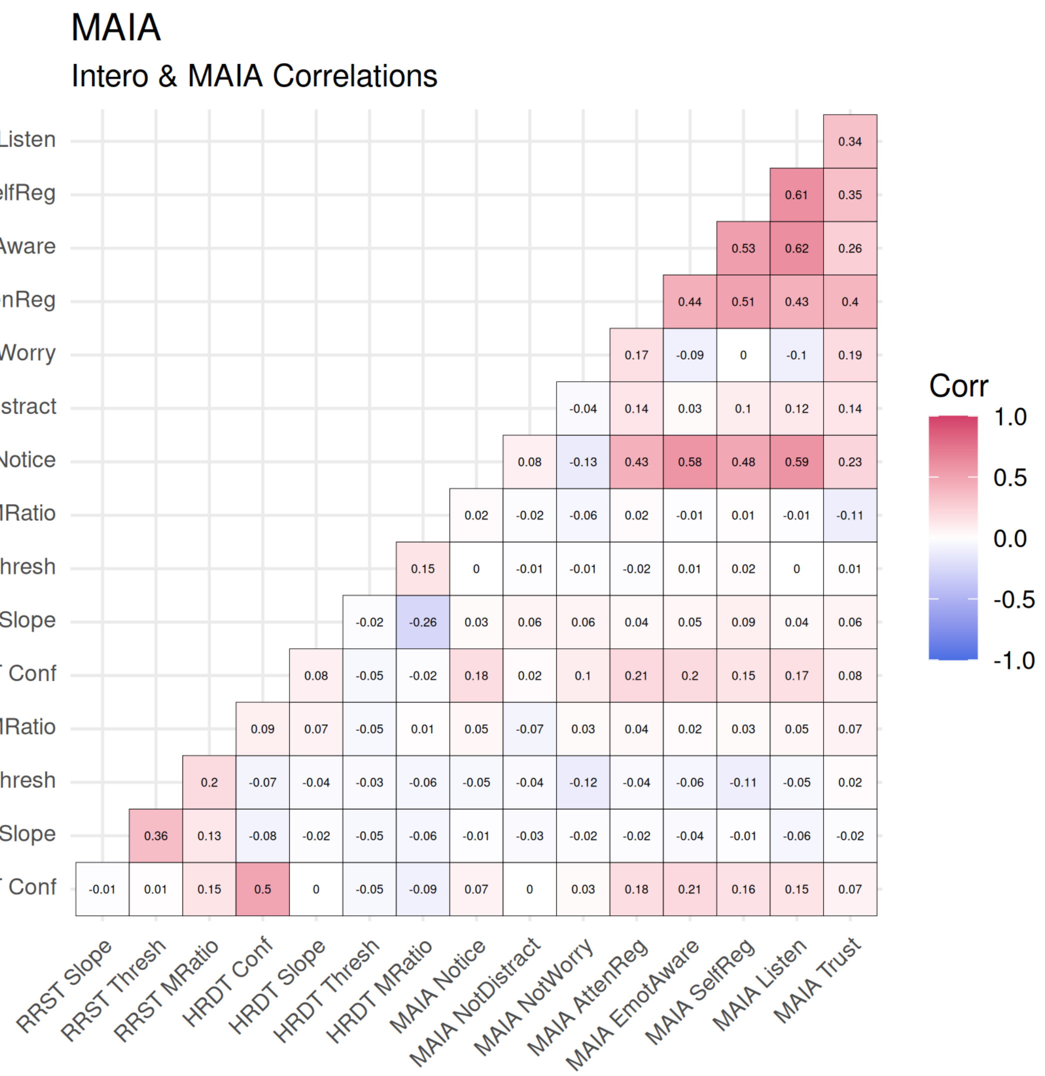


**Supplementary Figure 12: Full Spearman Correlation Coefficients between interoceptive sensibility and interoceptive psychophysics.** Cross Spearman correlations between the eight MAIA subscales and the respiratory (RRST) and cardiac (HRDT) interoception task measures. In red are positive correlations and in blue are negative correlations.

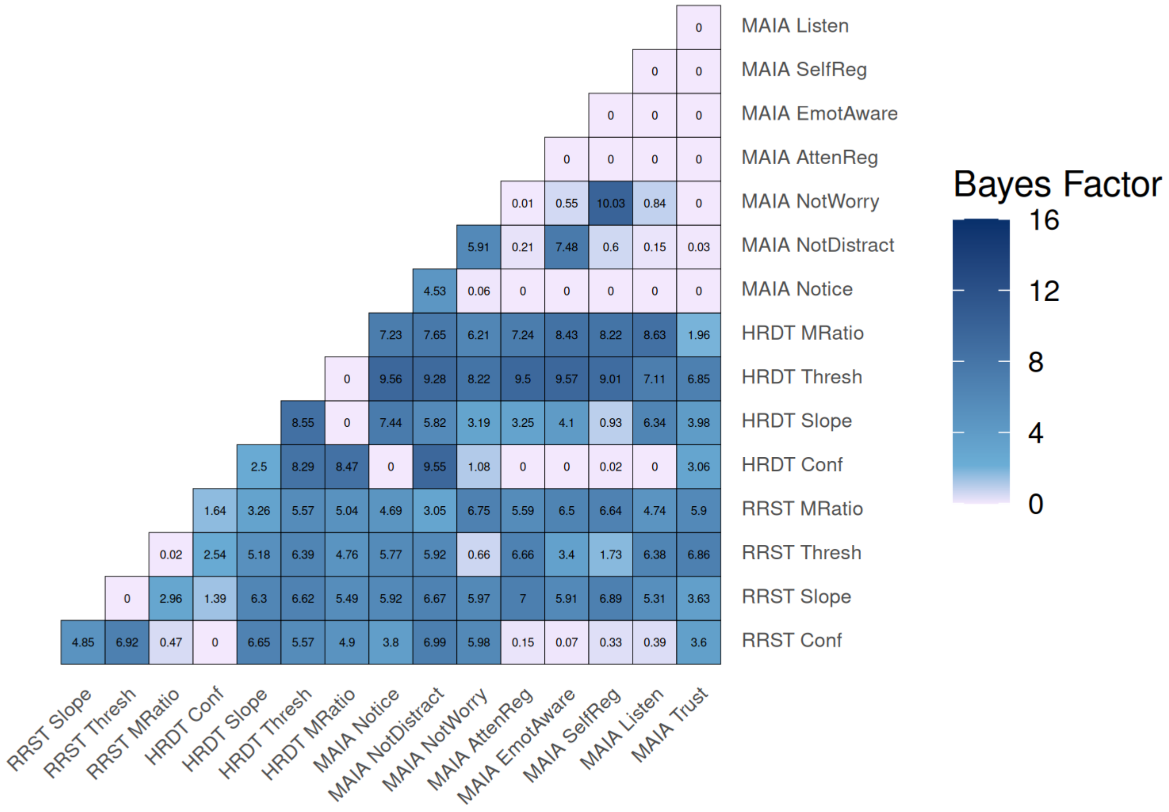


**Supplementary Figure 13: Full null Bayes factors (BF01) between interoceptive sensibility and interoceptive psychophysics**. Heatmap of the null Bayes factors ($BF_{01}$) between interoceptive sensibility and interoceptive psychophysics. $BF_{01} > 3$ reflects at least anecdotal evidence supporting no association.

**Supplementary Table 7:** Mental Health Inventory, including screening diagnosis cutoff percentages for the 500 individuals included in the Exploratory Factor Analyses.

| Survey | Full Name | N items | Range | Diagnosis Cutoff (%) |
|---|---|---|---|---|
| **AQ10**[3,4] | Autism Spectrum Quotient | 10 items | 1-4 | 8.6% *(binarised aq10 sum-score >= 6)*[3]*)* |
| **ASRS**[5] | Adult ADHD Self-Report Scale | 18 items | 1-5 | 17% *(binarised asrs.A sum-score >= 4)*[5] |
| **MD**[6,7] | Major Depression Inventory | 13 items | 1-6 | 1% high *(mdi sum-score >30)*<br>3.2% moderate *(mdi sum-score 26-30)*<br>4.6% mild *(mdi sum-score 21-25)*[6] |
| **PHQ9**[8,9] | Patient Health Questionnaire: depression module | 10 items | 1-4 | 0.2% severe *(phq9 sum-score >19)*<br>1.2% moderately-severe *(phq9 sum-score 15-19)*<br>8.2% moderate *(phq9 sum-score 10-14)*<br>28.6% mild *(phq9 sum-score 5-9)*[8] |
| **PHQ15**[10] | Patient Health Questionnaire: somatic symptoms module | 15 items | 1-3/4 | 2.8% high *(phq15 sum-score 15-30)*<br>13% medium *(phq15 sum-score 10-14)*[10] |
| **PSS**[11,12] | Perceived Stress Scale | 10 items | 1-5 | 2.6% high *(pss sum-score > 26)*<br>44.6% medium *(pss sum-score 14-26)*[13,14] |
| **SIAS**[15] | Social Interaction Anxiety Scale | 20 items | 0-4 | 20.4% *(sias sum-score >= 36)*[16] |
| **STAI (Trait)**[17] | State-Trait Anxiety Inventory P2 | 20 items | 1-4 | 20.2% *(stai-trait sum-score >= 44)*[18] |

**Supplementary Table 8: Mental health survey clinical cut-off samples sizes.** Above cut-off thresholds were defined as AQ10 ≥ 6 (autistic traits above screening threshold); ASRS-A ≥ 4 (ADHD symptoms above screening threshold); MDI ≥ 26 (moderate or higher depressive symptoms); PHQ-9 ≥ 10 (moderate or higher depressive symptoms); PHQ-15 ≥ 10 (medium or higher somatic symptom severity); PSS ≥ 14 (medium or higher perceived stress); SIAS ≥ 36 (elevated social anxiety); STAI-Trait ≥ 44 (elevated trait anxiety) (see Supplementary Table 7).

| Survey | Below cut-off N | Above cut-off N |
|---|---|---|
| **AQ10** | 457 | 43 |
| **ASRS.A** | 415 | 85 |
| **MDI** | 479 | 21 |
| **PHQ9** | 452 | 48 |
| **PHQ15** | 421 | 79 |
| **PSS** | 264 | 236 |
| **SIAS** | 398 | 102 |

**Supplementary Table 9**: Mann-Whitney U test analysis of interoception using survey clinical cut-off thresholds**.** Psychiatric screening cut-off thresholds were as follows: AQ10>=6, ASRS.A>=4, MDI>=26 (moderate+ depression), PHQ9>=10 (moderate+ depression), PHQ15>=10 (medium+ somatic symptoms), PSS>=14 (medium+ stress), SIAS>=36, STAI.trait>=44. Uncorrected significant results shown in bold (no significant results once corrected for multiple comparisons). See supplementary Table 7-8 for N and percentage of participants from full sample which cross diagnostic thresholds.

| Symptom | Interoception | N > cut-off | N < cut-off | Above cutoff: Median (IQR) | Below cutoff: Median (IQR) | W | r (rank-biserial) | p | p (FDR) | BF01 (null) |
|---|---|---|---|---|---|---|---|---|---|---|
| **AQ10** | HRDT Conf | 50 | 451 | 50.36 (15.15) | 54.09 (18.83) | 13476 | 0.2 | **0.0235** | 0.0939 | 0.537 |
| **AQ10** | HRDT MRatio | 37 | 370 | 0.69 (0.50) | 0.67 (0.48) | 6953 | 0.02 | 0.875 | 1 | 5.29 |
| **AQ10** | HRDT Slope | 50 | 451 | -9.77 (3.49) | -9.91 (2.81) | 10810 | -0.04 | 0.632 | 0.886 | 5.61 |
| **AQ10** | HRDT Thresh | 50 | 451 | 7.97 (12.57) | 9.49 (11.37) | 11697 | 0.04 | 0.664 | 0.886 | 5.31 |
| **AQ10** | RRST Conf | 23 | 243 | 53.40 (20.11) | 60.36 (16.02) | 3715 | 0.33 | **0.00908** | 0.0727 | 0.115 |
| **AQ10** | RRST MRatio | 23 | 233 | 0.91 (0.38) | 0.89 (0.28) | 2680 | 0 | 1 | 1 | 4.3 |
| **AQ10** | RRST Slope | 23 | 243 | -1.29 (0.38) | -1.29 (0.55) | 2413 | -0.14 | 0.28 | 0.56 | 1.93 |
| **AQ10** | RRST Thresh | 23 | 243 | 12.65 (1.48) | 13.15 (1.70) | 3179 | 0.14 | 0.276 | 0.56 | 3 |
| **ASRS.A** | HRDT Conf | 99 | 402 | 55.09 (19.80) | 52.89 (18.43) | 19123 | -0.04 | 0.548 | 0.73 | 7.98 |
| **ASRS.A** | HRDT MRatio | 79 | 328 | 0.66 (0.50) | 0.67 (0.50) | 12261 | -0.05 | 0.459 | 0.73 | 3.82 |
| **ASRS.A** | HRDT Slope | 99 | 402 | -10.52 (2.65) | -9.80 (2.77) | 22521 | 0.13 | **0.0422** | 0.169 | 1.2 |
| **ASRS.A** | HRDT Thresh | 99 | 402 | 9.45 (11.70) | 9.38 (11.57) | 19593 | -0.02 | 0.813 | 0.813 | 8.09 |
| **ASRS.A** | RRST Conf | 46 | 220 | 62.09 (18.26) | 59.64 (15.68) | 4567 | -0.1 | 0.299 | 0.599 | 3.22 |
| **ASRS.A** | RRST MRatio | 45 | 211 | 0.79 (0.33) | 0.90 (0.31) | 5532 | 0.17 | 0.0821 | 0.219 | 2.29 |
| **ASRS.A** | RRST Slope | 46 | 220 | -1.30 (0.57) | -1.29 (0.52) | 5279 | 0.04 | 0.645 | 0.737 | 4.39 |
| **ASRS.A** | RRST Thresh | 46 | 220 | 12.60 (1.46) | 13.17 (1.70) | 6151 | 0.22 | **0.0216** | 0.169 | 0.843 |
| **MDI** | HRDT Conf | 36 | 463 | 53.58 (26.24) | 53.53 (18.15) | 7819.5 | -0.06 | 0.537 | 0.86 | 2.88 |
| **MDI** | HRDT MRatio | 32 | 373 | 0.70 (0.45) | 0.67 (0.50) | 5303 | -0.11 | 0.296 | 0.77 | 2.8 |
| **MDI** | HRDT Slope | 36 | 463 | -10.87 (2.20) | -9.82 (2.80) | 9754 | 0.17 | 0.0885 | 0.617 | 1.98 |
| **MDI** | HRDT Thresh | 36 | 463 | 8.77 (9.41) | 9.48 (11.71) | 8292 | -0.01 | 0.96 | 0.96 | 5.37 |
| **MDI** | RRST Conf | 11 | 255 | 60.24 (14.68) | 60.01 (15.82) | 1329 | -0.05 | 0.77 | 0.96 | 3.29 |
| **MDI** | RRST MRatio | 11 | 245 | 0.90 (0.55) | 0.89 (0.28) | 1313 | -0.03 | 0.887 | 0.96 | 3.06 |
| **MDI** | RRST Slope | 11 | 255 | -1.29 (0.37) | -1.29 (0.55) | 1185 | -0.16 | 0.385 | 0.77 | 2.32 |
| **MDI** | RRST Thresh | 11 | 255 | 13.55 (1.82) | 13.13 (1.72) | 1046 | -0.25 | 0.154 | 0.617 | 0.841 |
| **PHQ9** | HRDT Conf | 65 | 433 | 54.80 (19.94) | 53.43 (18.49) | 13656.5 | -0.03 | 0.701 | 0.951 | 6.38 |
| **PHQ9** | HRDT MRatio | 59 | 345 | 0.67 (0.47) | 0.67 (0.51) | 10229 | 0.01 | 0.951 | 0.951 | 6.14 |
| **PHQ9** | HRDT Slope | 65 | 433 | -9.74 (2.73) | -9.87 (2.79) | 14221 | 0.01 | 0.891 | 0.951 | 6.88 |
| **PHQ9** | HRDT Thresh | 65 | 433 | 7.82 (7.72) | 9.57 (12.48) | 15321 | 0.09 | 0.249 | 0.951 | 2.34 |
| **PHQ9** | RRST Conf | 34 | 232 | 60.38 (22.55) | 59.77 (15.59) | 3679 | -0.07 | 0.528 | 0.951 | 4.5 |
| **PHQ9** | RRST MRatio | 33 | 223 | 0.93 (0.47) | 0.89 (0.27) | 3487 | -0.05 | 0.629 | 0.951 | 4.18 |
| **PHQ9** | RRST Slope | 34 | 232 | -1.31 (0.49) | -1.29 (0.53) | 4029 | 0.02 | 0.84 | 0.951 | 5.12 |
| **PHQ9** | RRST Thresh | 34 | 232 | 12.81 (1.32) | 13.15 (1.68) | 4170 | 0.06 | 0.59 | 0.951 | 5.07 |
| **PHQ15** | HRDT Conf | 98 | 415 | 53.67 (17.43) | 53.53 (19.24) | 20397.5 | 0 | 0.963 | 0.963 | 8.06 |
| **PHQ15** | HRDT MRatio | 85 | 332 | 0.66 (0.58) | 0.68 (0.48) | 14262 | 0.01 | 0.879 | 0.963 | 7.45 |
| **PHQ15** | HRDT Slope | 98 | 415 | -10.40 (2.64) | -9.80 (2.80) | 22820 | 0.12 | 0.0598 | 0.478 | 1.33 |
| **PHQ15** | HRDT Thresh | 98 | 415 | 10.26 (11.67) | 9.17 (11.25) | 18779 | -0.08 | 0.239 | 0.51 | 6.35 |
| **PHQ15** | RRST Conf | 45 | 222 | 61.07 (16.75) | 59.77 (15.79) | 4390 | -0.12 | 0.201 | 0.51 | 3.97 |

| | | | | | | | | | | |
|---|---|---|---|---|---|---|---|---|---|---|
| **PHQ15** | RRST MRatio | 45 | 212 | 0.81 (0.25) | 0.90 (0.31) | 5192 | 0.09 | 0.352 | 0.51 | 4.74 |
| **PHQ15** | RRST Slope | 45 | 222 | -1.37 (0.39) | -1.28 (0.53) | 5408 | 0.08 | 0.382 | 0.51 | 4.38 |
| **PHQ15** | RRST Thresh | 45 | 222 | 13.35 (1.49) | 13.11 (1.73) | 4471 | -0.1 | 0.268 | 0.51 | 2.92 |
| **PSS** | HRDT Conf | 246 | 252 | 54.24 (17.35) | 52.72 (19.99) | 29396.5 | -0.05 | 0.319 | 0.936 | 8.34 |
| **PSS** | HRDT MRatio | 206 | 198 | 0.68 (0.48) | 0.66 (0.52) | 19568 | -0.04 | 0.482 | 0.936 | 8.72 |
| **PSS** | HRDT Slope | 246 | 252 | -9.80 (2.75) | -9.95 (2.92) | 30695 | -0.01 | 0.852 | 0.936 | 9.9 |
| **PSS** | HRDT Thresh | 246 | 252 | 9.43 (11.34) | 9.33 (11.64) | 30619 | -0.01 | 0.815 | 0.936 | 10 |
| **PSS** | RRST Conf | 134 | 132 | 61.12 (15.44) | 59.12 (15.91) | 7994.5 | -0.1 | 0.176 | 0.936 | 3.45 |
| **PSS** | RRST MRatio | 131 | 125 | 0.88 (0.29) | 0.90 (0.29) | 8114 | -0.01 | 0.902 | 0.936 | 6.56 |
| **PSS** | RRST Slope | 134 | 132 | -1.30 (0.52) | -1.28 (0.55) | 8895 | 0.01 | 0.936 | 0.936 | 5.55 |
| **PSS** | RRST Thresh | 134 | 132 | 13.18 (1.57) | 12.96 (1.83) | 8432 | -0.05 | 0.512 | 0.936 | 6.49 |
| **SIAS** | HRDT Conf | 112 | 385 | 51.82 (16.58) | 54.16 (18.83) | 22920 | 0.06 | 0.309 | 0.578 | 5.77 |
| **SIAS** | HRDT MRatio | 95 | 309 | 0.67 (0.54) | 0.67 (0.47) | 14964 | 0.02 | 0.774 | 0.774 | 7.61 |
| **SIAS** | HRDT Slope | 112 | 385 | -10.12 (2.47) | -9.82 (2.82) | 23263 | 0.08 | 0.203 | 0.578 | 4.03 |
| **SIAS** | HRDT Thresh | 112 | 385 | 10.22 (10.89) | 8.97 (11.86) | 20338 | -0.06 | 0.361 | 0.578 | 7.2 |
| **SIAS** | RRST Conf | 55 | 211 | 59.27 (17.87) | 60.18 (16.36) | 6156 | 0.06 | 0.487 | 0.648 | 4.58 |
| **SIAS** | RRST MRatio | 52 | 204 | 0.84 (0.35) | 0.90 (0.29) | 5924 | 0.12 | 0.194 | 0.578 | 3.77 |
| **SIAS** | RRST Slope | 55 | 211 | -1.29 (0.42) | -1.29 (0.53) | 5511 | -0.05 | 0.567 | 0.648 | 5.88 |
| **SIAS** | RRST Thresh | 55 | 211 | 13.20 (1.58) | 13.11 (1.71) | 5337 | -0.08 | 0.36 | 0.578 | 3.83 |
| **STAI.trait** | HRDT Conf | 117 | 380 | 50.88 (16.59) | 54.24 (18.75) | 24123 | 0.09 | 0.164 | 0.427 | 4.49 |
| **STAI.trait** | HRDT MRatio | 96 | 308 | 0.66 (0.56) | 0.68 (0.48) | 15311 | 0.04 | 0.598 | 0.684 | 7.08 |
| **STAI.trait** | HRDT Slope | 117 | 380 | -10.22 (2.72) | -9.82 (2.77) | 23725 | 0.07 | 0.271 | 0.427 | 4.77 |
| **STAI.trait** | HRDT Thresh | 117 | 380 | 9.75 (10.44) | 9.06 (12.26) | 20776 | -0.07 | 0.285 | 0.427 | 8.01 |
| **STAI.trait** | RRST Conf | 63 | 203 | 58.76 (16.79) | 60.36 (15.70) | 6925 | 0.08 | 0.32 | 0.427 | 4.75 |
| **STAI.trait** | RRST MRatio | 62 | 194 | 0.81 (0.35) | 0.90 (0.32) | 6873 | 0.14 | 0.0907 | 0.427 | 2.42 |
| **STAI.trait** | RRST Slope | 63 | 203 | -1.22 (0.52) | -1.29 (0.55) | 5847 | -0.09 | 0.305 | 0.427 | 5.18 |
| **STAI.trait** | RRST Thresh | 63 | 203 | 13.03 (1.37) | 13.14 (1.82) | 6474 | 0.01 | 0.882 | 0.882 | 6.37 |

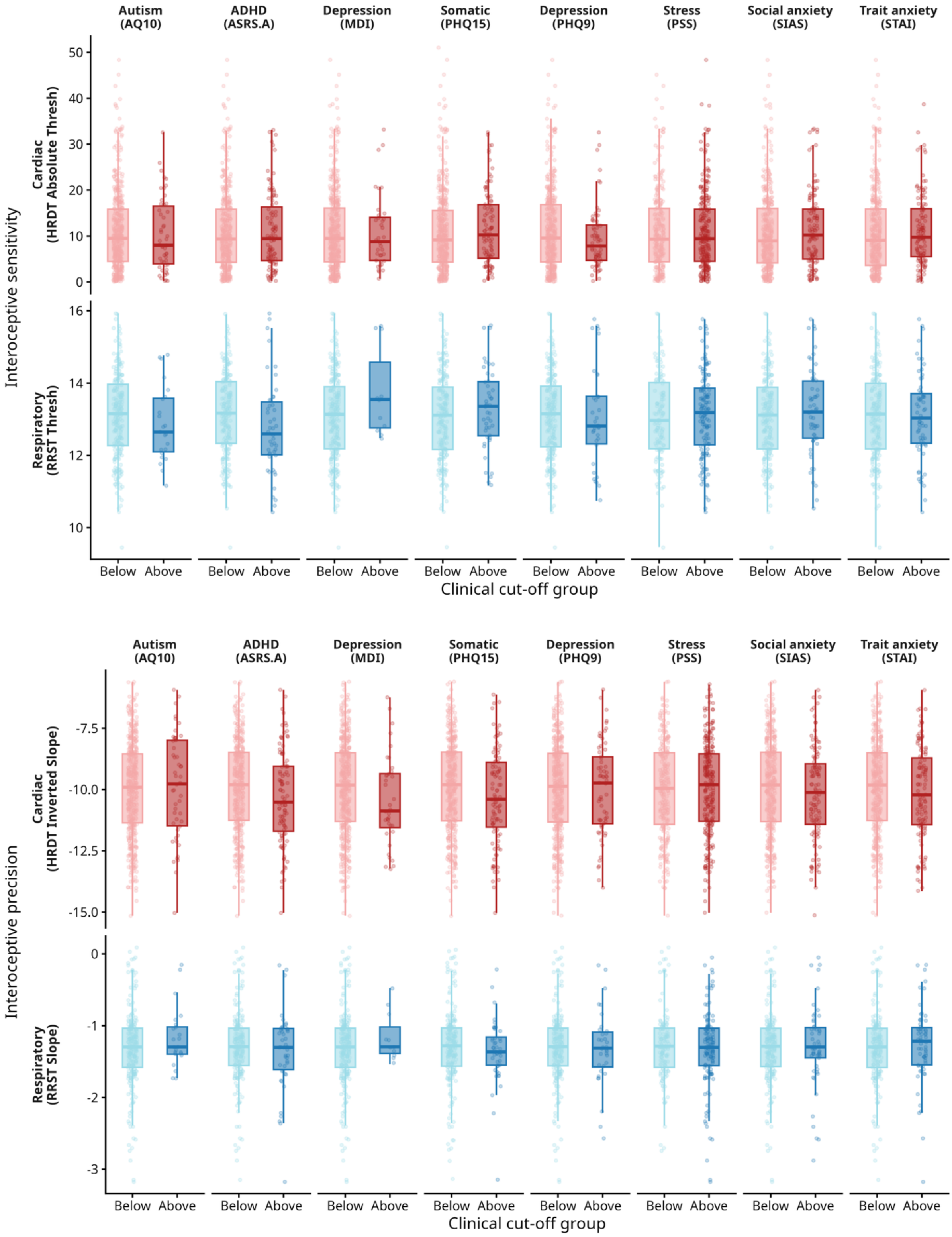


**Supplementary Figure 14: Boxplots of interoceptive performance across mental health survey cut-off groups.** Boxplots at the Top reflect interoceptive sensibility (HRDT/RRST threshold) and Bottom reflect interoceptive precision (HRDT/RRST slope) across groups above and below clinical cut-offs. Psychiatric screening cut-off thresholds were as follows: AQ10>=6, ASRS.A>=4, MDI>=26 (moderate+ depression), PHQ9>=10 (moderate+ depression), PHQ15>=10 (medium+ somatic symptoms), PSS>=14 (medium+ stress), SIAS>=36, STAI.trait>=44. Overall, there was minimal evidence for clinical group differences in interoception perceptual performance. While exploratory Mann–Whitney U tests indicated ADHD group differences in cardiac precision and respiratory sensibility, these effects did not remain significant after correction for multiple comparisons. See supplementary Table 7-8 for the total of participants which cross diagnostic thresholds and scoring details.

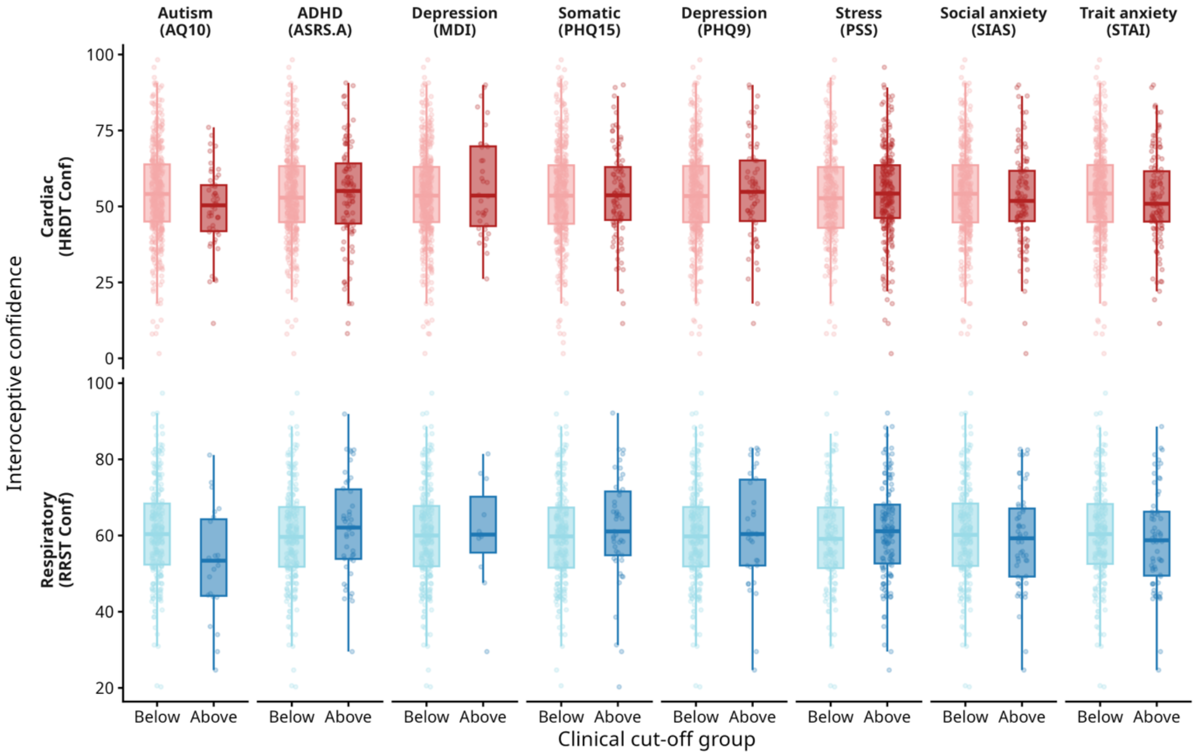


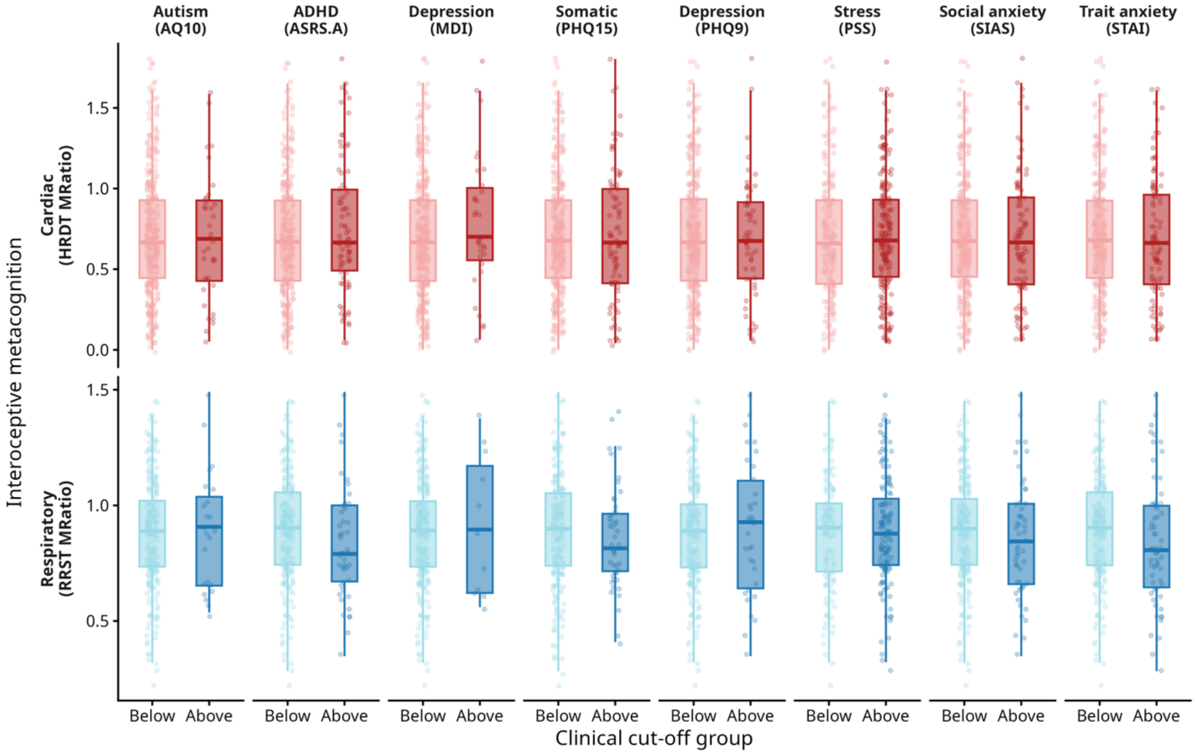


**Supplementary Figure 15: Boxplots of interoceptive metacognition across mental health survey cut-off groups.** Boxplots at the Top reflect interoceptive confidence (HRDT/RRST Conf) and Bottom reflect interoceptive metacognitive efficiency (HRDT/RRST M-Ratio) across groups above and below clinical cut-offs. Psychiatric screening cut-off thresholds were: AQ10>=6, ASRS.A>=4, MDI>=26 (moderate+ depression), PHQ9>=10 (moderate+ depression), PHQ15>=10 (medium+ somatic symptoms), PSS>=14 (medium+ stress), SIAS>=36, STAI.trait>=44. Overall, there was minimal evidence for clinical group differences in interoception metacognitive performance. While exploratory Mann–Whitney U tests indicated Autism group differences in cardiac and respiratory confidence, these effects did not remain significant after correction for multiple comparisons. See supplementary Table 7-8 for the total of participants which cross diagnostic thresholds and scoring details.

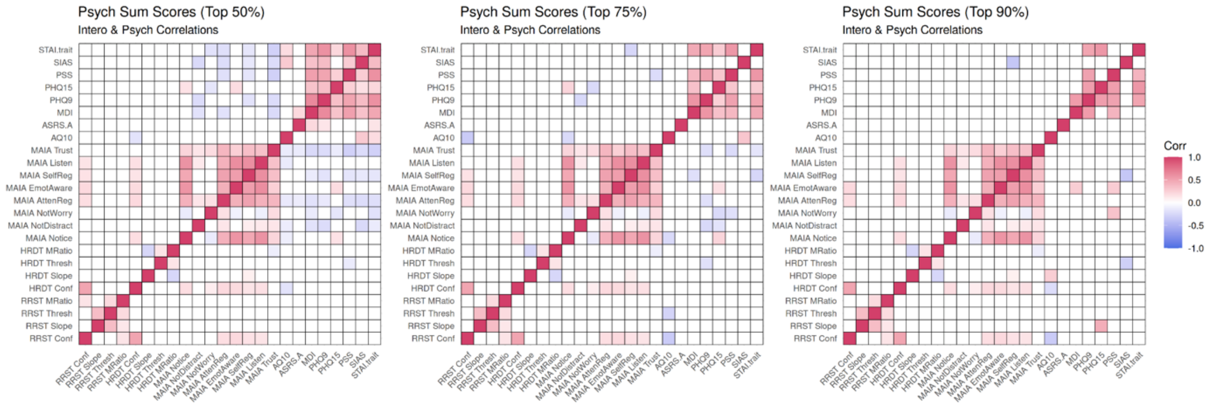


**Supplementary Figure 16: Upper symptom percentile-based analyses of interoceptive–mental health associations.** Spearman correlations between interoceptive task measures (cardiac HRDT and respiratory RRST sensitivity, precision, metacognitive bias, and metacognitive efficiency) and psychiatric symptom scores were recomputed after progressively restricting symptom distributions to higher severity ranges (top 50%, 75%, and 90% percentiles). Correlation coefficients are shown for each percentile threshold. False discovery rate (FDR) correction was applied within each percentile analysis. While a small number of associations reached significance at intermediate severity levels (e.g., autism traits with cardiac confidence at the top 50% analysis, and with respiratory confidence at the top 75% analysis), effect sizes did not systematically increase with greater symptom severity, and no associations survived correction at the highest severity threshold (top 90%).

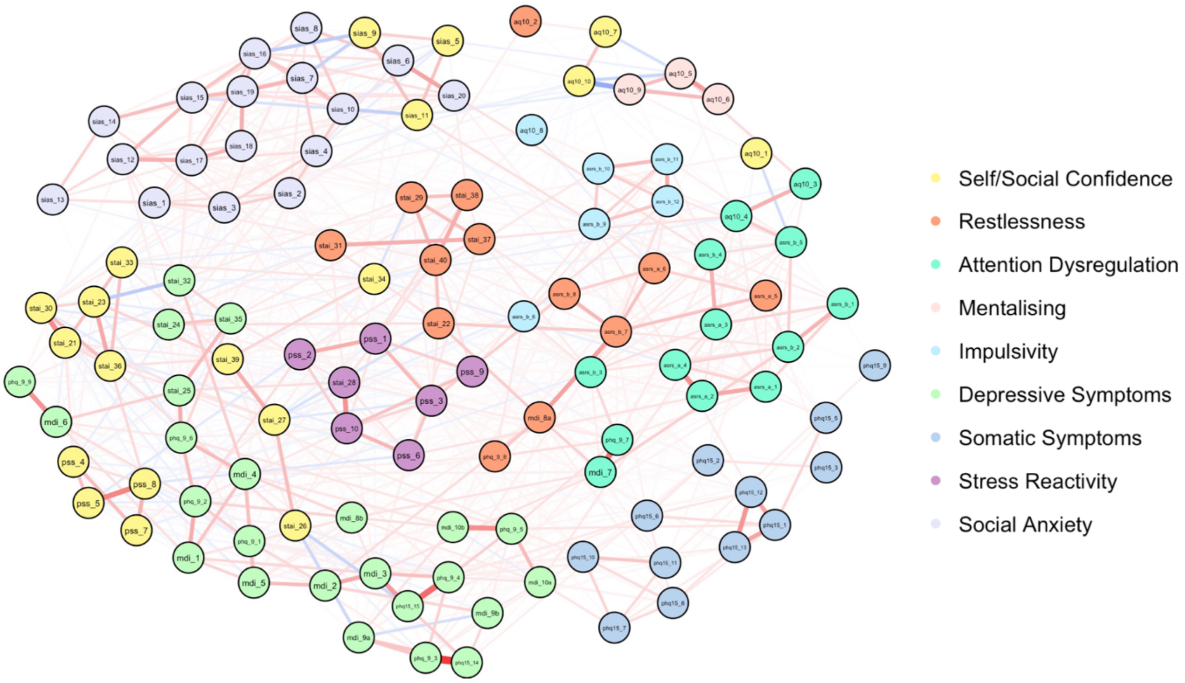


**Supplementary Figure 17: Psychiatric symptom network structure across questionnaire items.** Gaussian graphical model of item-level psychiatric symptoms estimated using EBICglasso. Nodes represent individual questionnaire items spanning autism (AQ10), ADHD (ASRS), depression (MDI, PHQ-9), somatic symptoms (PHQ-15), stress (PSS), social anxiety (SIAS), and trait anxiety (STAI-Trait). Edges represent partial correlations between symptoms after controlling for all other items, with red and blue edges indicating positive and negative associations, respectively (edge thickness reflects strength). Node colours denote 9 communities identified via spin-glass community detection, reflecting clusters of psychiatric symptoms as shown by the legend on the right (see Supplementary Table 10 for details of items belonging to each network community).

**Supplementary Table 10**: Mental health survey item details for each network community detected.

| Community | Item | Question |
|---|---|---|
| 1 | aq10_1 | I often notice small sounds when others do not |
| 1 | aq10_10 | I find it difficult to work out people's intentions |
| 1 | aq10_7 | When I'm reading a story I find it difficult to work out the characters' intentions |
| 1 | pss_4 | In the last month, how often have you felt confident about your ability to handle your personal problems? |
| 1 | pss_5 | In the last month, how often have you felt that things were going your way? |
| 1 | pss_7 | In the last month, how often have you been able to control irritations in your life? |
| 1 | pss_8 | In the last month, how often have you felt that you were on top of things? |
| 1 | sias_11 | I find it easy to think of things to talk about. |
| 1 | sias_5 | I find it easy to make friends my own age. |
| 1 | sias_9 | I am at ease meeting people at parties, etc. |
| 1 | stai_21 | I feel pleasant |
| 1 | stai_23 | I feel satisfied with myself |
| 1 | stai_26 | I feel rested |
| 1 | stai_27 | I am 'calm, cool, and collected' |
| 1 | stai_30 | I am happy |
| 1 | stai_33 | I feel secure |
| 1 | stai_34 | I make decisions easily |
| 1 | stai_36 | I am content |
| 1 | stai_39 | I am a steady person |
| 2 | aq10_2 | I usually concentrate more on the whole picture, rather than the small details |
| 2 | asrs_a_5 | How often do you fidget or squirm with your hands or feet when you have to sit down for a long time? |
| 2 | asrs_a_6 | How often do you feel overly active and compelled to do things, like you were driven by a motor? |
| 2 | asrs_b_7 | How often do you feel restless or fidgety? |
| 2 | asrs_b_8 | How often do you have difficulty unwinding and relaxing when you have time to yourself? |
| 2 | mdi_8a | Have you felt very restless? |
| 2 | phq_9_8 | Moving or speaking so slowly that other people could have noticed? Or the opposite — being so fidgety or restless that you have been moving around a lot more than usual |
| 2 | stai_22 | I feel nervous and restless |
| 2 | stai_29 | I worry too much over something that really doesn't matter |
| 2 | stai_31 | I have disturbing thoughts |
| 2 | stai_37 | Some unimportant thought runs through my mind and bothers me |
| 2 | stai_38 | I take disappointments so keenly that I can't put them out of my mind |
| 2 | stai_40 | I get in a state of tension or turmoil as I think over my recent concerns and interests |
| 3 | aq10_3 | I find it easy to do more than one thing at once |
| 3 | aq10_4 | If there is an interruption, I can switch back to what I was doing very quickly |
| 3 | asrs_a_1 | How often do you have trouble wrapping up the final details of a project, once the challenging parts have been done? |
| 3 | asrs_a_2 | How often do you have difficulty getting things in order when you have to do a task that requires organization? |
| 3 | asrs_a_3 | How often do you have problems remembering appointments or obligations? |
| 3 | asrs_a_4 | When you have a task that requires a lot of thought, how often do you avoid or delay getting started? |
| 3 | asrs_b_1 | How often do you make careless mistakes when you have to work on a boring or difficult project? |
| 3 | asrs_b_2 | How often do you have difficulty keeping your attention when you are doing boring or repetitive work? |
| 3 | asrs_b_3 | How often do you have difficulty concentrating on what people say to you, even when they are speaking to you directly? |
| 3 | asrs_b_4 | How often do you misplace or have difficulty finding things at home or at work? |
| 3 | asrs_b_5 | How often are you distracted by activity or noise around you? |
| 3 | mdi_7 | Have you had difficulty in concentrating, e.g. when reading the newspaper or watching TV? |
| 3 | phq_9_7 | Trouble concentrating on things, such as reading the newspaper or watching television |
| 4 | aq10_5 | I find it easy to 'read between the lines' when someone is talking to me |
| 4 | aq10_6 | I know how to tell if someone listening to me is getting bored |

| Community | Item | Question |
|---|---|---|
| 4 | aq10_9 | I find it easy to work out what someone is thinking or feeling just by looking at their face |
| 5 | aq10_8 | I like to collect information about categories of things (e.g. types of car, types of bird, types of train, types of plant etc) |
| 5 | asrs_b_10 | When you're in a conversation, how often do you find yourself finishing the sentences of the people you are talking to, before they can finish them themselves? |
| 5 | asrs_b_11 | How often do you have difficulty waiting your turn in situations when turn taking is required? |
| 5 | asrs_b_12 | How often do you interrupt others when they are busy? |
| 5 | asrs_b_6 | How often do you leave your seat in meetings or other situations in which you are expected to remain seated? |
| 5 | asrs_b_9 | How often do you find yourself talking too much when you are in social situations? |
| 6 | mdi_1 | Have you felt low in spirits or sad? |
| 6 | mdi_10a | Have you suffered from reduced appetite? |
| 6 | mdi_10b | Have you suffered from increased appetite? |
| 6 | mdi_2 | Have you lost interest in your daily activities? |
| 6 | mdi_3 | Have you felt lacking in energy and strength? |
| 6 | mdi_4 | Have you felt less self-confident? |
| 6 | mdi_5 | Have you had a bad conscience or feelings of guilt? |
| 6 | mdi_6 | Have you felt that life wasn't worth living? |
| 6 | mdi_8b | Have you felt subdued or slowed down? |
| 6 | mdi_9a | Have you been sleeping too little? |
| 6 | mdi_9b | Have you been sleeping too much? |
| 6 | phq15_14 | Trouble falling asleep, sleeping or sleeping too much |
| 6 | phq15_15 | Felt tired or only had little energy |
| 6 | phq_9_1 | Little interest or pleasure in doing things |
| 6 | phq_9_2 | Feeling down, depressed, or hopeless |
| 6 | phq_9_3 | Trouble falling or staying asleep, or sleeping too much |
| 6 | phq_9_4 | Feeling tired or having little energy |
| 6 | phq_9_5 | Poor appetite or overeating |
| 6 | phq_9_6 | Feeling bad about yourself — or that you are a failure or have let yourself or your family down |
| 6 | phq_9_9 | Thoughts that you would be better off dead or of hurting yourself in some way |
| 6 | stai_24 | I wish I could be as happy as others seem to be |
| 6 | stai_25 | I feel like a failure |
| 6 | stai_32 | I lack self-confidence |
| 6 | stai_35 | I feel inadequate |
| 7 | phq15_1 | Stomach pain |
| 7 | phq15_10 | Feeling your heart pound or race |
| 7 | phq15_11 | Shortness of breath |
| 7 | phq15_12 | Constipation, loose bowels, or diarrhea |
| 7 | phq15_13 | Nausea, gas, or indigestion |
| 7 | phq15_2 | Back pain |
| 7 | phq15_3 | Pain in your arms, legs, or joints (knees, hips, etc.) |
| 7 | phq15_5 | Pain or problems during sexual intercourse |
| 7 | phq15_6 | Headaches |
| 7 | phq15_7 | Chest pain |
| 7 | phq15_8 | Dizziness |
| 7 | phq15_9 | Fainting spells |
| 8 | pss_1 | In the last month, how often have you been upset because of something that happened unexpectedly? |
| 8 | pss_10 | In the last month, how often have you felt difficulties were piling up so high that you could not overcome them? |
| 8 | pss_2 | In the last month, how often have you felt that you were unable to control the important things in your life? |
| 8 | pss_3 | In the last month, how often have you felt nervous and stressed? |
| 8 | pss_6 | In the last month, how often have you found that you could not cope with all the things that you had to do? |
| 8 | pss_9 | In the last month, how often have you been angered because of things that happened that were outside of your control? |
| 8 | stai_28 | I feel that difficulties are piling up so that I cannot overcome them |
| 9 | sias_1 | I get nervous if I have to speak with someone in authority (teacher, boss, etc.). |
| 9 | sias_10 | I have difficulty talking with other people. |

| Community | Item | Question |
|---|---|---|
| 9 | sias_12 | I worry about expressing myself in case I appear awkward. |
| 9 | sias_13 | I find it difficult to disagree with another’s point of view. |
| 9 | sias_14 | I have difficulty talking to attractive persons of the opposite sex. |
| 9 | sias_15 | I find myself worrying that I won’t know what to say in social situations. |
| 9 | sias_16 | I am nervous mixing with people I don’t know well. |
| 9 | sias_17 | I feel I’ll say something embarrassing when talking. |
| 9 | sias_18 | When mixing in a group, I find myself worrying I will be ignored. |
| 9 | sias_19 | I am tense mixing in a group. |
| 9 | sias_2 | I have difficulty making eye contact with others. |
| 9 | sias_20 | I am unsure whether to greet someone I know only slightly." |
| 9 | sias_3 | I become tense if I have to talk about myself or my feelings. |
| 9 | sias_4 | I find it difficult to mix comfortably with the people I work with. |
| 9 | sias_6 | I tense up if I meet an acquaintance in the street. |
| 9 | sias_7 | When mixing socially, I am uncomfortable. |
| 9 | sias_8 | I feel tense if I am alone with just one other person. |

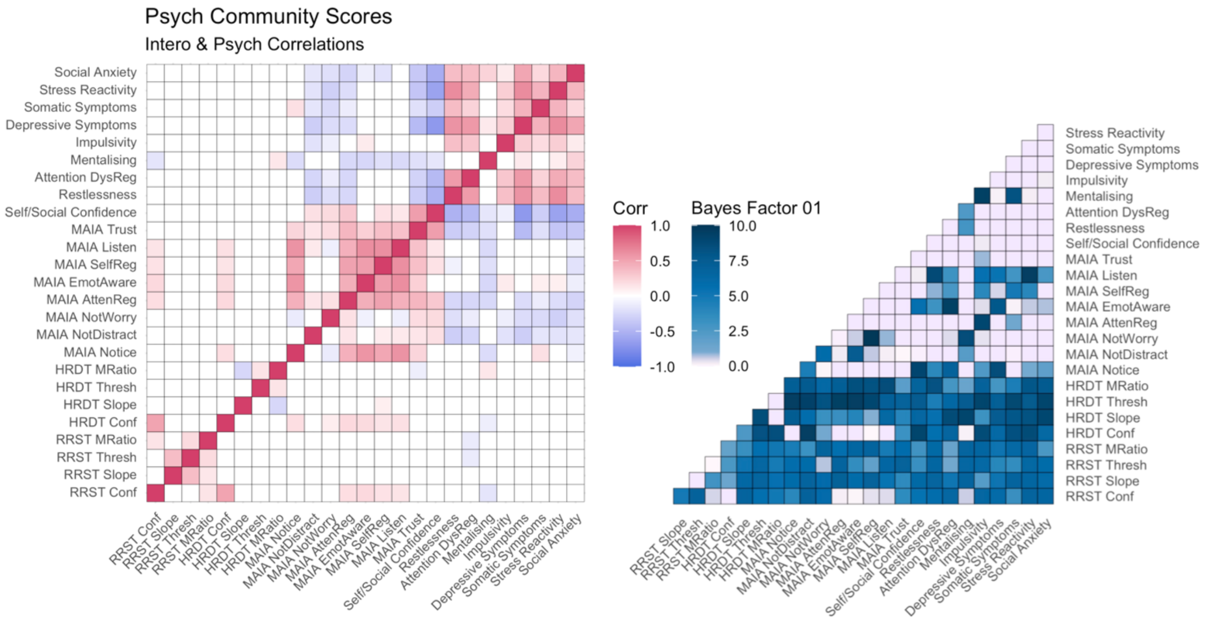


**Supplementary Figure 18: Correlations between mental health network community scores and interoception.** Left: Cross Spearman correlations across mental health network community scores and interoception task/sensibility variables. Mental health scale scores include: self/social-confidence, restlessness, attentional dysregulation, mentalising, impulsivity, depressive symptoms, somatic symptoms, stress reactivity, and social anxiety. Interoceptive task variables include sensitivity (threshold), precision (slope), metacognitive bias (mean confidence), and metacognitive efficiency (M-Ratio) across cardiac (HRDT) and respiratory (RRST) domains. Interoceptive sensibility variables include the 8 MAIA subscales. Upper diagonal depicts correlations after false discovery rate correction for multiple comparisons (Benjamini-Hochberg procedure at < 0.05 shown in coloured boxes), lower depicts raw uncorrected correlation statistics. In red are significant positive correlations and in blue are significant negative correlations ($p < 0.05$ shown in coloured boxes). Right: Heatmap depicting null bayes factors (BF01) for interoception and mental health scale scores. The majority of observed Null Bayes Factors show anecdotal (BF01 > 1) or moderate (BF01 > 3) evidence for a lack of association between mental health network community scores and interoceptive psychophysics.

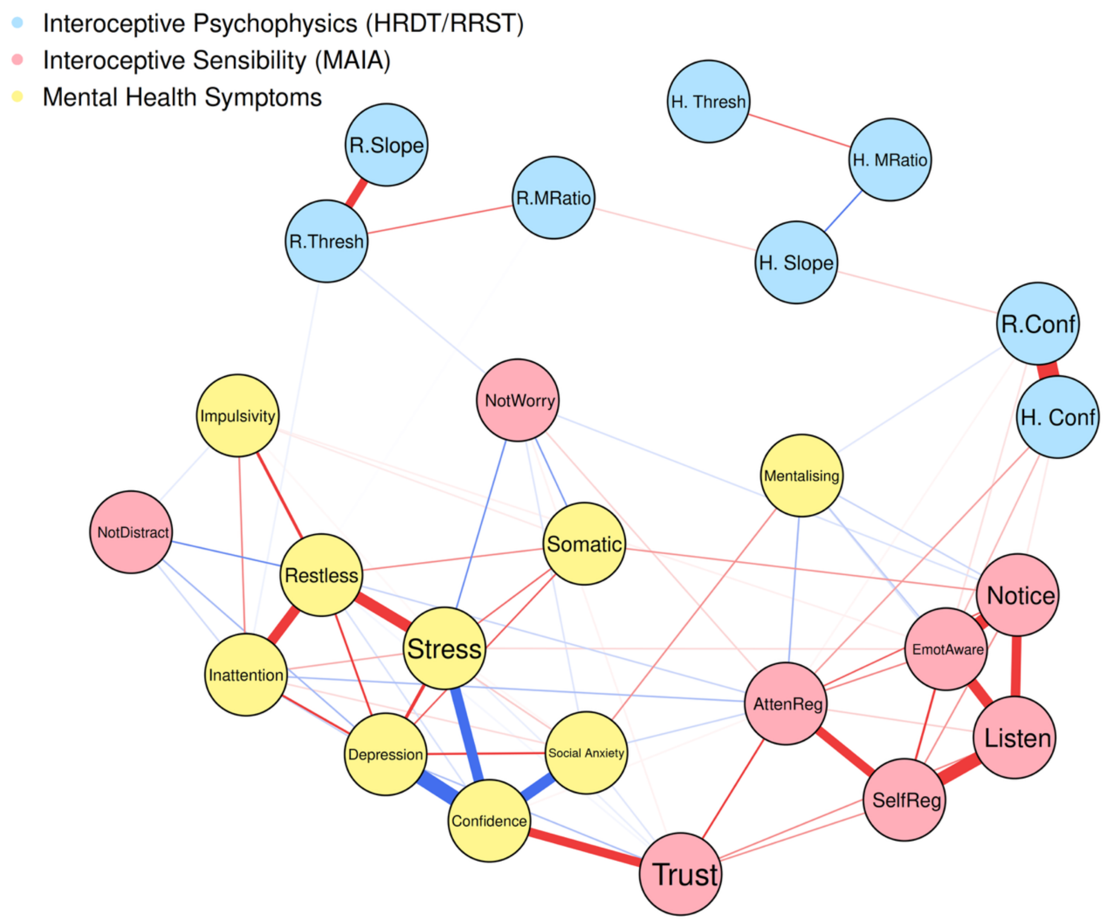


**Supplementary Figure 19. Joint network analysis of mental health and interoception.** EBICglasso-estimated network embedding mental health communities (yellow nodes from psychiatric network analysis of survey items in Figure 7), interoceptive psychophysics (cardiac HRDT ('H.' variables in blue) and respiratory RRST ('R.' nodes in blue) sensitivity ('Thresh'), precision ('Slope'), confidence ('Conf'), and metacognitive efficiency ('MRatio'), and interoceptive sensibility (MAIA subscales nodes in red). Edges represent regularised partial correlations after controlling for all other items, with red and blue edges indicating positive and negative associations, respectively (edge thickness reflects strength). Mental health–interoceptive psychophysics connections were sparse and weak, whereas several MAIA subscales showed moderate cross-domain associations, indicating stronger coupling of subjective interoceptive sensibility than objective interoceptive performance with psychiatric symptom domains.

**Supplementary Table 11**: **Joint network analysis of mental health network communities and interoception psychophysics (cardiac HRDT and respiratory RRST) and sensibility (MAIA subscales).** Non-zero edge weights of the joint network analysis of mental health and interoception. This shows predominant connectivity between mental health and interoceptive sensibility (MAIA), in comparison to weak and mostly non-zero connectivity with interoceptive psychophysics.

| Interoception Category | Mental Health Node | Interoception Node | Edge Weight |
|---:|---:|---:|---:|
| MH–MAIA | Self/Social Confidence | MAIA Trust | 0.230 |
| MH–MAIA | Restlessness | MAIA NotDistract | -0.115 |
| MH–MAIA | Stress Reactivity | MAIA NotWorry | -0.111 |
| MH–MAIA | Somatic Symptoms | MAIA NotWorry | -0.096 |
| MH–MAIA | Somatic Symptoms | MAIA Notice | 0.076 |
| MH–MAIA | Mentalising | MAIA AttenReg | -0.072 |
| MH–MAIA | Depressive Symptoms | MAIA NotDistract | -0.070 |
| MH–MAIA | Attention DysReg | MAIA AttenReg | -0.064 |
| MH–MAIA | Depressive Symptoms | MAIA Trust | -0.063 |
| MH–MAIA | Mentalising | MAIA EmotAware | -0.048 |
| MH–MAIA | Restlessness | MAIA AttenReg | -0.041 |
| MH–MAIA | Mentalising | MAIA Notice | -0.040 |
| MH–MAIA | Social Anxiety | MAIA AttenReg | -0.040 |
| MH–MAIA | Attention DysReg | MAIA NotDistract | -0.035 |
| MH–MAIA | Stress Reactivity | MAIA EmotAware | 0.029 |
| MH–MAIA | Mentalising | MAIA Listen | -0.025 |
| MH–MAIA | Social Anxiety | MAIA NotWorry | -0.025 |
| MH–MAIA | Social Anxiety | MAIA Trust | -0.023 |
| MH–Psychophysics | Mentalising | RRST Conf | -0.022 |
| MH–MAIA | Impulsivity | MAIA NotDistract | -0.019 |
| MH–MAIA | Stress Reactivity | MAIA Trust | -0.016 |
| MH–Psychophysics | Attention DysReg | RRST Thresh | -0.013 |
| MH–MAIA | Impulsivity | MAIA EmotAware | 0.011 |
| MH–MAIA | Restlessness | MAIA Trust | -0.008 |
| MH–MAIA | Self/Social Confidence | MAIA AttenReg | 0.007 |
| MH–Psychophysics | Attention DysReg | RRST MRatio | -0.002 |

**Supplementary Table 12: Bridge strength from interoceptive variables to mental health symptoms.** Bridge strength quantifies the summed absolute edge weights linking each psychophysical or interoceptive sensibility node to nodes in the estimated Gaussian graphical network model. Communities were defined a priori based on psychophysical interoceptive performance, interoceptive sensibility (MAIA), and mental health symptom dimensions. Larger values indicate stronger cross-community connectivity.

| Interoception Node | Interoception Category | Bridge strength |
|---:|---:|---:|
| MAIA Trust | MAIA | 0.340 |
| MAIA AttenReg | MAIA | 0.301 |
| MAIA NotWorry | MAIA | 0.257 |
| MAIA NotDistract | MAIA | 0.239 |
| MAIA EmotAware | MAIA | 0.160 |
| HRDT Conf | Psychophysics | 0.136 |
| MAIA Notice | MAIA | 0.133 |
| RRST Conf | Psychophysics | 0.055 |
| RRST Thresh | Psychophysics | 0.038 |
| MAIA Listen | MAIA | 0.025 |
| MAIA SelfReg | MAIA | 0.004 |
| RRST MRatio | Psychophysics | 0.002 |
| RRST Slope | Psychophysics | 0.000 |
| HRDT Slope | Psychophysics | 0.000 |
| HRDT Thresh | Psychophysics | 0.000 |
| HRDT MRatio | Psychophysics | 0.000 |

**Supplementary Table 13: Semantic similarity analysis of interoceptive sensibility scale: MAIA[1].** Semantic similarity analysis of the MAIA survey was completed with the online Semantic Scale Network tool[19]. The top 15 scales with the highest semantic similarity (cosine similarity) are shown.

| Scales | Similarity |
|---|---|
| sexual objectification - body evaluation (Kozee et al., 2007) | 0.627 |
| transgender congruence[19] | 0.534 |
| body awareness[20] | 0.525 |
| carroll depression - hypochondriasis (Feinberg & Carroll, 1986) | 0.514 |
| concept of health physical functioning[21] | 0.489 |
| health orientation - hs[22] | 0.474 |
| Mindfulness (Baer et al., 2004) | 0.451 |
| disgust - death[23] | 0.439 |
| tetradimensional depression - depressive mood[24] | 0.437 |
| self-report symptom - somatization | 0.436 |
| adolescent femininity ideology (Tolman & Porche, 2000) | 0.429 |
| competitive state anxiety - somatic anxiety[25] | 0.422 |
| health orientation - pho[22] | 0.412 |
| body-centred countertransference (Egan & Carr, 2008) | 0.406 |
| kellner symptom - somatic (Benasi et al., 2020) | 0.404 |

**Supplementary Table 14**: **Semantic similarity analysis of interoceptive sensibility subscales: MAIA.** Semantic similarity analysis of the MAIA survey subscales was completed with the online Semantic Scale Network tool[19]. The top 15 scales for each MAIA subscale with the highest semantic similarity (cosine similarity) are shown.

| Scales | Similarity |
|---|---|
| **Attention Regulation** | |
| exhibitionistic | 0.511 |
| mindfulness | 0.505 |
| ad involvement-effort | 0.473 |
| introversion | 0.454 |
| social-confidence | 0.417 |
| health orientation-status | 0.401 |
| gregariousness | 0.397 |
| depression-hypochondriasis | 0.397 |
| reading involvement | 0.394 |
| sexual objectification-body | 0.389 |
| organization | 0.386 |
| surgency/extraversion | 0.385 |
| mindful attention awareness | 0.385 |
| road safety judgments | 0.382 |
| pay equity perceptions | 0.365 |
| **Body Listening** | |
| sexual objectification-body | 0.481 |
| transgender congruence | 0.45 |
| consumer perceived community | 0.45 |
| consumer disclosure willingness | 0.448 |
| depression-hypochondriasis | 0.438 |
| general product usefulness | 0.423 |
| disgust-death | 0.42 |
| website responsiveness | 0.416 |
| adolescent femininity ideology | 0.405 |
| negative body talk | 0.4 |
| argument strength | 0.398 |
| website attitudes-security | 0.385 |
| website responsiveness | 0.378 |
| perceived privacy-company's policy | 0.37 |
| body awareness | 0.363 |
| **Emotional Awareness** | |
| sexual objectification-body | 0.566 |
| body awareness | 0.532 |
| transgender congruence | 0.52 |
| depression-hypochondriasis | 0.423 |
| health physical functioning | 0.407 |
| depressive mood | 0.403 |
| competitive somatic anxiety | 0.394 |
| body odor disgust | 0.387 |
| adolescent femininity ideology | 0.369 |
| disgust-death | 0.352 |
| health orientation-status | 0.348 |
| negative body talk | 0.342 |
| sport strategies-relaxe | 0.34 |
| disgust-hygiene | 0.331 |
| personal health orientation | 0.329 |
| **Not Distracting** | |
| gender stereotypes masc physical | 0.411 |
| parkinsons disease-body | 0.381 |
| depression dementia-behav | 0.378 |
| quality life-marrow transplant | 0.372 |
| health orientation-motivation | 0.369 |
| quality life-breast cancer | 0.357 |
| restless legs severity | 0.355 |
| girls education-stem expose | 0.351 |
| quality life-ovarian cancer | 0.344 |
| fibrofatigue | 0.341 |
| pain catastrophizing | 0.336 |
| somatic symptom | 0.331 |

| | |
|---|---|
| patient pain | 0.328 |
| caregiving-positive | 0.324 |
| posttraumatic symptom-arousal | 0.318 |
| **Noticing** | |
| sexual objectification-body | 0.604 |
| body awareness | 0.55 |
| body odor disgust | 0.487 |
| transgender congruence | 0.442 |
| mindfulness | 0.421 |
| health physical functioning | 0.406 |
| depression-hypochondriasis | 0.4 |
| depressive mood | 0.385 |
| competitive somatic anxiety | 0.379 |
| health orientation-status | 0.373 |
| disgust-death | 0.365 |
| sport strategies-relax | 0.354 |
| personal health orientation | 0.344 |
| body esteem | 0.34 |
| adolescent femininity ideology | 0.334 |
| **Not Worrying** | |
| postpartum body stress | 0.376 |
| health orientation-status | 0.375 |
| self-compassion-overidentif | 0.374 |
| depressive mood | 0.365 |
| somatization | 0.34 |
| illness/injury sensitivity | 0.337 |
| state worry | 0.336 |
| somatic symptom | 0.327 |
| health physical functioning | 0.327 |
| emotion regulation-non-accept | 0.323 |
| gender stereotypes masc physical | 0.322 |
| girls education-stem expose | 0.321 |
| transgender congruence | 0.312 |
| parkinsons disease-body | 0.309 |
| consumer sympathy | 0.309 |
| **Self Regulation** | |
| body state-freedom | 0.429 |
| sexual objectification-body | 0.428 |
| body state-confinement | 0.416 |
| health physical functioning | 0.407 |
| transgender congruence | 0.405 |
| competitive somatic anxiety | 0.394 |
| mindfulness | 0.373 |
| strain | 0.357 |
| sport strategies-relax | 0.348 |
| body countertransference | 0.341 |
| somatic symptom | 0.335 |
| somatization | 0.334 |
| bodyparts dissatisfaction | 0.334 |
| binge eating-arousal | 0.332 |
| body awareness | 0.329 |
| **Trusting** | |
| sexual objectification-body | 0.609 |
| depression hypochondriasis | 0.58 |
| adolescent femininity ideology | 0.534 |
| disgust-death | 0.533 |
| transgender congruence | 0.529 |
| competitive somatic anxiety | 0.472 |
| negative body talk | 0.47 |
| body awareness | 0.406 |
| body control | 0.403 |
| physical attractiveness | 0.402 |
| somatization | 0.394 |
| health orientation status | 0.393 |
| health physical functioning | 0.391 |
| self-image-body | 0.37 |
| eating disorders | 0.368 |

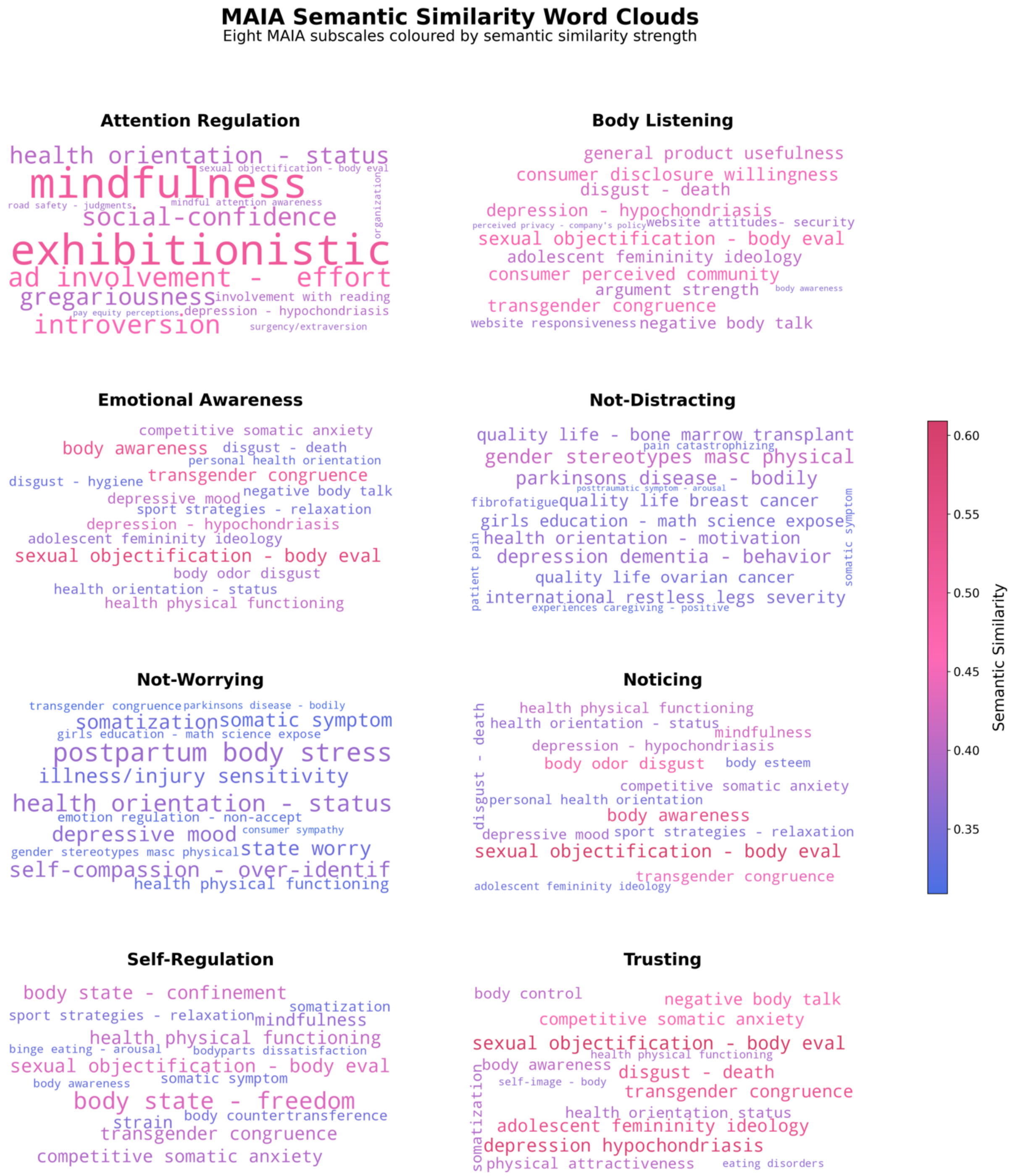


**Supplementary Figure 20: Latent semantic similarity analysis of eight MAIA survey subscales.** Word cloud of latent semantic similarity analysis of the Multidimensional Assessment of Interoceptive Awareness (MAIA) questionnaire items (for each subscale) in relation to other psychological scales, using the online Semantic Scale Network tool[19]. The size and colour of the words represent the cosine similarity of the MAIA subscale to other psychological questionnaires, with larger pink/red words indicating higher similarity.

**Supplementary Table 15**: Individual items of the Multidimensional Assessment of Interoceptive Awareness (MAIA) survey which measure interoceptive sensibility (eight subscales shown as subheadings)[1]. The instructions were as follows: “Below you will find a list of statements. Please indicate how often each statement applies to you GENERALLY IN DAILY LIFE.”. Participants rated their responses on a scale from 0 (Never) to 5 (Always). Items were averaged for each subscale score, with items 5 to 9 reverse-coded.

| Item | Question |
|---|---|
| | **Noticing** *(Awareness of uncomfortable, comfortable, and neutral body sensations)* |
| maia_1 | When I am tense I notice where the tension is located in my body |
| maia_2 | I notice when I am uncomfortable in my body |
| maia_3 | I notice where in my body I am comfortable |
| maia_4 | I notice changes in my breathing, such as whether it slows down or speeds up |
| | **Not-Distracting** *(Tendency not to ignore or distract oneself from sensations of pain or discomfort )* |
| maia_5 | I do not notice (I ignore) physical tension or discomfort until they become more severe |
| maia_6 | I distract myself from sensations of discomfort |
| maia_7 | When I feel pain or discomfort, I try to power through it |
| | **Not-Worrying** *(Tendency not to worry or experience emotional distress with sensations of pain or discomfort)* |
| maia_8 | When I feel physical pain, I become upset |
| maia_9 | I start to worry that something is wrong if I feel any discomfort |
| maia_10 | I can notice an unpleasant body sensation without worrying about it |
| | **Attention Regulation** *(Ability to sustain and control attention to body sensations)* |
| maia_11 | I can pay attention to my breath without being distracted by things happening around me |
| maia_12 | I can maintain awareness of my inner bodily sensations even when there is a lot going on around me |
| maia_13 | When I am in conversation with someone, I can pay attention to my posture |
| maia_14 | I can return awareness to my body if I am distracted |
| maia_15 | I can refocus my attention from thinking to sensing my body |
| maia_16 | I can maintain awareness of my whole body even when a part of me is in pain or discomfort |
| maia_17 | I am able to consciously focus on my body as a whole |
| | **Emotional Awareness** *(Awareness of the connection between body sensations and emotional states)* |
| maia_18 | I notice how my body changes when I am angry |
| maia_19 | When something is wrong in my life I can feel it in my body |
| maia_20 | I notice that my body feels different after a peaceful experience |
| maia_21 | I notice that my breathing becomes free and easy when I feel comfortable |
| maia_22 | I notice how my body changes when I feel happy / joyful |
| | **Self-Regulation** *(Ability to regulate distress by attention to body sensations)* |
| maia_23 | When I feel overwhelmed I can find a calm place inside |
| maia_24 | When I bring awareness to my body I feel a sense of calm |
| maia_25 | I can use my breath to reduce tension |
| maia_26 | When I am caught up in thoughts, I can calm my mind by focusing on my body/breathing |
| | **Body Listening** *(Active listening to the body for insight)* |
| maia_27 | I listen for information from my body about my emotional state |

| | |
|---|---|
| maia_28 | When I am upset, I take time to explore how my body feels |
| maia_29 | I listen to my body to inform me about what to do |
| | **Trusting** <br> *(Experience of one's body as safe and trustworthy)* |
| maia_30 | I am at home in my body |
| maia_31 | I feel my body is a safe place |
| maia_32 | I trust my body sensations |

**Supplementary Table 16**: Individual items included in the Mental Health EFA Models (two items coloured in red were excluded due to a larger number of missing values, one item coloured in orange was excluded as Kaiser-Meyer-Olkin test < 0.5).

| Item | Question |
|---|---|
| aq10_1 | I often notice small sounds when others do not |
| aq10_2 | I usually concentrate more on the whole picture, rather than the small details |
| aq10_3 | I find it easy to do more than one thing at once |
| aq10_4 | If there is an interruption, I can switch back to what I was doing very quickly |
| aq10_5 | I find it easy to 'read between the lines' when someone is talking to me |
| aq10_6 | I know how to tell if someone listening to me is getting bored |
| aq10_7 | When I'm reading a story I find it difficult to work out the characters' intentions |
| aq10_8 | I like to collect information about categories of things (e.g. types of car, types of bird, types of train, types of plant etc) |
| aq10_9 | I find it easy to work out what someone is thinking or feeling just by looking at their face |
| aq10_10 | I find it difficult to work out people's intentions |
| asrs_a_1 | How often do you have trouble wrapping up the final details of a project, once the challenging parts have been done? |
| asrs_a_2 | How often do you have difficulty getting things in order when you have to do a task that requires organization? |
| asrs_a_3 | How often do you have problems remembering appointments or obligations? |
| asrs_a_4 | When you have a task that requires a lot of thought, how often do you avoid or delay getting started? |
| asrs_a_5 | How often do you fidget or squirm with your hands or feet when you have to sit down for a long time? |
| asrs_a_6 | How often do you feel overly active and compelled to do things, like you were driven by a motor? |
| asrs_b_1 | How often do you make careless mistakes when you have to work on a boring or difficult project? |
| asrs_b_2 | How often do you have difficulty keeping your attention when you are doing boring or repetitive work? |
| asrs_b_3 | How often do you have difficulty concentrating on what people say to you, even when they are speaking to you directly? |
| asrs_b_4 | How often do you misplace or have difficulty finding things at home or at work? |
| asrs_b_5 | How often are you distracted by activity or noise around you? |
| asrs_b_6 | How often do you leave your seat in meetings or other situations in which you are expected to remain seated? |
| asrs_b_7 | How often do you feel restless or fidgety? |
| asrs_b_8 | How often do you have difficulty unwinding and relaxing when you have time to yourself? |
| asrs_b_9 | How often do you find yourself talking too much when you are in social situations? |
| asrs_b_10 | When you're in a conversation, how often do you find yourself finishing the sentences of the people you are talking to, before they can finish them themselves? |
| asrs_b_11 | How often do you have difficulty waiting your turn in situations when turn taking is required? |
| asrs_b_12 | How often do you interrupt others when they are busy? |

| | |
|---|---|
| mdi_1 | Have you felt low in spirits or sad? |
| mdi_2 | Have you lost interest in your daily activities? |
| mdi_3 | Have you felt lacking in energy and strength? |
| mdi_4 | Have you felt less self-confident? |
| mdi_5 | Have you had a bad conscience or feelings of guilt? |
| mdi_6 | Have you felt that life wasn't worth living? |
| mdi_7 | Have you had difficulty in concentrating, e.g. when reading the newspaper or watching TV? |
| mdi_8a | Have you felt very restless? |
| mdi_8b | Have you felt subdued or slowed down? |
| mdi_9a | Have you been sleeping too little? |
| mdi_9b | Have you been sleeping too much? |
| mdi_10a | Have you suffered from reduced appetite? |
| mdi_10b | Have you suffered from increased appetite? |
| phq_9_1 | Little interest or pleasure in doing things |
| phq_9_2 | Feeling down, depressed, or hopeless |
| phq_9_3 | Trouble falling or staying asleep, or sleeping too much |
| phq_9_4 | Feeling tired or having little energy |
| phq_9_5 | Poor appetite or overeating |
| phq_9_6 | Feeling bad about yourself — or that you are a failure or have let yourself or your family down |
| phq_9_7 | Trouble concentrating on things, such as reading the newspaper or watching television |
| phq_9_8 | Moving or speaking so slowly that other people could have noticed? Or the opposite — being so fidgety or restless that you have been moving around a lot more than usual |
| phq_9_9 | Thoughts that you would be better off dead or of hurting yourself in some way |
| phq_9_10 | If you checked off any problems, how difficult have these problems made it for you to do your work, take care of things at home, or get along with other people? |
| phq15_1 | Stomach pain |
| phq15_2 | Back pain |
| phq15_3 | Pain in your arms, legs, or joints (knees, hips, etc.) |
| phq15_4 | Menstrual cramps or other problems with your periods (women only) |
| phq15_5 | Pain or problems during sexual intercourse |
| phq15_6 | Headaches |
| phq15_7 | Chest pain |
| phq15_8 | Dizziness |
| phq15_9 | Fainting spells |
| phq15_10 | Feeling your heart pound or race |
| phq15_11 | Shortness of breath |
| phq15_12 | Constipation, loose bowels, or diarrhea |
| phq15_13 | Nausea, gas, or indigestion |
| phq15_14 | Trouble falling asleep, sleeping or sleeping too much |
| phq15_15 | Felt tired or only had little energy |
| pss_1 | In the last month, how often have you been upset because of something that happened unexpectedly? |
| pss_2 | In the last month, how often have you felt that you were unable to control the important things in your life? |
| pss_3 | In the last month, how often have you felt nervous and stressed? |

| | |
|---|---|
| pss_4 | In the last month, how often have you felt confident about your ability to handle your personal problems? |
| pss_5 | In the last month, how often have you felt that things were going your way? |
| pss_6 | In the last month, how often have you found that you could not cope with all the things that you had to do? |
| pss_7 | In the last month, how often have you been able to control irritations in your life? |
| pss_8 | In the last month, how often have you felt that you were on top of things? |
| pss_9 | In the last month, how often have you been angered because of things that happened that were outside of your control? |
| pss_10 | In the last month, how often have you felt difficulties were piling up so high that you could not overcome them? |
| sias_1 | I get nervous if I have to speak with someone in authority (teacher, boss, etc.). |
| sias_2 | I have difficulty making eye contact with others. |
| sias_3 | I become tense if I have to talk about myself or my feelings. |
| sias_4 | I find it difficult to mix comfortably with the people I work with. |
| sias_5 | I find it easy to make friends my own age. |
| sias_6 | I tense up if I meet an acquaintance in the street. |
| sias_7 | When mixing socially, I am uncomfortable. |
| sias_8 | I feel tense if I am alone with just one other person. |
| sias_9 | I am at ease meeting people at parties, etc. |
| sias_10 | I have difficulty talking with other people. |
| sias_11 | I find it easy to think of things to talk about. |
| sias_12 | I worry about expressing myself in case I appear awkward. |
| sias_13 | I find it difficult to disagree with another's point of view. |
| sias_14 | I have difficulty talking to attractive persons of the opposite sex. |
| sias_15 | I find myself worrying that I won't know what to say in social situations. |
| sias_16 | I am nervous mixing with people I don't know well. |
| sias_17 | I feel I'll say something embarrassing when talking. |
| sias_18 | When mixing in a group, I find myself worrying I will be ignored. |
| sias_19 | I am tense mixing in a group. |
| sias_20 | I am unsure whether to greet someone I know only slightly." |
| stai_21 | I feel pleasant |
| stai_22 | I feel nervous and restless |
| stai_23 | I feel satisfied with myself |
| stai_24 | I wish I could be as happy as others seem to be |
| stai_25 | I feel like a failure |
| stai_26 | I feel rested |
| stai_27 | I am 'calm, cool, and collected' |
| stai_28 | I feel that difficulties are piling up so that I cannot overcome them |
| stai_29 | I worry too much over something that really doesn't matter |
| stai_30 | I am happy |
| stai_31 | I have disturbing thoughts |
| stai_32 | I lack self-confidence |
| stai_33 | I feel secure |
| stai_34 | I make decisions easily |
| stai_35 | I feel inadequate |
| stai_36 | I am content |
| stai_37 | Some unimportant thought runs through my mind and bothers me |

| | |
|---|---|
| stai_38 | I take disappointments so keenly that I can't put them out of my mind |
| stai_39 | I am a steady person |
| stai_40 | I get in a state of tension or turmoil as I think over my recent concerns and interests |

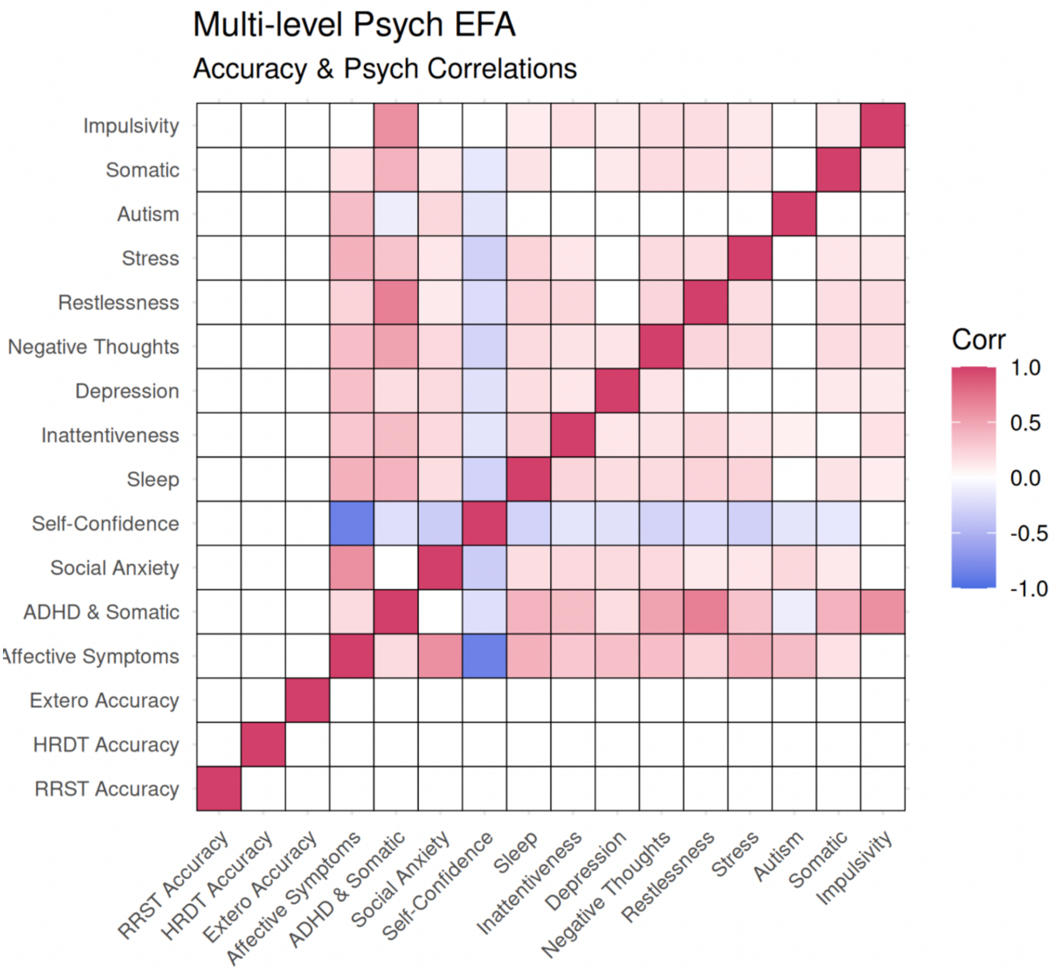


**Supplementary Figure 21: Correlations between task accuracy and mental health dimensions.** Cross Spearman correlations across mental health dimensions and interoception task accuracy across cardiac (HRDT), respiratory (RRST), and auditory (Extero) sensory modalities. Mental health factors encompass 11 lower-level and two higher-level dimensions. Accuracy is estimated as the number of correct trials divided by the number of trials. In red are positive correlations and in blue are negative correlations. The upper triangle depicts correlations that survive false discovery rate correction for multiple comparisons (Benjamini-Hochberg procedure at < 0.05 shown in upper triangle-coloured boxes), whereas the lower triangle depicts correlation coefficients significant at the uncorrected threshold (uncorrected p < .05; lower triangle-coloured boxes).
